# Autoencoder-based phenotyping of ophthalmic images highlights genetic loci influencing retinal morphology and provides informative biomarkers

**DOI:** 10.1101/2023.06.15.23291410

**Authors:** Panagiotis I. Sergouniotis, Adam Diakite, Kumar Gaurav, Ewan Birney, Tomas Fitzgerald

**Affiliations:** European Molecular Biology Laboratory, European Bioinformatics Institute (EMBL-EBI), Wellcome Genome Campus, Cambridge, UK; Division of Evolution, Infection and Genomics, School of Biological Sciences, Faculty of Biology, Medicine and Health, University of Manchester, Manchester, UK; Manchester Centre for Genomic Medicine, Saint Mary’s Hospital, Manchester University NHS Foundation Trust, Manchester, UK; Manchester Royal Eye Hospital, Manchester University NHS Foundation Trust, Manchester, UK

**Keywords:** autoencoder, U-Net, retinal imaging, optical coherence tomography, imaging-derived phenotypes

## Abstract

Genome-wide association studies (GWAS) have been remarkably successful in identifying associations between genetic variation and imaging-derived phenotypes. To date, the main focus of these analyses has been established, clinically-used imaging features. Here, we sought to investigate if deep learning approaches can help detect more nuanced patterns of image variability. To this end, we used an autoencoder to represent retinal optical coherence tomography (OCT) images from 31,135 UK Biobank participants. For each study subject, we obtained a 64-dimensional vector representing features of retinal structure. GWAS of these autoencoder-derived imaging parameters identified 118 statistically significant loci; 17 of these associations also reached genome-wide significance in a replication analysis that included 10,409 UK Biobank volunteers. These loci encompassed variants previously linked with retinal thickness measurements, ophthalmic disorders and/or neurodegenerative conditions (including dementia). Notably, the generated retinal phenotypes were found to contribute to predictive models for glaucoma and cardiovascular disorders. Overall, we demonstrate that self-supervised phenotyping of OCT images enhances the discoverability of genetic factors influencing retinal morphology and provides epidemiologically informative biomarkers.

## INTRODUCTION

Imaging technologies have greatly enhanced the scope and precision of phenotype discovery. A wide range of imaging-derived phenotypes are easily amenable to human identification and are routinely used in biomedical contexts, including in clinical practice (Oren 2020). However, to capture the complexity of human biology, there is a need to go beyond traditional clinically-focused and/or expert-curated imaging features (Gong 2022).

Artificial neural networks (ANN) are machine-learning models inspired by information processing in biological neural networks (LeCun 2015; Hinton 2018; Hasson 2020). ANNs can be used to extract granular information from images without introducing certain biases associated with human curation. An autoencoder is a type of ANN that is designed to transform an input set of data into a lower-dimensional code (*i.e.* a set of latent space variables or ‘embeddings’) and then to recreate the input from the encoded representation (Hinton 2006; Michelucci 2022). Broadly, autoencoders can be used to efficiently compress an image by identifying the key features that lead to optimal reconstruction performance.

The most optically accessible part of the central nervous system is the retina, the multilayered tissue that lines the back of the eyes. The retina is particularly vulnerable to disease, and disruption of its normal architecture (e.g. in conditions like age-related macular degeneration or glaucoma) can lead to visual disability (Sheffield 2011; Zhao 2023).

Examination of the retina relies, to a great extent, on imaging, especially the use of optical coherence tomography (OCT). OCT is a non-invasive, non-contact method for cross-sectional imaging that has a resolution approaching that of histopathology (Bouma 2022). Application of ANN-based algorithms in OCT image processing is attracting increasing attention with key advantages including the rapid speed, high consistency and quantitative nature of the analyses (De Fauw 2018; Yim 2020; Keenan 2021; Diaz-Pinto 2022).

To date, genetic studies of imaging phenotypes have mostly focused on features associated with long-established clinical diagnostic processes (Xie 2022). In our own previous work, we used standardised OCT-derived thickness measurements of the inner (Currant 2021) and outer (Currant 2023) retinal layers to good effect, discovering new genetic associations and exploring relationships with disease. Here we performed genomic analyses on OCT imaging phenotypes extracted using a self-supervised autoencoder-based approach. We highlight the autoencoder’s ability to derive biologically meaningful phenotypes (with association to genetic variants not seen in previous studies), and to contribute to predictive models for health outcomes such as glaucoma and cardiovascular conditions.

## RESULTS

### Obtaining autoencoder-derived phenotypes from OCT images

We used OCT images from the UK Biobank, a biomedical resource containing genomic and health information from >500,000 individuals (Bycroft 2018). After applying standard genetic and OCT quality control filters (Patel 2016; Currant 2021), we defined a subset of the UK Biobank population that (i) can be considered genetically well-mixed (*i.e.* includes participants that were assigned by genotype principal component analysis (PCA) to a cluster with subjects of mostly European-like ancestries) and (ii) only contains individuals with high-quality OCT images. This cohort included 31,135 individuals and had a similar sex and age profile to the overall UK Biobank population (Currant 2023) (**Supplementary** Fig.1). Most study subjects were female (54%) and self-identified as White British (91%). The mean age at OCT imaging was 56 years (standard deviation: 8 years).

Study subjects had an OCT ‘volume scan’ of the central retina in each eye. Each volume scan contained 128 cross-sectional images and was generated using a horizontal raster scanning protocol. To extract thickness information and to compress these 128 images into a single retinal ‘thickness map’, we developed an ANN algorithm involving a U-Net architecture (Ronneberger 2015) (**Fig.1; Methods**).

**Figure 1.**
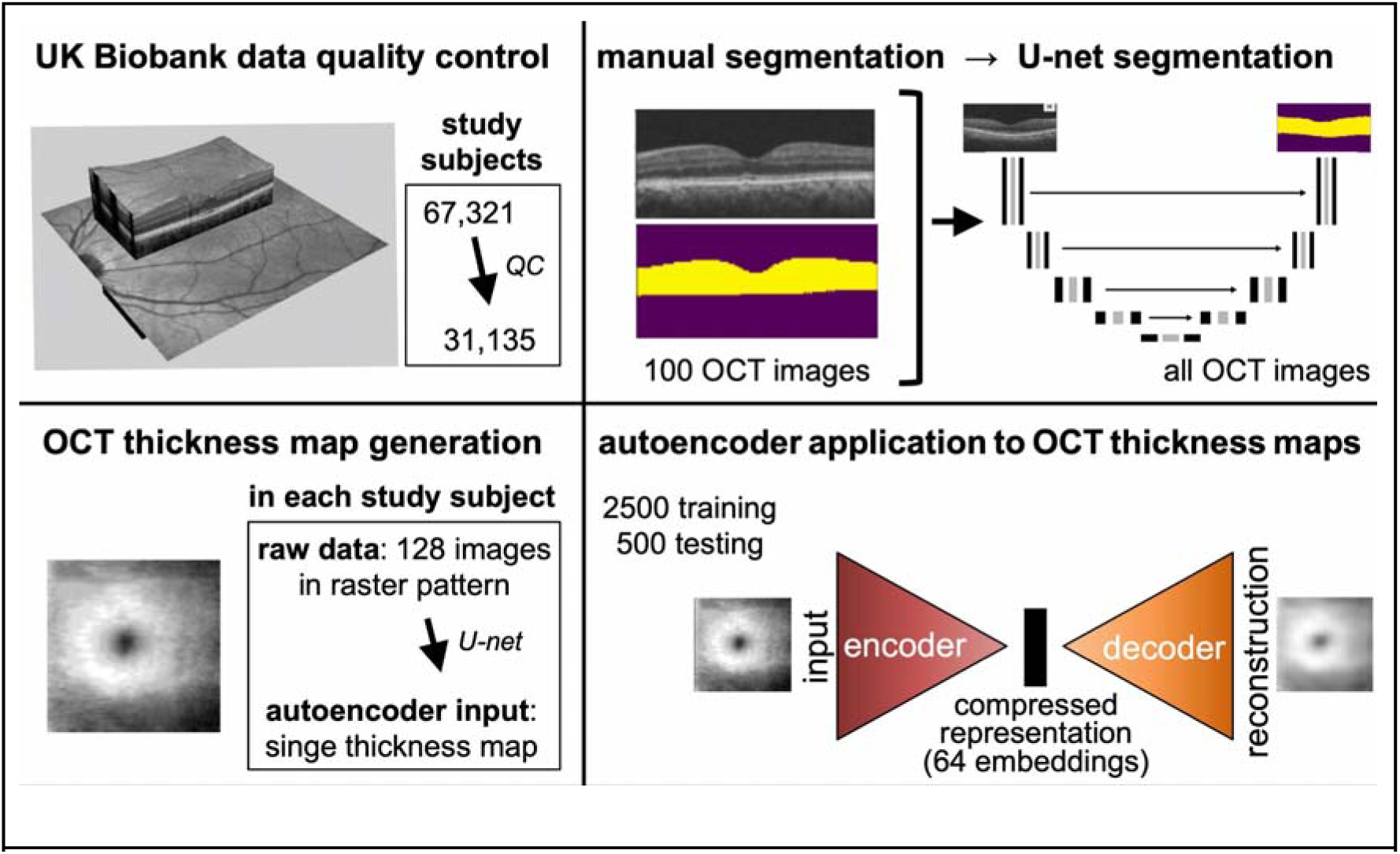
Outline of the experimental approach. OCT images from the central retinae of 67,321 UK Biobank participants were analysed. After applying quality control (QC) filters considering genetic information and image quality, a cohort of 31,135 study subjects was identified. Aiming to generate retinal ‘thickness maps’ for these individuals, OCT image segmentation was performed using an artificial neural network (U-Net) approach. In brief, 100 OCT images were manually segmented and the generated segmentation masks (examples shown in yellow) were used as input to the U-Net which subsequently segmented all other images. This allowed conversion of the 128 cross-sectional images obtained from each tested eye into a single thickness map image. The thickness maps of the left eyes were then used as input to an autoencoder. This was trained utilising 2500 training and 500 test images. The output of the embedding network was designed to be a 64-dimensional vector (*i.e.* 64 variables were obtained for each study subject). These 64 autoencoder-derived embeddings were then used for genetic association studies, correlation analyses and predictive modelling.

The 31,135 left eye retinal thickness maps that we generated were then used as input to an autoencoder. This was trained end-to-end for 150 epochs utilising 2500 training and 500 test images. We explored various embedding dimensionalities and opted for a 64-dimensional vector (*i.e.* the latent space or ‘bottleneck layer’ contained 64 features) (**Fig.1; Methods**). It has been previously shown that this autoencoder architecture can sufficiently represent datasets of similar complexity (Schroff 2015; Song 2015). A reconstruction error of 0.0037 was obtained (**Supplementary** Fig.2).

The univariate distributions of the 64 embeddings are shown in **Supplementary** Fig.3. Mostly unimodal or bimodal distributions were observed.

To create an alternative representation allowing information to be combined across different variables within the latent space, we used the 64 embeddings as input to a PCA. The first 25 principal components, representing 98.5% of the variance within the embeddings, were studied further and used for genetic association tests.

### Genetic association studies of autoencoder-derived OCT phenotypes

To look for genetic factors associated with the obtained autoencoder-derived embedded features (*i.e.* the 64 embeddings and the first 25 embedding-related principal components), we performed common-variant genome-wide association studies (GWAS). We used REGENIE (Mbatchou 2021) and incorporated the following set of covariates into the model: age at recruitment, sex, height, weight, refractive error and genetic principal components 1 to 20. Notably, each embedded feature was inverse rank normalised prior to performing genetic association testing. As we anticipated a degree of correlation between autoencoder-derived phenotypes, we also conducted a multi-trait meta-analysis using MTAG (Turley 2018). This involved identifying genetically correlated embeddings and leveraging these relationships to obtain adjusted GWAS results for each of the 64 embeddings (**Methods**).

Overall, 418,312 association signals from 17,022 common variants reached the genome-wide significance threshold (p-value < 5 x 10^-8^) (**Table 1**; **Fig.2**). These merged into 239 lead loci following analysis with GCTA-COJO (conditional and joint multiple-variant analysis) (Yang 2012) (**Supplementary Table 1**); 118 of these remained significant when a conservative/higher (“study-wide”) threshold was used to account for all the different association routes that were utilised (p-value < 3.2 x 10^-10^ following Bonferroni correction for 153 tests).

**Figure 2.**
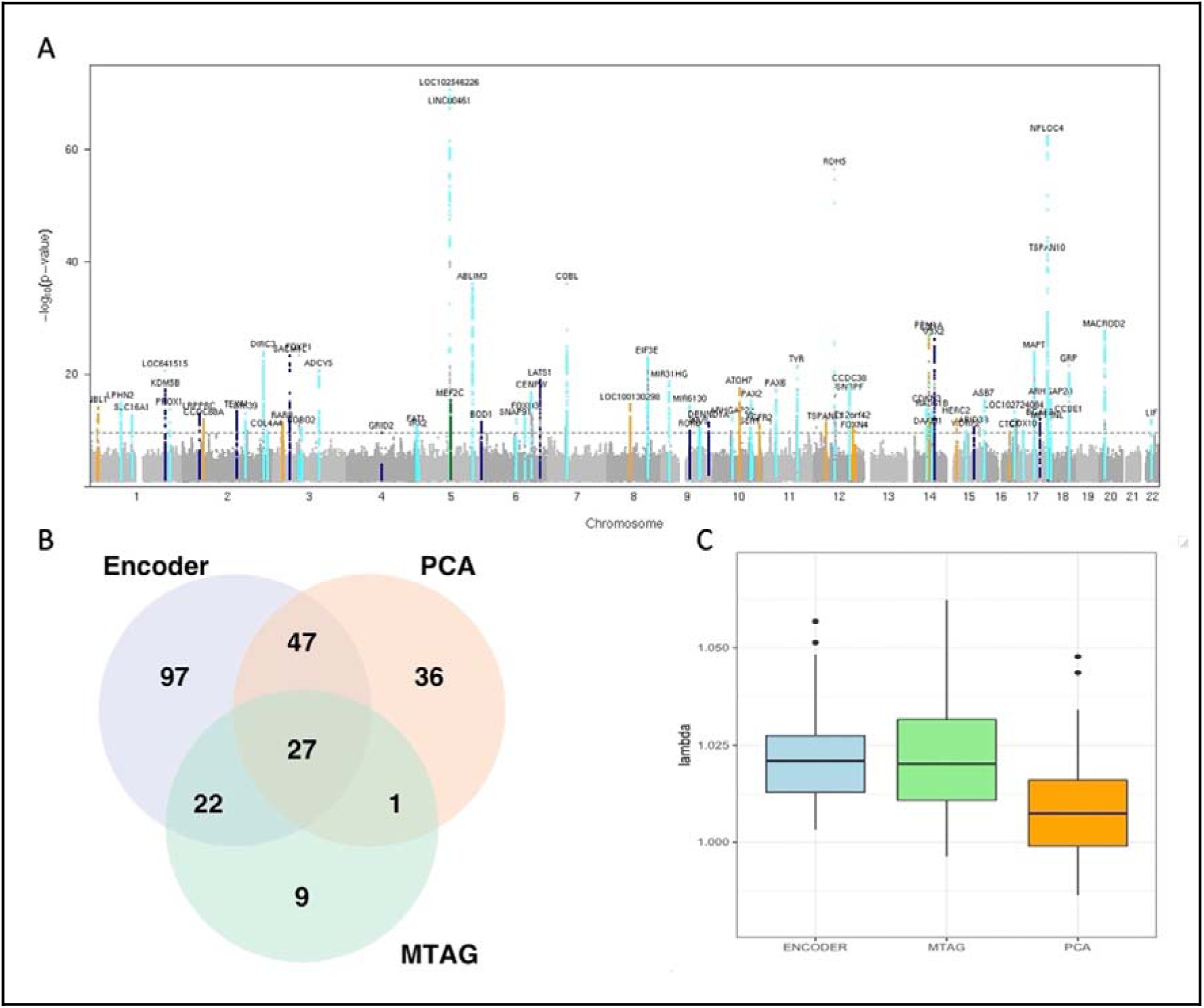
Genome-wide association studies of autoencoder-derived retinal OCT phenotypes (primary analysis). [A] Manhattan plot showing the p-values obtained from common-variant GWAS of embedded features (64 embeddings and first 25 embedding-related principal components). Signals that reached genome-wide significance (p-value < 5 x 10^-8^) only in embedding variable analyses are highlighted with dark blue. Signals that reached genome-wide significance only in analyses of embedding-related principal components are highlighted with orange. Signals that reach genome-wide significance only in MTAG of embedding variables are highlighted with green. All other genome-wide significant signals are highlighted with cyan. [B] Venn diagram showing the overlap of lead signals among: conventional GWAS of the 64 embeddings (“encoder” group in light blue); MTAG of the 64 embeddings (“MTAG” group in light green) and conventional GWAS of the first 25 embedding-related principal components (“PCA” group in light orange). [C] Genomic inflation factor lamda (λ) for 64 embedding-, 64 MTAG- and 25 PCA-GWAS (median λGC = 1.016).

**Table 1.** Comparative analyses of conventional and MTAG GWAS results (primary analysis)

|  |  |  |  |
| --- | --- | --- | --- |
|  | **GWAS**<br>**64 embeddings** | **MTAG**<br>**64 embeddings** | **GWAS**<br>**25 PCAs** |
| Total genetic variants (p-value < 5 x 10<sup>-8</sup>) | 14,885 | 9,520 | 11,075 |
| Lead genetic variants (p-value < 5 x 10<sup>-8</sup>) | 443 | 99 | 157 |
| GWAS, genome-wide association study; MTAG, multi-trait analysis of GWAS; PCA, principal component analysis. |  |  |  |

A replication study was conducted using OCT scans from 10,409 UK Biobank participants that were not included in the primary analysis. There was a high level of concordance in the findings of the two association studies (**Supplementary** Fig.4). A total of 17 loci passed both the conservative study-wide threshold (p-value < 3.2 x 10-10) in the primary analysis and the conventional genome-wide threshold (p-value < 5 x 10-8) in the replication study. Most of these loci encompass variants previously linked to retinal layer thickness parameters (including around *LINC00461, TSPAN10* and *COBL* (Gao 2019; Currant 2021; Currant 2023) while a subset of them has also been linked to monogenic retinal disorders (including *RDH5* [retinal dystrophy], *TYR* [albinism] and *GNB3* [congenital stationary night blindness]) (**Table 2; Supplementary Table 1**).

**Table 2.** Summary of the 10 top-ranking loci associated with autoencoder-derived retinal OCT phenotypes.

|  |  |  |  |  |  |  |
| --- | --- | --- | --- | --- | --- | --- |
| **top-ranking**<br>**common**<br>**variant in**<br>**locus** | **chr:**<br>**position**<br>**(GRCh37)** | **key**<br>**gene(s)** | **allele**<br>**freq**<br>**(UKB)** | **minimum**<br>**p-value** | **embeddin**<br>**gs with**<br>**significant**<br>**result for**<br>**the locus** | **selected previous**<br>**association(s) with the**<br>**detected significant**<br>**variants in the locus**<br>**GWAS catalog; (PanelApp)** |
| rs17421627 | 5:87847586 | *LINC00461* | 0.07 | 4 x 10<sup>-68</sup> | 83 | retinal thickness measurements, retinal vascular fractal density |
| rs62075722 | 17:79611271 | *TSPAN10*<br>*/NPLOC4*<br>*/PDE6G* | 0.65 | 1 x 10<sup>-62</sup> | 83 | retinal thickness measurements, refractive error, eye colour, hair colour |
| rs3138142 | 12:56115585 | *RDH5*<br>*/CD63* | 0.24 | 1 x 10<sup>-56</sup> | 91 | retinal thickness measurements, refractive error, retinal vascular fractal density; (retinal dystrophy) |
| rs13171669 | 5:148601243 | *AFAP1L1*<br>*/ABLIM3* | 0.43 | 1 x 10<sup>-36</sup> | 84 | retinal thickness measurements, height, waste-hip ratio, lung function |
| rs12719025 | 7:51100190 | *COBL* | 0.46 | 1 x 10<sup>-36</sup> | 113 | retinal thickness measurements, refractive error |
| rs33912345 | 14:60976537 | *SIX6*<br>*/C14orf39*<br>*/PPPM1A* | 0.61 | 4 x 10<sup>-28</sup> | 6 | retinal thickness measurements, glaucoma, height; (ocular malformations) |
| rs887595 | 14:74666641 | *VSX2*/<br>*LIN52* | 0.82 | 6 x 10<sup>-27</sup> | 85 | retinal thickness measurements; (microphthalmia) |
| rs17279437 | 3:45814094 | *SLC6A20* | 0.11 | 8 x 10<sup>-24</sup> | 33 | retinal thickness measurements, macular telangiectasia, brain measurements, metabolite measurements; (hyperglycynuria) |
| rs1042602 | 11:88911696 | *TYR* | 0.37 | 5 x 10<sup>-22</sup> | 29 | retinal thickness measurements, brain measurements, skin colour, hair colour; (albinism) |
| rs62175360 | 2:218520035 | *DIRC3* | 0.07 | $9 \times 10^{-22}$ | 21 | retinal thickness measurements, optic disc measurements, brain measurements, metabolite measurements, height cancer |
| The above loci were identified after selecting fine mapped variants that had a p-value $< 3.2 \times 10^{-10}$ in the primary analysis and a p-value $< 5 \times 10^{-8}$ in the replication study. Manual inspection of linkage disequilibrium patterns was subsequently performed to further refine the signals and the 10 loci with the lowest p-value were selected. UKB, UK Biobank. |  |  |  |  |  |  |

For each of the 118 lead loci that were found to be significant in the primary analysis (p-value < 3.2 x 10^-10^), we compared the retinal thickness maps of heterozygotes for the key variant to that of homozygotes. Interestingly, some genetic alterations appeared to have recessive effects (e.g. rs62075722) while others appeared to have dominant effects (e.g. rs11051131); topographical variation was also noted (**Supplementary File 1**).

Our primary analysis identified notable associations between multiple embeddings and a locus encompassing the *MAPT* (microtubule-associated protein tau) gene. The detected signal appears to be driven by a common ancestral genomic inversion at 17q21.31 (**Fig.3A**) (Stefansson 2005; Espinosa 2023). Using the pattern of alternative alleles across this genomic region, we were able to classify 487,409 UK Biobank participants as either reference:reference (no inversion), reference:inversion (heterozygous inversion) or inversion:inversion (homozygous inversion) (**Fig.3B**). Similarly to previous studies (Steinberg 2013), we found that the 17q21.31 inversion is common in individuals of European-like ancestries, rare in individuals of African-like ancestries and very rare in Asian-like populations (allele frequency of 0.22, 0.01 and 0.004 respectively). When we compared the retinal thickness profiles between study subjects that carry heterozygous and homozygous inversion genotypes, we found that the 17q21.31 inversion appears to affect retinal thickness in an apparently recessive pattern (**Fig.3C**). We then performed a phenome-wide association study (PheWAS) of the 17q21.31 inversion using disease-related ICD10 codes. After Bonferroni correction, we found six statistically significant signals for ICD10 codes, including one for Parkinson disease (G20; p-value = 5.3 x 10^-7^; beta −0.61) (**Fig.3D**).

**Figure 3.**
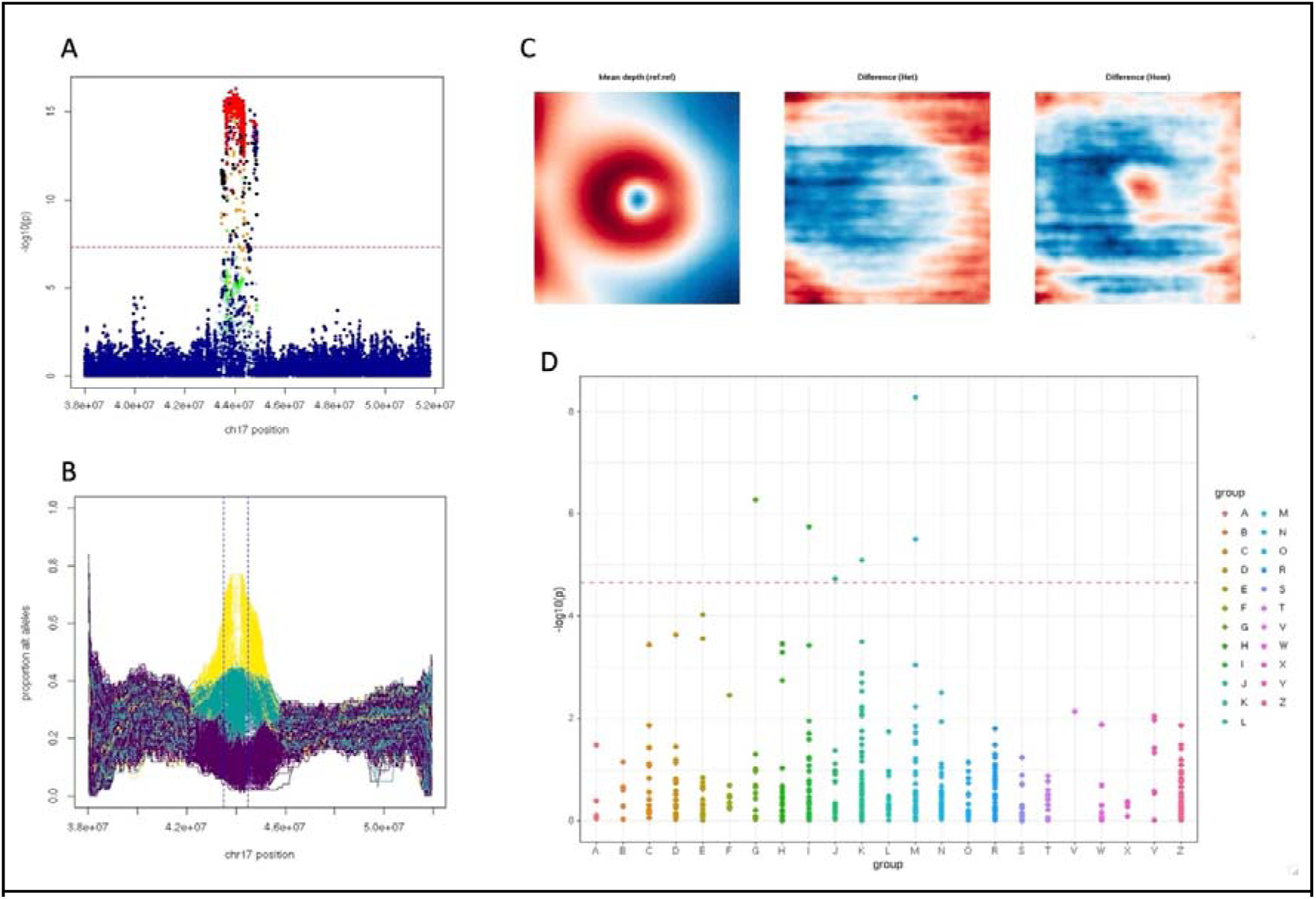
Analysis of the chromosome 17q21.31 inversion association signal. [A] Genetic association study result highlighting a group of 2,936 common variants that passed the genome-wide significance threshold for MTAG of embedding no.21. The genetic alterations are coloured based on their linkage disequilibrium (LD; R^2^) relationship to the inversion genotype. [B] Classification of the inversion status based on the pattern of alternative alleles across the 17q21.31 region for 487,409 UK Biobank participants. [C] Left eye retinal thickness maps showing the difference in retinal structure between individuals with different inversion-related alleles. Left: mean depth (thickness) representation for reference:reference (no inversion) alleles. Middle: difference between image mean for reference:reference and image mean for reference:inversion (heterozygous inversion) genotypes. Right: difference between image mean for reference:reference and image mean for inversion:inversion (homozygous inversion) genotypes. A paracentral area of differential retinal thickness can only be visualised in the reference-to-homozygous difference map (in keeping with a recessive effect). [D] Phenome-wide associations for the inversion genotype against 454 ICD10 disease codes for which there were >1000 cases in the UK Biobank cohort (when only data obtained after the date of OCT image acquisition were considered); six codes (M16, G20, I84, M20, K60, J84) remained significant after Bonferroni correction; −log10 p-values are shown grouped by high-level ICD10 category.

### Investigating how autoencoder-derived OCT phenotypes are related between them and with other retinal traits and diseases

To gain insights into the nature of the autoencoder-derived embedded features, we performed correlation and logistic regression analyses. First, we examined the direct pairwise correlation between the 64 embeddings; a few prominent clusters were noted (**Fig.4A - upper triangle**). Then we looked at genetic correlation (**Fig.4A - lower triangle**); a notable observation was the discrepancy between the degree of direct and genetic correlation for many groups of embeddings. This suggests that although the latent space is complex and includes (linearly) correlated features, the different embeddings are able to represent discrete factors related to different aspects of retinal morphology genetics.

**Figure 4.**
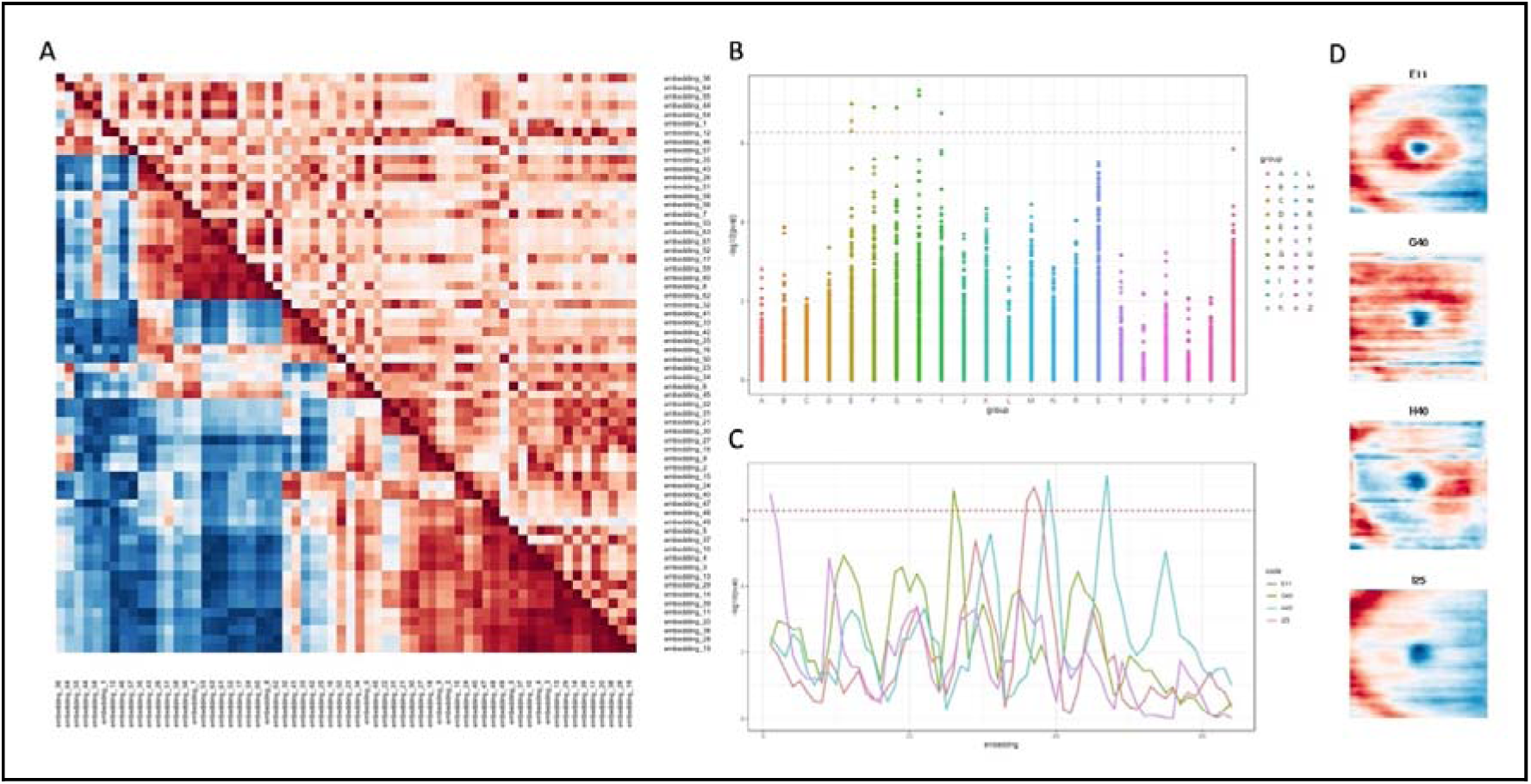
Correlation and logistic regression analyses of autoencoder-derived retinal OCT phenotypes. [A] Direct (upper triangle) and genetic (lower triangle) correlations among embedded features (64 embeddings). [B] Logistic regression analysis of the 64 embeddings against high-level ICD10 disease codes; only data obtained after the date of OCT image acquisition were included and only ICD10 codes for which there were >1000 cases in the UK Biobank cohort were considered; sex, age, height and weight were factored in as covariates. A total of 8 signals for 5 distinct ICD10 codes remained significant after Bonferroni correction: E11 (3), G40 (1), H40 (2), I25 (1), F10 (1). [C] Graph showing which specific embeddings were significantly correlated with the lead signals of the logistic regression analysis, *i.e.* non-insulin-dependent diabetes (E11), epilepsy (G40), glaucoma (H40) and chronic ischaemic heart disease (I25); −log10 p-values are shown for all 64 embedded features. [D] Left eye retinal thickness maps showing the difference in retinal structure between UK Biobank participants who were diagnosed with non-insulin-dependent diabetes (E11; first row), epilepsy (G40; second row), glaucoma (H40; third row) and chronic ischaemic heart disease (I25; fourth row) after having an OCT scan against the groups of individuals that have not been assigned the relevant ICD10 codes.

We subsequently investigated the relationship between the 64 embedded features and a set of traits and disease codes (ICD10) that are available in the UK Biobank dataset. Unsurprisingly, most embeddings correlated with retinal layer thickness parameters (**Supplementary** Fig.5). We then used a logistic regression approach (with sex, age, height and weight as covariates) and detected significant associations between specific embeddings and the following conditions: non-insulin-dependent diabetes, epilepsy, glaucoma and chronic ischaemic heart disease (**Fig.4B**). Two of these lead signals (epilespy and chronic ischaemic heart disease) are associated very specifically to only one embedding each (embedding no.1 and no.26 respectively). In contrast, glaucoma is associated with two different embeddings (no.39 and no.47) and diabetes to three sequential embeddings (no.36-38) (**Fig.4C**). Reassuringly, GWAS analysis of embeddings no.36-38 revealed statistically significant signals linked to *ADCY5* (**Supplementary Table 1**), a gene that influences glucose metabolism and has been previously linked to non-insulin-dependent diabetes by multiple association studies (Roman 2017).

To understand which aspects of retinal morphology drove the association between the embedded features and the lead disease codes (non-insulin-dependent diabetes, epilepsy, glaucoma and chronic ischaemic heart disease) we inspected a set of retinal thickness difference maps. These compared retinal thickness in UK Biobank participants that had been assigned the relevant ICD10 code (after OCT imaging) to those that have not (**Fig.4D**). In keeping with previous observations: (i) the main areas of difference for diabetes were the paracentral region and the areas temporal to the optic disc (corresponding to the major retinal vessels) (Li 2021); (ii) the main area of difference for glaucoma corresponded to what is described in the glaucoma literature as the “macular vulnerability zone” (Hood 2017).

### Using autoencoder-derived OCT phenotypes to gain insights into disease risk

We investigated if autoencoder-derived embedded features from an individual’s OCT scan can help predict the occurrence of certain diseases, including glaucoma and cardiovascular disorders. We used survival analysis (Cox proportional hazard regression) and found significant links between specific embeddings and the occurrence of disease (after the OCT scan date) (**Fig.5A**). High risk cohorts identified based on the embedded features showed a higher chance of being affected by glaucoma or cardiovascular conditions compared to the sex-stratified baseline rate of disease occurrence. In other words, the embedded features could help identify high-risk cohorts (**Fig.5B**). It is highlighted that a few embeddings appear to be linked to multiple diseases (e.g. no.28), while others have no effect on any disease or are specific to single disease codes (e.g. embedding no.18 for chronic ischaemic heart disease). A notable observation is the link between multiple embeddings and essential hypertension. This is often in the presence of signals from other cardiovascular disease codes suggesting that changes in blood pressure can lead to alterations in OCT-evaluated retinal structure which may in turn be a marker for the development of cardiovascular complications (**Fig.5C**).

**Figure 5.**
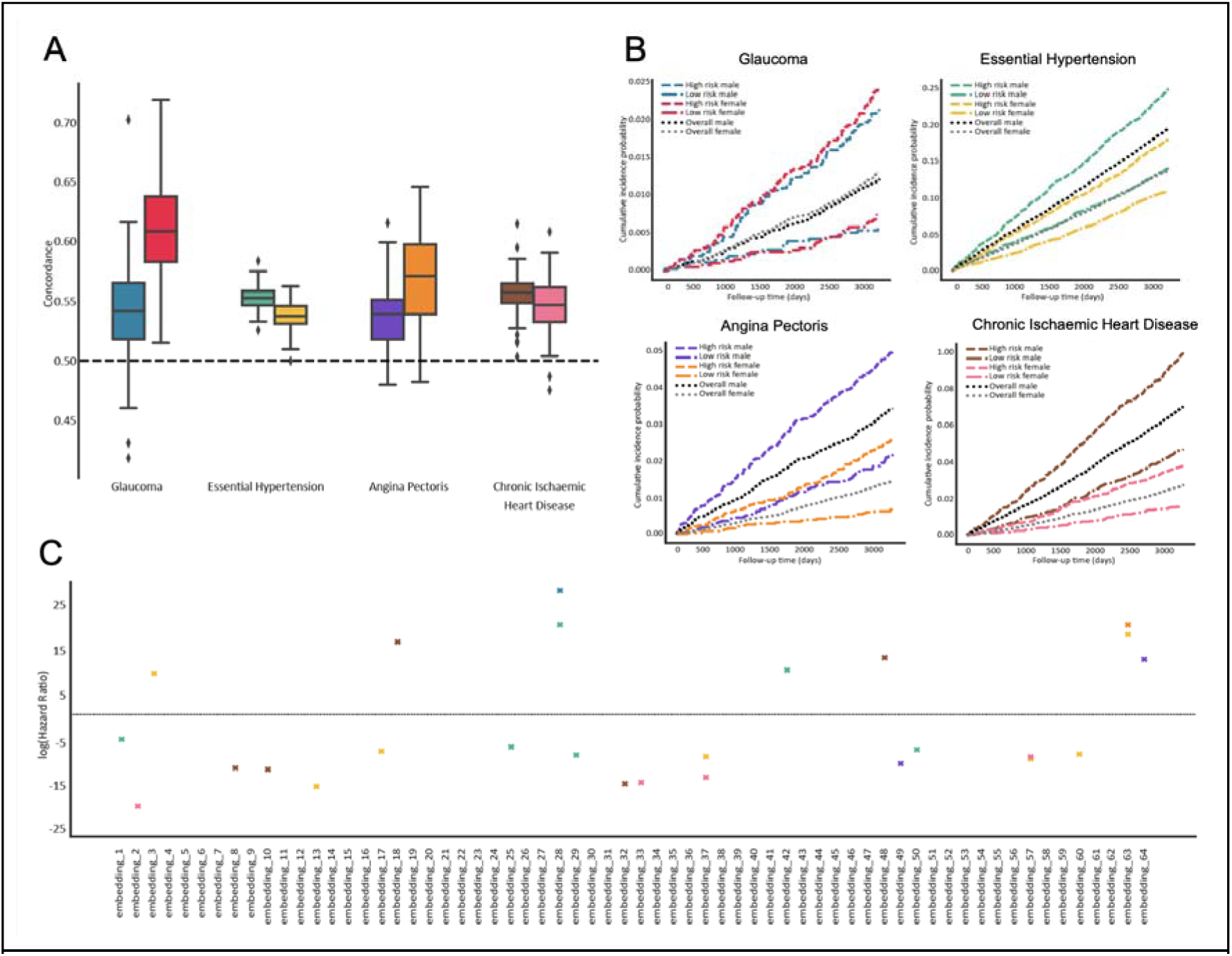
Survival analysis investigating the contribution of embedded features upon the time-to-diagnosis for four ICD10 disease codes. [A] Concordance index evaluating the embedding-incorporating model’s ability to discriminate sex-stratified disease occurrence; the distribution across 20 repetitions of five-fold cross-validation are shown (n = 100 for each box plot); all box plots demarcate quartiles and median values, while whiskers extend to 1.5x the interquartile range. [B] Kaplan-Meier plots showing sex-stratified risk of disease occurrence for the overall population as well as for high-risk cohorts determined by the embedding-incorporating model (top 25% based on Cox regression). [C] Graph highlighting which embedded features have a significant relationship with the selected diseases in male and female cohorts; −log10 hazard ratios are shown.

## DISCUSSION

Phenotypes are abstract entities that can be thought of as simplified maps carved from higher dimension spaces (Cortese 2021). These maps are generally influenced by a combination of genetic, environmental and stochastic factors. Discovering phenotypes that represent distinct biological pathways and/or have pragmatic medical significance is of particular interest (Dahl & Zaitlen 2020). Here, we show that a computational, autoencoder-based approach can be used to efficiently extract informative phenotypes from retinal OCT images.

Analysis of the genetic basis of autoencoder-derived embedded features revealed 118 statistically significant (p-value < 3.2 x 10^-10^) association signals. Notably, three recent studies that used a similar analytical approach but focused on different imaging modalities — fundus photography (Kirchler 2022; Xie 2023) and cardiac magnetic resonance images (Bonazzola 2023) — identified a slightly smaller number of genetic associations (Supplementary Table 2). Whilst most of the loci detected here have prior links to retinal phenotypes, a subset of them have no such prior associations. One example is the locus around *LPHN2/ADGRL2*, a gene encoding a synaptic adhesion molecule implicated in guiding neural circuit connectivity (Donahue 2021) (lead marker: rs1492258; association with 7 autoencoder-derived embedded features; minimum p-value 1.4 x 10^-15^). Although this gene is expressed in the retina, especially in the bipolar cells (Karlsson 2021), it has not been previously associated with a retinal phenotype.

Reassuringly, there was a significant overlap between the findings of the present study and the results of previous analyses that investigated the genetic architecture of traditional OCT-derived retinal phenotypes. These include three UK Biobank studies: (i) one that looked at macular (*i.e* total central retinal) thickness and reported 139 loci (Gao 2019), and (ii) two from our group that investigated OCT-derived measurements of inner and outer retinal layers, and reported 46 and 111 loci respectively (Currant 2021; Currant 2023). Overall, 36% (98/273) of the combined lead loci from these studies also reached genome-wide significance in the present analysis (58%, 33% and 41% for Currant 2021, Currant 2023 and Gao 2019 respectively). Interestingly, the two signals with the highest statistical significance in the macular thickness GWAS conducted by Gao and colleagues (Gao 2019) were also the most significant hits in this study (**Fig.2**). The marker with the highest statistical significance was within the *LINC00461* locus. *LINC00461* is a long noncoding RNA that is the primary transcript of miR-9-2. *LINC00461* is highly expressed in neural stem cells and a decrease in its expression has been shown to alter the timing of retinal neurogenesis (Thomas 2022). The locus with the second highest statistical significance encompassed the *TSPAN10* gene.

In the eye, *TSPAN10* is predominantly expressed in melanin-containing cells (retinal pigment epithelia [RPE] and uveal melanocytes), and the corresponding protein is thought to have a role in regulating retinal cell fate and development (Dornier 2012; Haining 2012; Orozco 2020). Further functional genomic analyses of these two key loci are expected to provide important insights into developmental processes shaping human retinal morphology.

An intriguing association that we detected was that between certain autoencoder-derived retinal phenotypes and a common 17q21.31 inversion encompassing the *MAPT* gene. *MAPT* is primarily expressed in brain neurons, and genetic alterations impacting the *MAPT* locus have been linked to several neurodegenerative disorders including Alzheimer disease, frontotemporal dementia and parkinsonism (Wang 2016; Shi 2021). Recently, inner retinal layer thickness parameters and glaucoma have been added to the growing list of phenotypes associated with the *MAPT* locus (Gharahkhani 2021; Diaz-Torres 2023). Further work is required to pinpoint which (and how many) genes within the *MAPT* region are causally associated with retinal and brain phenotypes (Diaz-Torres 2023). More broadly, the extent to which the overlap between neurodegenerative disorders, retinal morphology and glaucoma reflects pleiotropy rather than causal relationships remains to be determined. Of note, causal genetic effects in both directions have been previously suggested between retinal imaging traits and Alzheimer disease (Zhao 2023) while little support has been found for a causal relationship between glaucoma and Alzheimer disease (Budu-Aggrey 2020).

Deep learning approaches have been shown to be able to detect imaging patterns that are not amenable to human identification and which can assist with prediction tasks (Radhakrishnan 2023). For example, neural networks can predict sex and age with good accuracy from retinal OCT images (Chueh 2022; Le Goallec 2022) whereas human experts find these tasks impossible. Here, we investigated if autoencoders can identify OCT parameters that can be used to predict health outcomes (glaucoma and cardiovascular disease). Although the overall predictive ability of the generated models was moderate, the autoencoder-derived features were shown to enhance risk stratification. These observations suggest that it is not inconceivable that purpose-built autoencoders will play a role in improving the efficiency of medical screening programs in the future.

This study has a number of limitations. First, the autoencoder input was retinal thickness maps generated using a U-Net approach which made our framework semi-automated (as a small amount of manual labelling was required). Using three-dimensional autoencoders to extract features directly from OCT volume scans could fully automate the pipeline, minimising any subjective aspects and reducing the burden of data curation (Diaz-Pinto 2022). Second, we only performed common-variant genetic association analyses of the obtained embedded features. The increasing availability of genome sequencing data in UK Biobank participants will allow us to more comprehensively look for genetic associations, including with rare variants and with copy number alterations in the future. Third, the fact that relationships were detected between embeddings and certain health outcomes does not necessarily imply causation. The main aim of this study was to assess if autoencoders can be utilised to produce biologically and clinically relevant phenotypes. In-depth confounder adjustment and causal inference studies were therefore not performed. Furthermore, the predictive models described here have a proof-of-concept nature and are not intended for implementation (especially as the data used for training and evaluation were highly homogeneous).

In summary, this study proposes a framework for retinal phenotyping based on a self-supervised deep learning approach. Our findings highlight that autoencoder-based techniques can be used to extract knowledge about the genetic factors determining retinal morphology. The outlined approach is flexible and can be adapted and extended to other organs and imaging modalities.

## ONLINE METHODS

### Cohort characteristics

The UK Biobank is a biomedical resource containing in-depth genetic and health information from >500,000 individuals from across the UK. Participants were recruited between 2006 and 2010 and, at enrolment, were between 40 and 69 years of age. At the initial assessment, UK Biobank volunteers provided consent, answered questions on socio-demographic, lifestyle and health-related factors, completed a range of physical measures, and provided biological samples. DNA was extracted from the donated blood samples and was used to generate genotyping array data. The baseline information has been extended in several ways. For example, repeat assessments were conducted in subsets of the cohort every few years (Bycroft 2018). Notably, thousands of UK Biobank participants underwent ophthalmic phenotyping including imaging of the central retina using OCT (>84,000 individuals) (Patel 2016; Chua 2019). A total of 67,664 individuals were imaged at the time of their baseline visit (Instance 0, “Initial assessment visit (2006-2010)”); this cohort was the focus of the primary analysis. A further 17,090 different participants were imaged for the first time during their first repeat assessment (Instance 1, “First repeat assessment visit (2012-13)”); these were included in the replication study.

We performed quality control considering genetic and imaging parameters. First, to reduce the impact of population stratification and to increase the validity of the conducted genetic association studies, we focused on individuals within a genetically well-mixed, European-like subset of the UK Biobank. This was achieved by applying PCA to UK Biobank genotypic data using standard, previously-implemented methods (Currant 2023). Additional participants were excluded as their OCT scans failed to meet a set of previously-described, rigorous quality control criteria (Patel 2016; Currant 2021; Currant 2023). Finally, participants were removed on the basis of being recommended for exclusion from genetic studies by the UK Biobank or because they were related to third degree or more. The final dataset for the primary analysis included 31,135 study subjects (**Supplementary** Fig.1). Similar criteria were used for the replication study with the exception of the imaging quality control parameters which were identical to those described by Zekavat and colleagues (Zekavat 2023).

### Generation of thickness maps from OCT volume scans

All the UK Biobank volunteers that were analysed as part of this study were imaged using the 3D OCT-1000 Mark II device (Topcon, Japan). OCT imaging was carried out in a dark room without pupil dilation using the 3D 6×6 mm^2^ macular volume scan mode (128 horizontal B-scans in a 6×6 mm raster pattern). The right eye was imaged first (Patel 2016; Chua 2019). Our analysis focused on left eye images as we assumed that familiarity with the test would have led to scans that, on average, had higher overall quality. A total of 128 PNG images were generated from each tested eye with the dimensions of each PNG image being 650 x 512 x 1 grayscale pixels. After cropping the top (superior) and bottom (inferior) edge of the image area, PNG images with dimensions of 512 x 512 x 1 pixels were obtained.

The 128 images of each OCT scan were used to create a “thickness map”, *i.e.* a single image displaying the retinal thickness throughout the imaged area. To achieve this, segmentation of all the scans in the dataset was performed using a U-Net based approach. The utilised U-Net method was first described in 2015 (Ronneberger 2015) and involves a fully convolutional network that consists of a contracting path (that extracts features) and an expansive path (that localises objects).

Initially, the inner- and outer-most limits of the retina (corresponding to the inner limiting membrane and the Bruch’s membrane respectively) were manually identified in 100 randomly-selected OCT images using the https://www.makesense.ai tool. The original images and the generated “ground truth” segmentation masks were subsequently utilised to train the U-Net. Adaptive Moment Estimation (Adam) was used to optimize the algorithm for training the network parameters and training was performed for 50 epochs. The output of the U-Net consisted of segmented OCT images (analogous to the provided masks). These were used to calculate retinal thickness (*i.e.* the vertical distance between the top and bottom edge of the mask in each of the 512 points of the horizontal axis). The obtained measurements were compared to those acquired through the purpose-built Topcon Advanced Boundary Segmentation (TABS) software (the latter are available in the UK Biobank dataset). Good correlation was observed in retinae both with and without pathology, increasing confidence in the utilised approach (**Supplementary** Fig.6). Finally, the thickness measurements from the 128 images (‘slices’) that were obtained in each tested left eye were combined and used to generate a thickness map for each UK Biobank participant that met the inclusion criteria of this study (**Fig.1**).

### Autoencoder set-up

An autoencoder was used for self-supervised feature extraction from the 31,135 left eye OCT-derived thickness maps. A conventional autoencoder architecture was utilised (Hinton 2006; Michelucci 2022): the encoder network projected the input images to a low-dimensional space (‘latent space’) with 64 variables (‘embeddings’), and a function was used to try to reconstruct the original images from these 64 latent space representations. A Mean Squared Error (MSE) loss function was employed to measure the deviation between reconstructed and input data (but otherwise the reconstructed images were not used in the primary analysis). It is noted that the autoencoder was trained end-to-end for 150 epochs utilising 2500 training and 500 test images. We trialled different autoencoder layouts with bottleneck layers of the following sizes: 128, 64, 32 and 16. For 128 and 64, we obtained very similar reconstruction loss curves during training over 300 epochs. In contrast, for both 32 and 16 the image reconstruction loss could not be dropped below 0.006 suggesting that these models were unable to generalise as well as the larger bottleneck sizes. We then selected a bottleneck size of 64 since this was the smallest size with the best image reconstruction accuracy among the layouts that we tested.

To extract further information from the latent space, PCA (*i.e.* linear dimensionality reduction) was performed using the 64 embeddings as input; the first 25 principal components were then considered for further analyses.

### Genome-wide association studies: primary analysis

GWAS analyses of autoencoder-derived embedded features (64 embeddings and 25 embedding-related principal components) were performed using an additive linear model implemented in REGENIE v3.1.1 (https://rgcgithub.github.io/regenie/) (Mbatchou 2021). All embedded variables were inverse rank normalised prior to modelling with REGENIE to avoid any potential bias that could be introduced by outlier values. The following quality-control filters were applied on the imputed genotype data (UK Biobank data-field 22828) during the creation of the whole-genome regression model (REGENIE step 1): a minor allele frequency (MAF)_J≥_J5%; Hardy–Weinberg equilibrium test not exceeding *P*_J>_J1 × 10^−15^; a genotyping rate above 99%; not present in a low-complexity region, a region of long-range linkage disequilibrium or a sex chromosome (Mbatchou 2021). This resulted in up to 7,114,193 genotyped variants that were tested for association using a Firth logistic regression model (REGENIE step 2). Correction for the following covariates was undertaken: age at recruitment (data-field 21022), sex (data-field 31), height (data-field 50), weight (data-field 21002), refractive error (calculated as spherical error + 0.5 × cylindrical error; data-fields 5085 and 5086) and genetic principal components 1 to 20 (data-field 22009). Two levels of statistical significance were used: the conventional genome-wide significance (p-value < 5 × 10^−8^) and a conservative, “study-wide” threshold (p-value < 3.2 x 10^-10^ following Bonferroni correction for 153 tests).

A degree of correlation was expected among autoencoder-derived embeddings and the summary statistics obtained from the GWAS analyses were used to perform a multi-trait meta-analysis. First, embeddings with a high genetic correlation (*i.e.* with Pearson correlation coefficient R > 0.9) were identified. Then, the MTAG v1.0.8 tool (https://github.com/JonJala/mtag) (Turley 2018) was used to conduct a single meta-analysis for every individual inverse rank normalised embedding, leveraging the findings from correlated embedded features and producing an updated set of GWAS summary statistics for each of these 64 variables. Under certain assumptions, the generated estimates will be more precise than those obtained from the input GWAS (Turley 2018).

To refine the obtained association signals, further analyses were performed using the GCTA-COJO tool (https://yanglab.westlake.edu.cn/software/gcta/#COJO) (Yang 2010). These analyses were conducted utilising linkage disequilibrium estimates from a reference sample (Currant 2023) and summary statistics from: (i) the 64 embedding GWAS, (ii) the 25 embedding-related principal component GWAS, (iii) the 64 embedding MTAG-GWAS. Genetic variants in loci that were on different chromosomes or more than 10 Mb distant from each other were assumed to be uncorrelated.

Genetic changes in the main variant set were annotated using Ensembl (Cunningham 2022), Open Targets (Ochoa 2021) and GWAS Catalog (Sollis 2023) data. To accurately summarise the strongest signals (presented in **Table 2**), the linkage disequilibrium metrics of the changes that were highlighted as lead variants by GCTA-COJO analysis and were within 1 Mb of one another were manually inspected using the LDlink tool (Myers 2020).

### Genome-wide association studies: replication

We sought to replicate the genetic associations detected in the primary analysis in a different set of OCT images. As the number of open resources that have sufficiently large human cohorts with combined genomic and OCT imaging data is very small, we made use of the UK Biobank “Instance 1” left eye scans (data-field: 21017_1_0). This included images from 17,090 participants that were not part of the discovery/primary cohort and were not used for training either the U-net segmentation or the autoencoder. It is noted that these additional OCT images were obtained at a different time (2012-13) compared to the scans in the discovery/primary cohort (2006-2010). Due to the inconsistent capture of certain OCT-related metrics in the replication cohort scans, we used a different set of image QC exclusion criteria. Following the removal of poor quality and outlier images (using the approach described in Zekavat 2023), the replication cohort included 10,439 high quality scans from unrelated UK Biobank participants of predominantly European-like genetic ancestries (as determined by PCA of genotypes). A replication GWAS was then performed using exactly the same parameters as in the discovery/primary study (outlined above). To gain insights into the extent to which the findings of the primary and the replication study were in agreement, we assessed the degree of correlation between the detected effect size estimates; the relevant beta-beta plots are shown in **Supplementary** Fig.4.

### Correlation and logistic regression analyses

Direct pairwise comparisons between the 64 embeddings were performed and the relevant Pearson correlation coefficients (R) were calculated. Genetic correlation was also estimated, again using Pearson correlation coefficients but this time utilising the effect size estimates from across the significant associations for all 64 embeddings. The two correlation matrices that were generated were then displayed using a heatmap where rows and columns were ordered by the distances obtained via hierarchical clustering (on the embedding value correlation matrix only) (**Fig.4**).

In addition to evaluating the relationship between pairs of the studied embedded features, correlation analyses were performed to look for links between each of these 64 features and four ophthalmic traits (**Supplementary** Fig.3). Furthermore, a logistic regression approach was used to look for relationships between embeddings and a set of diseases (high-level ICD10 codes); only the 454 disease-related codes for which there were >1000 cases in the UK Biobank cohort were considered (when factoring in only data obtained after the date of OCT image acquisition (2012)). Age, sex, height and weight were used as covariates and the statistical significance threshold was determined using Bonferroni correction.

### Predictive modelling

Survival analysis was performed using penalized Cox proportional hazard regression; a mixture of L1 and L2 regularisation was utilised (often referred to as the Cox elastic net). We focused on two main outcomes – glaucoma and cardiovascular disorders (essential hypertension, angina pectoris and chronic ischaemic heart disease). These included ICD10 codes that were highlighted as significant by the logistic regression analyses described in the previous section, and were chosen as predicting them was deemed to be of clinical significance. Only diagnoses assigned after the date of OCT image acquisition were considered. To evaluate discriminative performance, we used Harrell’s C-index as a measure of the concordance between predicted and actual risk. The hyperparameter of L1/L2 penalization strength was set to 0.1 and 20 repetitions of five-fold cross-validation were used to evaluate model performance. Survival curves were estimated using the Kaplan-Meier estimator.

### Ethics approval

The UK Biobank has received approval from the National Information Governance Board for Health and Social Care and the National Health Service North West Centre for Research Ethics Committee (Ref: 11/NW/0382). This research was conducted using the UK Biobank Resource under projects 49978, 53144 and 2112. All investigations were conducted in accordance with the tenets of the Declaration of Helsinki.

## DATA AVAILABILITY

UK Biobank data are available under restricted access through a procedure described at http://www.ukbiobank.ac.uk/using-the-resource/. All other data supporting the findings of this study are available within the article (including its supplementary information files).

## CODE AVAILABILITY

The scripts used to analyse the datasets included in this study are available at https://github.com/tf2/autoencoder-oct.

## ACKNOWLEDGEMENTS

We acknowledge the following sources of funding: the Wellcome Trust (224643/Z/21/Z, Clinical Research Career Development Fellowship to P.I.S.); the UK National Institute for Health Research (NIHR) Clinical Lecturer Programme (CL-2017-06-001 to P.I.S.); the EMBL European Bioinformatics Institute (EMBL-EBI) (A.D., K.G., E.B., T.F.). This research was co-funded by the NIHR Manchester Biomedical Research Centre (NIHR203308). The views expressed are those of the author(s) and not necessarily those of the NIHR or the Department of Health and Social Care.

We also acknowledge the contribution of the UK Biobank Eye and Vision Consortium. Members of this consortium include: Naomi Allen, Tariq Aslam, Denize Atan, Sarah Barman, Jenny Barrett, Paul Bishop, Graeme Black, Tasanee Braithwaite, Roxana Carare, Usha Chakravarthy, Michelle Chan, Sharon Chua, Alexander Day, Parul Desai, Bal Dhillon, Andrew Dick, Alexander Doney, Cathy Egan, Sarah Ennis, Paul Foster, Marcus Fruttiger, John Gallacher, David Garway-Heath, Jane Gibson, Jeremy Guggenheim, Chris Hammond, Alison Hardcastle, Simon Harding, Ruth Hogg, Pirro Hysi, Pearse Keane, Peng Tee Khaw, Anthony Khawaja, Gerassimos Lascaratos, Thomas Littlejohns, Andrew Lotery, Robert Luben, Phil Luthert, Tom Macgillivray, Sarah Mackie, Savita Madhusudhan, Bernadette Mcguinness, Gareth Mckay, Martin Mckibbin, Tony Moore, James Morgan, Eoin O’Sullivan, Richard Oram, Chris Owen, Praveen Patel, Euan Paterson, Tunde Peto, Axel Petzold, Nikolas Pontikos, Jugnoo Rahi, Alicja Rudnicka, Naveed Sattar, Jay Self, Panagiotis Sergouniotis, Sobha Sivaprasad, David Steel, Irene Stratton, Nicholas Strouthidis, Cathie Sudlow, Zihan Sun, Robyn Tapp, Dhanes Thomas, Emanuele Trucco, Adnan Tufail, Ananth Viswanathan, Veronique Vitart, Mike Weedon, Cathy Williams, Katie Williams, Jayne Woodside, Max Yates, Jennifer Yip, Yalin Zheng.

The UK Biobank Eye and Vision Consortium is supported by funding from the NIHR Biomedical Research Centre at Moorfields Eye Hospital and UCL Institute of Ophthalmology, the Alcon Foundation and the Desmond Foundation.

## AUTHOR CONTRIBUTIONS STATEMENT

P.I.S., E.B. and T.F. conceived and designed the experiments. P.I.S., A.D., K.G., E.B. and T.F. analysed the data. P.I.S. and T.F. wrote the manuscript with support from E.B.. All authors critically revised and approved the manuscript.

## REFERENCES

Bonazzola R, Ferrante E, Ravikumar N, Xia Y, Keavney B, Plein S, Syeda-Mahmood T, Frangi AF. Unsupervised ensemble-based phenotyping helps enhance the discoverability of genes related to heart morphology. arXiv 2023.doi: 10.48550/arXiv.2301.02916.

Bouma BE, de Boer JF, Huang D, Jang IK, Yonetsu T, Leggett CL, Leitgeb R, Sampson DD, Suter M, Vakoc B, Villiger M, Wojtkowski M. Optical coherence tomography. Nat Rev Methods Primers. 2022;2:79. doi: 10.1038/s43586-022-00162-2.

Bulik-Sullivan BK, Loh PR, Finucane HK, Ripke S, Yang J; Schizophrenia Working Group of the Psychiatric Genomics Consortium; Patterson N, Daly MJ, Price AL, Neale BM. LD Score regression distinguishes confounding from polygenicity in genome-wide association studies. Nat Genet. 2015;47(3):291–5. doi: 10.1038/ng.3211.

Budu-Aggrey A, Hysi P, Kehoe PG, Igo RP, Wiggs JL, Cooke Bailey J, Haines J, Pasquale LR, MacGregor S, NEIGHBORHOOD consortium, International Glaucoma Genetics Consortium, UK Biobank, Davey Smith G, Davies NM, Atan D. The relationship between open angle glaucoma, optic disc morphology and Alzheimer’s Disease: a Mendelian randomization study. bioRxiv. 2020.doi: 10.1101/2020.08.30.20184846.

Bycroft C, Freeman C, Petkova D, Band G, Elliott LT, Sharp K, Motyer A, Vukcevic D, Delaneau O, O’Connell J, Cortes A, Welsh S, Young A, Effingham M, McVean G, Leslie S, Allen N, Donnelly P, Marchini J. The UK Biobank resource with deep phenotyping and genomic data. Nature. 2018;562(7726):203–209. doi: 10.1038/s41586-018-0579-z.

Chua SYL, Thomas D, Allen N, Lotery A, Desai P, Patel P, Muthy Z, Sudlow C, Peto T, Khaw PT, Foster PJ; UK Biobank Eye & Vision Consortium. Cohort profile: design and methods in the eye and vision consortium of UK Biobank. BMJ Open. 2019;9(2):e025077. doi: 10.1136/bmjopen-2018-025077.

Cortese A, Yamamoto A, Hashemzadeh M, Sepulveda P, Kawato M, De Martino B. Value signals guide abstraction during learning. Elife. 2021;10:e68943. doi: 10.7554/eLife.68943.

Cunningham F, Allen JE, Allen J, Alvarez-Jarreta J, Amode MR, Armean IM, Austine-Orimoloye O, Azov AG, Barnes I, Bennett R, Berry A, Bhai J, Bignell A, Billis K, Boddu S, Brooks L, Charkhchi M, Cummins C, Da Rin Fioretto L, Davidson C, Dodiya K, Donaldson S, El Houdaigui B, El Naboulsi T, Fatima R, Giron CG, Genez T, Martinez JG, Guijarro-Clarke C, Gymer A, Hardy M, Hollis Z, Hourlier T, Hunt T, Juettemann T, Kaikala V, Kay M, Lavidas I, Le T, Lemos D, Marugán JC, Mohanan S, Mushtaq A, Naven M, Ogeh DN, Parker A, Parton A, Perry M, Piližota I, Prosovetskaia I, Sakthivel MP, Salam AIA, Schmitt BM, Schuilenburg H, Sheppard D, Pérez-Silva JG, Stark W, Steed E, Sutinen K, Sukumaran R, Sumathipala D, Suner MM, Szpak M, Thormann A, Tricomi FF, Urbina-Gómez D, Veidenberg A, Walsh TA, Walts B, Willhoft N, Winterbottom A, Wass E, Chakiachvili M, Flint B, Frankish A, Giorgetti S, Haggerty L, Hunt SE, IIsley GR, Loveland JE, Martin FJ, Moore B, Mudge JM, Muffato M, Perry E, Ruffier M, Tate J, Thybert D, Trevanion SJ, Dyer S, Harrison PW, Howe KL, Yates AD, Zerbino DR, Flicek P. Ensembl 2022. Nucleic Acids Res. 2022;50(D1):D988–D995. doi: 10.1093/nar/gkab1049

Currant H, Hysi P, Fitzgerald TW, Gharahkhani P, Bonnemaijer PWM, Senabouth A, Hewitt AW; UK Biobank Eye and Vision Consortium; International Glaucoma Genetics Consortium; Atan D, Aung T, Charng J, Choquet H, Craig J, Khaw PT, Klaver CCW, Kubo M, Ong JS, Pasquale LR, Reisman CA, Daniszewski M, Powell JE, Pébay A, Simcoe MJ, Thiadens AAHJ, van Duijn CM, Yazar S, Jorgenson E, MacGregor S, Hammond CJ, Mackey DA, Wiggs JL, Foster PJ, Patel PJ, Birney E, Khawaja AP. Genetic variation affects morphological retinal phenotypes extracted from UK Biobank optical coherence tomography images. PLoS Genet. 2021;17(5):e1009497. doi: 10.1371/journal.pgen.1009497.

Currant H, Fitzgerald TW, Patel PJ, Khawaja AP; UK Biobank Eye and Vision Consortium; Webster AR, Mahroo OA, Birney E. Sub-cellular level resolution of common genetic variation in the photoreceptor layer identifies continuum between rare disease and common variation. PLoS Genet. 2023;19(2):e1010587. doi: 10.1371/journal.pgen.1010587.

Chueh KM, Hsieh YT, Chen HH, Ma IH, Huang SL. Identification of sex and age from macular optical coherence tomography and feature analysis using deep learning. Am J Ophthalmol. 2022;235:221–228. doi: 10.1016/j.ajo.2021.09.015.

Dahl A, Zaitlen N. Genetic influences on disease subtypes. Annu Rev Genomics Hum Genet. 2020;21:413–435. doi: 10.1146/annurev-genom-120319-095026.

De Fauw J, Ledsam JR, Romera-Paredes B, Nikolov S, Tomasev N, Blackwell S, Askham H, Glorot X, O’Donoghue B, Visentin D, van den Driessche G, Lakshminarayanan B, Meyer C, Mackinder F, Bouton S, Ayoub K, Chopra R, King D, Karthikesalingam A, Hughes CO, Raine R, Hughes J, Sim DA, Egan C, Tufail A, Montgomery H, Hassabis D, Rees G, Back T, Khaw PT, Suleyman M, Cornebise J, Keane PA, Ronneberger O. Clinically applicable deep learning for diagnosis and referral in retinal disease. Nat Med. 2018;24(9):1342–1350. doi: 10.1038/s41591-018-0107-6.

Diaz-Pinto A, Ravikumar N, Attar R, Suinesiaputra A, Zhao Y, Levelt E, Dall’Armellina E, Lorenzi M, Chen Q, Keenan TDL, Agron E, Chew EY, Lu Z, Gale CP, Gale RP, Plein S, Frangi AF. Predicting myocardial infarction through retinal scans and minimal personal information. Nat Mach Intell 2022;4:55–61. doi: 10.1038/s42256-021-00427-7.

Diaz-Torres S, He W, Thorp J, Seddighi S, Mullany S; IGGC International Glaucoma Genetics Consortium; Hammond CJ, Hysi PG, Pasquale LR, Khawaja AP, Hewitt AW, Craig JE, Mackey DA, Wiggs JL, van Duijn C, Lupton MK, Ong JS, MacGregor S, Gharahkhani P. Disentangling the genetic overlap and causal relationships between primary open-angle glaucoma, brain morphology and four major neurodegenerative disorders. EBioMedicine. 2023;92:104615. doi: 10.1016/j.ebiom.2023.104615.

Donohue JD, Amidon RF, Murphy TR, Wong AJ, Liu ED, Saab L, King AJ, Pae H, Ajayi MT, Anderson GR. Parahippocampal latrophilin-2 (ADGRL2) expression controls topographical presubiculum to entorhinal cortex circuit connectivity. Cell Rep. 2021;37(8):110031. doi: 10.1016/j.celrep.2021.110031.

Dornier E, Coumailleau F, Ottavi JF, Moretti J, Boucheix C, Mauduit P, Schweisguth F, Rubinstein E. TspanC8 tetraspanins regulate ADAM10/Kuzbanian trafficking and promote Notch activation in flies and mammals. J Cell Biol. 2012;199(3):481–96. doi: 10.1083/jcb.201201133.

Elliott LT, Sharp K, Alfaro-Almagro F, Shi S, Miller KL, Douaud G, Marchini J, Smith SM. Genome-wide association studies of brain imaging phenotypes in UK Biobank. Nature. 2018;562(7726):210–216. doi: 10.1038/s41586-018-0571-7.

Espinosa I, Alfonso-Sánchez MA, Gómez-Pérez L, Peña JA. Neolithic expansion and the 17q21.31 inversion in Iberia: an evolutionary approach to H2 haplotype distribution in the Near East and Europe. Mol Genet Genomics. 2023;298(1):153–160. doi: 10.1007/s00438-022-01969-0.

Gao XR, Huang H, Kim H. Genome-wide association analyses identify 139 loci associated with macular thickness in the UK Biobank cohort. Hum Mol Genet. 2019;28(7):1162–1172. doi: 10.1093/hmg/ddy422.

Gharahkhani P, Jorgenson E, Hysi P, Khawaja AP, Pendergrass S, Han X, Ong JS, Hewitt AW, Segrè AV, Rouhana JM, Hamel AR, Igo RP Jr, Choquet H, Qassim A, Josyula NS, Cooke Bailey JN, Bonnemaijer PWM, Iglesias A, Siggs OM, Young TL, Vitart V, Thiadens AAHJ, Karjalainen J, Uebe S, Melles RB, Nair KS, Luben R, Simcoe M, Amersinghe N, Cree AJ, Hohn R, Poplawski A, Chen LJ, Rong SS, Aung T, Vithana EN; NEIGHBORHOOD consortium; ANZRAG consortium; Biobank Japan project; FinnGen study; UK Biobank Eye and Vision Consortium; GIGA study group; 23 and Me Research Team; Tamiya G, Shiga Y, Yamamoto M, Nakazawa T, Currant H, Birney E, Wang X, Auton A, Lupton MK, Martin NG, Ashaye A, Olawoye O, Williams SE, Akafo S, Ramsay M, Hashimoto K, Kamatani Y, Akiyama M, Momozawa Y, Foster PJ, Khaw PT, Morgan JE, Strouthidis NG, Kraft P, Kang JH, Pang CP, Pasutto F, Mitchell P, Lotery AJ, Palotie A, van Duijn C, Haines JL, Hammond C, Pasquale LR, Klaver CCW, Hauser M, Khor CC, Mackey DA, Kubo M, Cheng CY, Craig JE, MacGregor S, Wiggs JL. Genome-wide meta-analysis identifies 127 open-angle glaucoma loci with consistent effect across ancestries. Nat Commun. 2021;12(1):1258. doi: 10.1038/s41467-020-20851-4.

Gong W, Bai S, Zheng YQ, Smith SM, Beckmann CF. Supervised phenotype discovery from multimodal brain imaging. IEEE Trans Med Imaging. 2022;PP. doi: 10.1109/TMI.2022.3218720.

Haining EJ, Yang J, Bailey RL, Khan K, Collier R, Tsai S, Watson SP, Frampton J, Garcia P, Tomlinson MG. The TspanC8 subgroup of tetraspanins interacts with A disintegrin and metalloprotease 10 (ADAM10) and regulates its maturation and cell surface expression. J Biol Chem. 2012;287(47):39753–65. doi: 10.1074/jbc.M112.416503.

Hasson U, Nastase SA, Goldstein A. Direct Fit to Nature: An evolutionary perspective on biological and artificial neural networks. Neuron. 2020;105(3):416–434. doi: 10.1016/j.neuron.2019.12.002.

Hinton GE, Salakhutdinov RR. Reducing the dimensionality of data with neural networks. Science. 2006;313(5786):504-7. doi: 10.1126/science.1127647.

Hinton G. Deep learning-A technology with the potential to transform health care. JAMA. 2018;320(11):1101–1102. doi: 10.1001/jama.2018.11100.

Hood DC. Improving our understanding, and detection, of glaucomatous damage: An approach based upon optical coherence tomography (OCT). Prog Retin Eye Res. 2017;57:46–75. doi: 10.1016/j.preteyeres.2016.12.002.

Karlsson M, Zhang C, Méar L, Zhong W, Digre A, Katona B, Sjöstedt E, Butler L, Odeberg J, Dusart P, Edfors F, Oksvold P, von Feilitzen K, Zwahlen M, Arif M, Altay O, Li X, Ozcan M, Mardinoglu A, Fagerberg L, Mulder J, Luo Y, Ponten F, Uhlén M, Lindskog C. A single-cell type transcriptomics map of human tissues. Sci Adv. 2021;7(31):eabh2169. doi: 10.1126/sciadv.abh2169.

Keenan TDL, Clemons TE, Domalpally A, Elman MJ, Havilio M, Agrón E, Benyamini G, Chew EY. Retinal specialist versus artificial intelligence detection of retinal fluid from OCT: Age-Related Eye Disease Study 2: 10-year follow-on study. Ophthalmology. 2021;128(1):100–109. doi: 10.1016/j.ophtha.2020.06.038.

Kirchler M, Konigorski S, Norden M, Meltendorf C, Kloft M, Schurmann C, Lippert C. transferGWAS: GWAS of images using deep transfer learning. Bioinformatics. 2022;38(14):3621–3628. doi: 10.1093/bioinformatics/btac369.

LeCun Y, Bengio Y, Hinton G. Deep learning. Nature. 2015;521(7553):436–44. doi: 10.1038/nature14539.

Le Goallec A, Dai Y, Collin S, Vincent T, Patel CJ. Deep learning of fundus and optical coherence tomography images enables identification of diverse genetic and environmental factors associated with eye aging. bioRxiv. 2022. doi: 10.1101/2021.06.24.21259471.

Li H, Yu X, Zheng B, Ding S, Mu Z, Guo L. BMC Ophthalmol. 2021 May 17;21(1):220. doi: 10.1186/s12886-021-01975-7. Early neurovascular changes in the retina in preclinical diabetic retinopathy and its relation with blood glucose. BMC Ophthalmol. 2021;21(1):220. doi: 10.1186/s12886-021-01975-7.

Mbatchou J, Barnard L, Backman J, Marcketta A, Kosmicki JA, Ziyatdinov A, Benner C, O’Dushlaine C, Barber M, Boutkov B, Habegger L, Ferreira M, Baras A, Reid J, Abecasis G, Maxwell E, Marchini J. Computationally efficient whole-genome regression for quantitative and binary traits. Nat Genet. 2021;53(7):1097–1103. doi: 10.1038/s41588-021-00870-7.

Michelucci U. An introduction to autoencoders. arXiv 2022.doi: 10.48550/arXiv.2201.03898.

Myers TA, Chanock SJ, Machiela MJ. *LDlinkR*: An R package for rapidly calculating linkage disequilibrium statistics in diverse populations. Front Genet. 2020;11:157. doi: 10.3389/fgene.2020.00157.

Ochoa D, Hercules A, Carmona M, Suveges D, Gonzalez-Uriarte A, Malangone C, Miranda A, Fumis L, Carvalho-Silva D, Spitzer M, Baker J, Ferrer J, Raies A, Razuvayevskaya O, Faulconbridge A, Petsalaki E, Mutowo P, Machlitt-Northen S, Peat G, McAuley E, Ong CK, Mountjoy E, Ghoussaini M, Pierleoni A, Papa E, Pignatelli M, Koscielny G, Karim M, Schwartzentruber J, Hulcoop DG, Dunham I, McDonagh EM. Open Targets Platform: supporting systematic drug-target identification and prioritisation. Nucleic Acids Res. 2021;49(D1):D1302–D1310. doi: 10.1093/nar/gkaa1027.

Orozco LD, Chen HH, Cox C, Katschke KJ Jr, Arceo R, Espiritu C, Caplazi P, Nghiem SS, Chen YJ, Modrusan Z, Dressen A, Goldstein LD, Clarke C, Bhangale T, Yaspan B, Jeanne M, Townsend MJ, van Lookeren Campagne M, Hackney JA. Integration of eQTL and a single-cell atlas in the human eye identifies causal genes for age-related macular degeneration. Cell Rep. 2020;30(4):1246–1259.e6. doi: 10.1016/j.celrep.2019.12.082.

Patel PJ, Foster PJ, Grossi CM, Keane PA, Ko F, Lotery A, Peto T, Reisman CA, Strouthidis NG, Yang Q; UK Biobank Eyes and Vision Consortium. Spectral-domain optical coherence tomography imaging in 67 321 adults: Associations with macular thickness in the UK Biobank Study. Ophthalmology. 2016;123(4):829–40. doi: 10.1016/j.ophtha.2015.11.009.

Pers TH, Karjalainen JM, Chan Y, Westra HJ, Wood AR, Yang J, Lui JC, Vedantam S, Gustafsson S, Esko T, Frayling T, Speliotes EK; Genetic Investigation of ANthropometric Traits (GIANT) Consortium; Boehnke M, Raychaudhuri S, Fehrmann RS, Hirschhorn JN, Franke L. Biological interpretation of genome-wide association studies using predicted gene functions. Nat Commun. 2015;6:5890. doi: 10.1038/ncomms6890.

Radhakrishnan A, Friedman SF, Khurshid S, Ng K, Batra P, Lubitz SA, Philippakis AA, Uhler C. Cross-modal autoencoder framework learns holistic representations of cardiovascular state. Nat Commun. 2023;14:2436. doi: 10.1038/s41467-023-38125-0.

Roman TS, Cannon ME, Vadlamudi S, Buchkovich ML, Wolford BN, Welch RP, Morken MA, Kwon GJ, Varshney A, Kursawe R, Wu Y, Jackson AU; National Institutes of Health Intramural Sequencing Center (NISC) Comparative Sequencing Program; Erdos MR, Kuusisto J, Laakso M, Scott LJ, Boehnke M, Collins FS, Parker SCJ, Stitzel ML, Mohlke KL. A Type 2 diabetes-associated functional regulatory variant in a pancreatic islet enhancer at the ADCY5 locus. Diabetes. 2017;66(9):2521–2530. doi: 10.2337/db17-0464.

Ronneberger O, Fischer P, Brox T. U-Net: Convolutional networks for biomedical image segmentation. arXiv. 2015.doi: 10.48550/arXiv.1505.04597.

Schroff F, Kalenichenko D, Philbin J. FaceNet: A unified embedding for face recognition and clustering. arXiv. 2015.doi: 10.48550/arXiv.1503.03832.

Oren O, Gersh BJ, Bhatt DL. Artificial intelligence in medical imaging: switching from radiographic pathological data to clinically meaningful endpoints. Lancet Digit Health. 2020;2(9):e486–e488. doi: 10.1016/S2589-7500(20)30160-6.

Sheffield VC, Stone EM. Genomics and the eye. N Engl J Med. 2011;364(20):1932–42. doi: 10.1056/NEJMra1012354.

Shi Y, Zhang W, Yang Y, Murzin AG, Falcon B, Kotecha A, van Beers M, Tarutani A, Kametani F, Garringer HJ, Vidal R, Hallinan GI, Lashley T, Saito Y, Murayama S, Yoshida M, Tanaka H, Kakita A, Ikeuchi T, Robinson AC, Mann DMA, Kovacs GG, Revesz T, Ghetti B, Hasegawa M, Goedert M, Scheres SHW. Structure-based classification of tauopathies. Nature. 2021;598(7880):359-363. doi: 10.1038/s41586-021-03911-7.

Sollis E, Mosaku A, Abid A, Buniello A, Cerezo M, Gil L, Groza T, Güneş O, Hall P, Hayhurst J, Ibrahim A, Ji Y, John S, Lewis E, MacArthur JAL, McMahon A, Osumi-Sutherland D, Panoutsopoulou K, Pendlington Z, Ramachandran S, Stefancsik R, Stewart J, Whetzel P, Wilson R, Hindorff L, Cunningham F, Lambert SA, Inouye M, Parkinson H, Harris LW. The NHGRI-EBI GWAS Catalog: knowledgebase and deposition resource. Nucleic Acids Res. 2023;51(D1):D977–D985. doi: 10.1093/nar/gkac1010

Song H, Xiang Y, Jegelka S, Savarese S. Deep metric learning via lifted structured feature embedding. arXiv 2015.doi: 10.48550/arXiv.1511.06452.

Stefansson H, Helgason A, Thorleifsson G, Steinthorsdottir V, Masson G, Barnard J, Baker A, Jonasdottir A, Ingason A, Gudnadottir VG, Desnica N, Hicks A, Gylfason A, Gudbjartsson DF, Jonsdottir GM, Sainz J, Agnarsson K, Birgisdottir B, Ghosh S, Olafsdottir A, Cazier JB, Kristjansson K, Frigge ML, Thorgeirsson TE, Gulcher JR, Kong A, Stefansson K. A common inversion under selection in Europeans. Nat Genet. 2005;37(2):129–37. doi: 10.1038/ng1508.

Steinberg KM, Antonacci F, Sudmant PH, Kidd JM, Campbell CD, Vives L, Malig M, Scheinfeldt L, Beggs W, Ibrahim M, Lema G, Nyambo TB, Omar SA, Bodo JM, Froment A, Donnelly MP, Kidd KK, Tishkoff SA, Eichler EE. Structural diversity and African origin of the 17q21.31 inversion polymorphism. Nat Genet. 2012;44(8):872–80. doi: 10.1038/ng.2335.

Thomas ED, Timms AE, Giles S, Harkins-Perry S, Lyu P, Hoang T, Qian J, Jackson VE, Bahlo M, Blackshaw S, Friedlander M, Eade K, Cherry TJ. Cell-specific cis-regulatory elements and mechanisms of non-coding genetic disease in human retina and retinal organoids. Dev Cell. 2022;57(6):820–836.e6. doi: 10.1016/j.devcel.2022.02.018.

Turley P, Walters RK, Maghzian O, Okbay A, Lee JJ, Fontana MA, Nguyen-Viet TA, Wedow R, Zacher M, Furlotte NA; 23andMe Research Team; Social Science Genetic Association Consortium; Magnusson P, Oskarsson S, Johannesson M, Visscher PM, Laibson D, Cesarini D, Neale BM, Benjamin DJ. Multi-trait analysis of genome-wide association summary statistics using MTAG. Nat Genet. 2018;50(2):229–237. doi: 10.1038/s41588-017-0009-4.

Wang Y, Mandelkow E. Tau in physiology and pathology. Nat Rev Neurosci. 2016;17(1):5–21. doi: 10.1038/nrn.2015.1.

Xie Z, Zhang T, Kim S, Lu J, Zhang W, Lin C, Wu M, Davis A, Channa R, Giancardo L, Chen H, Wang S, Chen R, Zhi D. iGWAS: image-based genome-wide association of self-supervised deep phenotyping of human medical images. medRxiv 2023. doi: 10.1101/2022.05.26.22275626.

Yang J, Ferreira T, Morris AP, Medland SE; Genetic Investigation of ANthropometric Traits (GIANT) Consortium; DIAbetes Genetics Replication And Meta-analysis (DIAGRAM) Consortium; Madden PA, Heath AC, Martin NG, Montgomery GW, Weedon MN, Loos RJ, Frayling TM, McCarthy MI, Hirschhorn JN, Goddard ME, Visscher PM. Conditional and joint multiple-SNP analysis of GWAS summary statistics identifies additional variants influencing complex traits. Nat Genet. 2012;44(4):369–75, S1-3. doi: 10.1038/ng.2213.

Yim J, Chopra R, Spitz T, Winkens J, Obika A, Kelly C, Askham H, Lukic M, Huemer J, Fasler K, Moraes G, Meyer C, Wilson M, Dixon J, Hughes C, Rees G, Khaw PT, Karthikesalingam A, King D, Hassabis D, Suleyman M, Back T, Ledsam JR, Keane PA, De Fauw J. Predicting conversion to wet age-related macular degeneration using deep learning. Nat Med. 2020;26(6):892–899. doi: 10.1038/s41591-020-0867-7.

Zekavat SM, Jorshery SD, Shweikh Y, Horn K, Rauscher FG, Sekimitsu S, Kayoma S, Ye Y, Raghu V, Zhao H, Ghassemi M, Elze T, Segre AV, Wiggs JL, Scholz M, Del Priore L, Wang JC, Natarajam P, Zebardast N. medRxiv 2023. doi: 10.1101/2023.05.16.23290063

Zhao B, Li Y, Fan Z, Wu Z, Shu J, Yang X, Yang Y, Wang X, Li B, Wang X, Copana C, Yang Y, Lin J, Li Y, Stein JL, O’Brien JM, Li T, Zhu H. Eye-brain connections revealed by multimodal retinal and brain imaging genetics in the UK Biobank. medRxiv 2023. doi: 10.1101/2023.02.16.23286035

