## Supplementary Information for "Autoencoder-based phenotyping of ophthalmic images highlights genetic loci influencing retinal morphology and provides informative biomarkers"

### SUPPLEMENTARY FIGURES

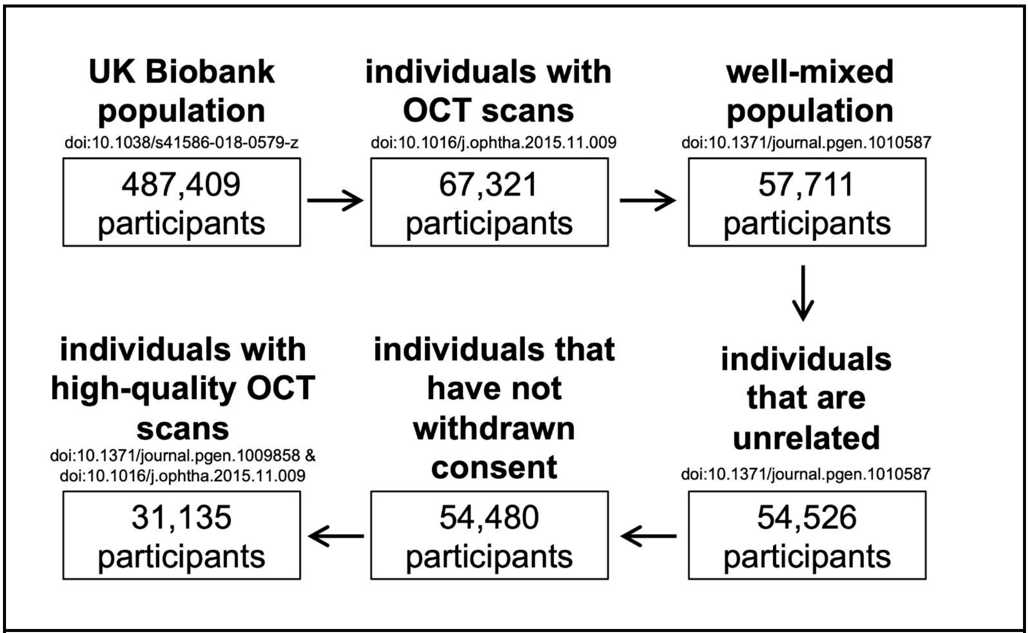


**Supplementary Figure 1.** Quality control workflow.
Genotypic information and imaging parameters were taken into account.

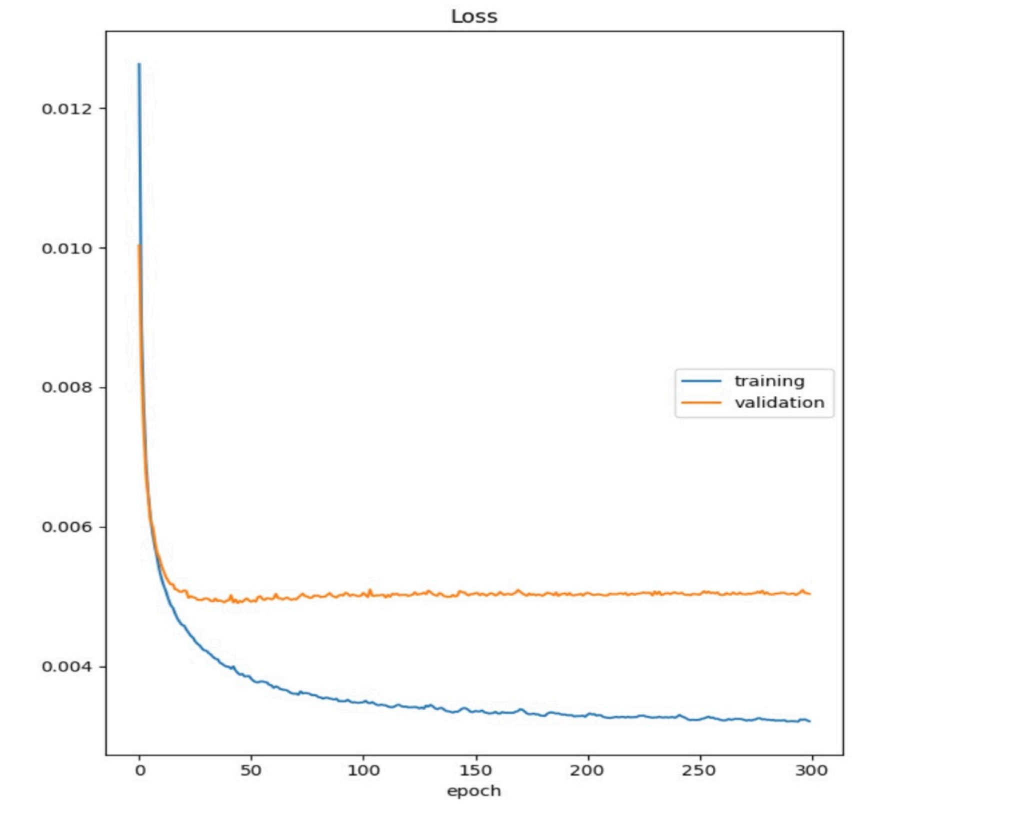


**Supplementary Figure 2.** Autoencoder training. The image reconstruction loss mean squared error (MSE) is plotted against the number of epochs used in the context of the training and the validation sets (consisting of 2500 and 500 images respectively). Autoencoder training was conducted using batches of 80 images for each epoch over a total of 300 epochs.

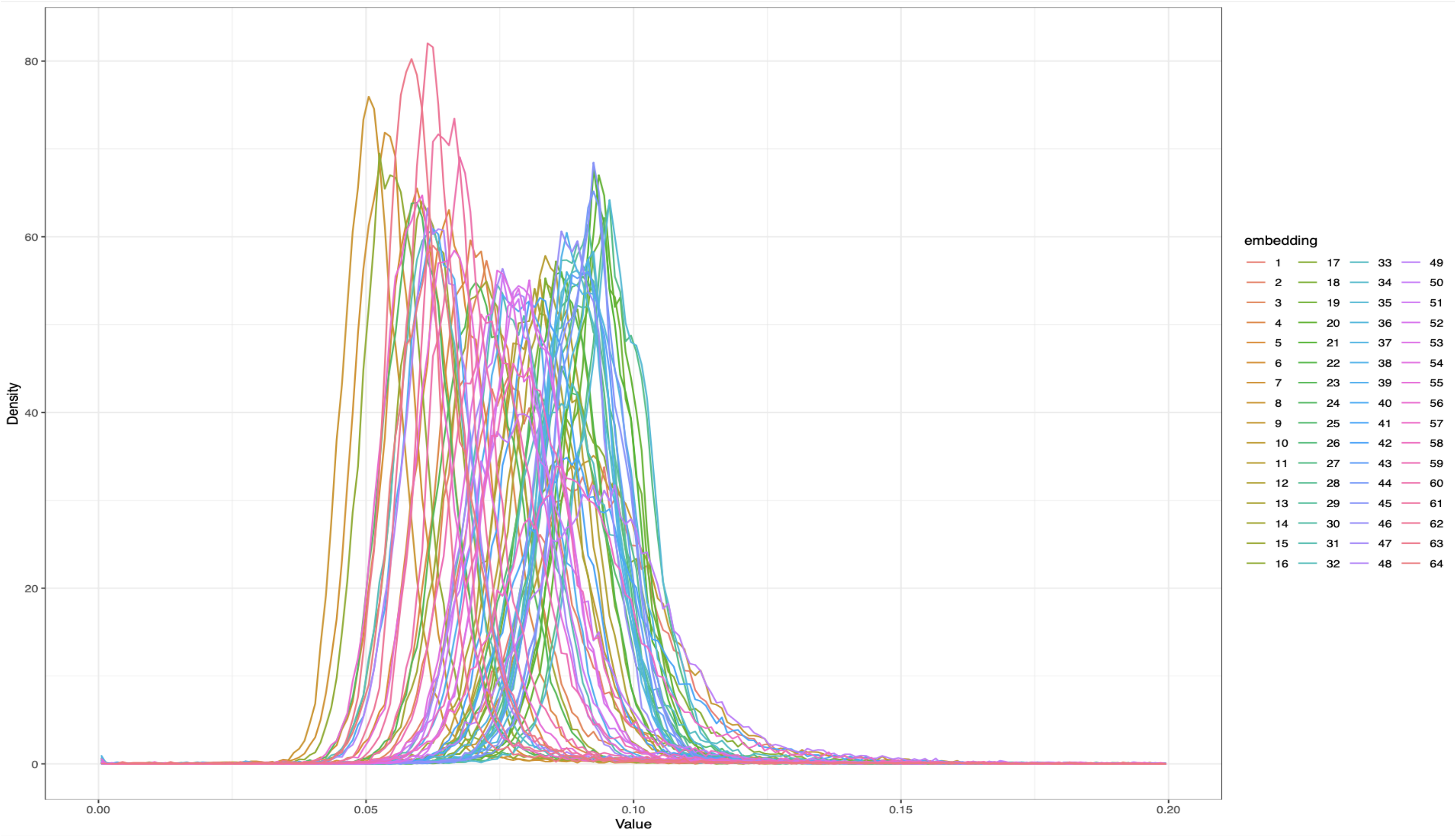


**Supplementary Figure 3.** Overlaid histograms outlining the univariate distribution for each of the 64 embeddings.

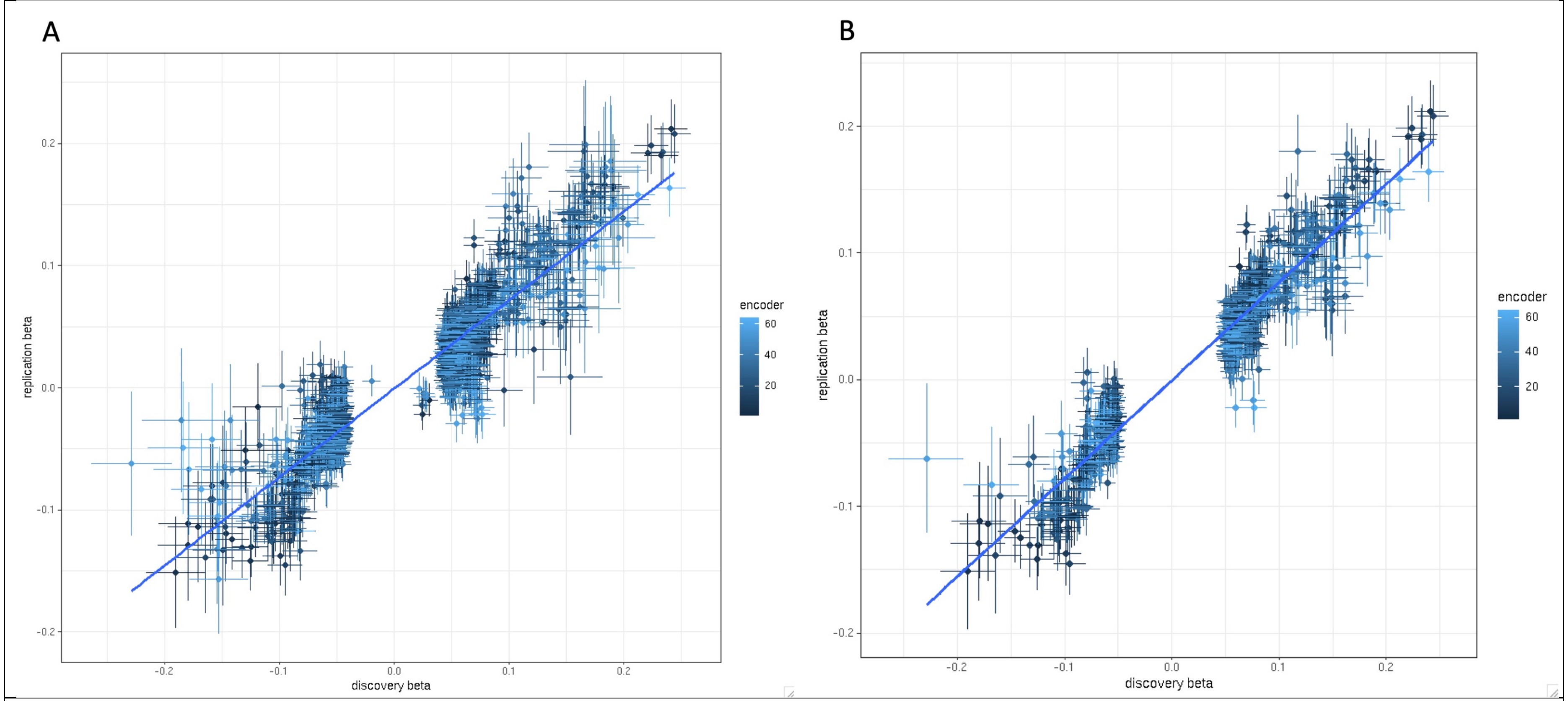


**Supplementary Figure 4.** Beta – beta plot showing the correlation between the effect sizes derived from the genetic association studies in the discovery and replication UK Biobank cohorts. The effect sizes for all fine mapped, statistically significant signals obtained using either a genome-wide significance threshold (p-value < $5 \times 10^{-8}$) (A) or a more conservative study-wide threshold (p-value < $3.2 \times 10^{-10}$ following Bonferroni correction for 153 tests) (B) are shown. The relevant $R^2$ coefficient was 0.84 for genome-wide significant variants and 0.95 for study-wide significant changes.

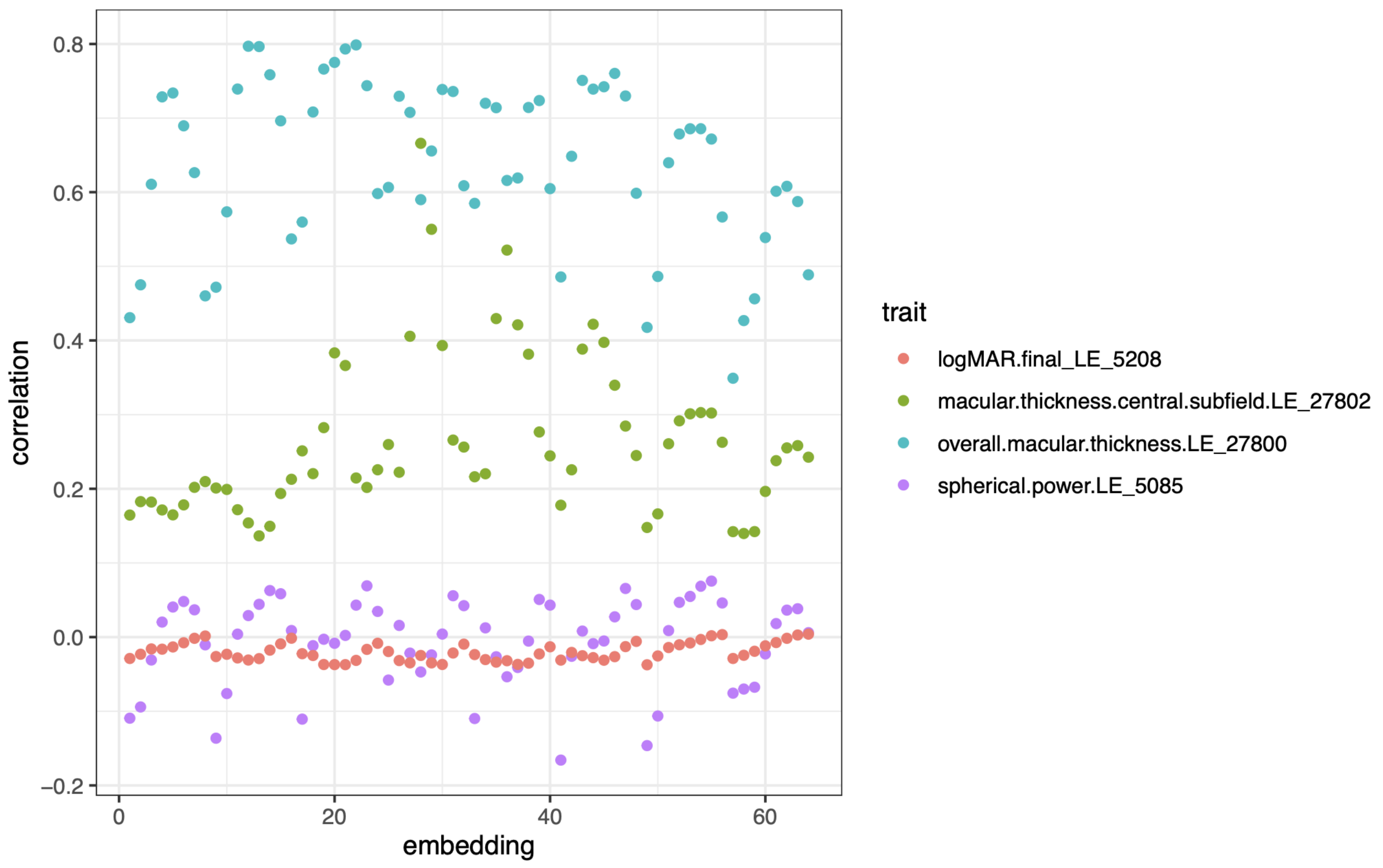


**Supplementary Figure 5.** Correlation between each of the 64 embeddings and four key ophthalmic phenotypes. These include left eye visual acuity (logMAR.final_LE_5208), average thickness of the left retina in a central circle with a 1 mm diameter (macular.thickness.central.subfield.LE_27802), average thickness of the left retina in a central circle with a 6 mm diameter (overall.macular.thickness.LE_27800) and a left eye size surrogate (spherical.power.LE_5085). The Pearson correlation coefficient R is shown in the y-axis

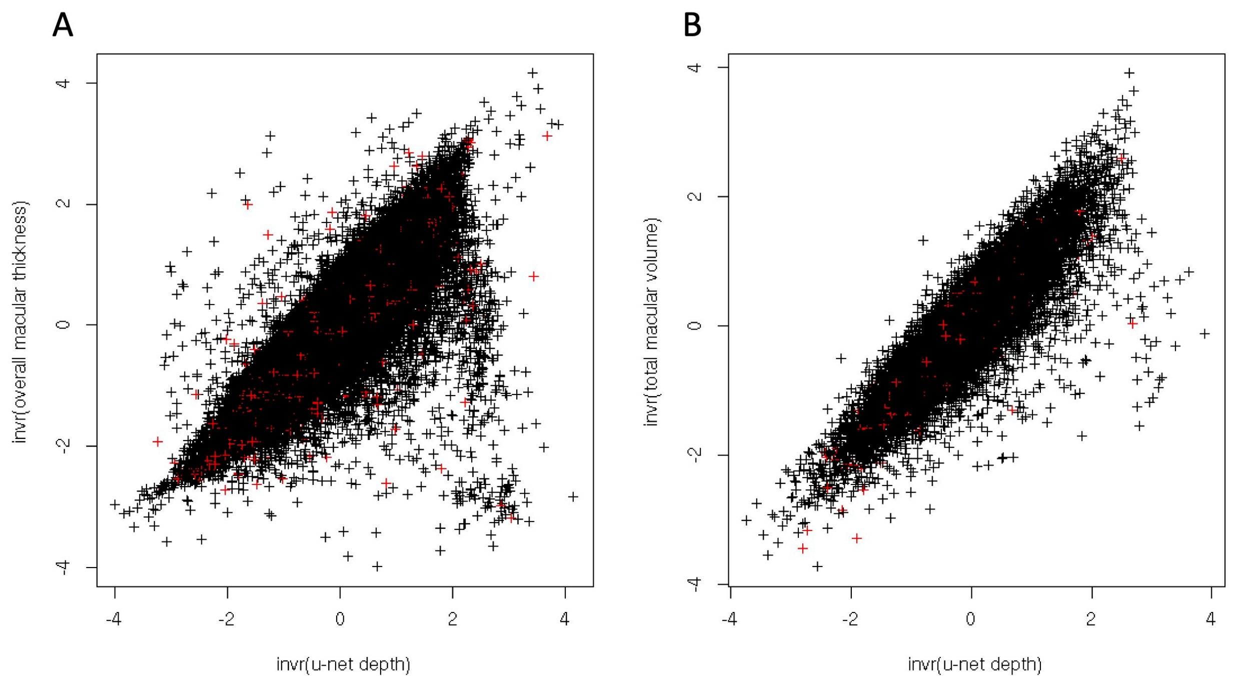


**Supplementary Figure 6.** Correlation between retinal thickness measurements obtained from UK Biobank left eye OCT scans using our U-Net based approach and either the "total macular volume" measurements obtained using the Topcon Advanced Boundary Segmentation (TABS) software (data-field 27820) (A) or the "overall macular thickness" measurements obtained using the same purpose-built tool (data-field 27800). The relevant $R^2$ coefficient was 0.87 for the former (A) and 0.79 for the latter (B). It is noted that total macular volume assesses retinal thickness across the whole image (similarly to our approach) while overall macular thickness evaluates thickness in a central circle with a 6 mm diameter.

To evaluate the performance of our U-Net based approach in retinae with pathology we identified UK Biobank participants that have been assigned one of the ICD-10 codes in the H35 group ("Other retinal disorders", including age-related macular degeneration) (data-field 41270). In these cases, the measurements obtained using our U-Net again correlated well with those from the TABS software (red crosses in the graph) The relevant $R^2$ coefficient was 0.90 and 0.80 for "total macular volume" and "overall macular thickness" respectively.

### SUPPLEMENTARY FILE

**Supplementary File 1.** Left eye retinal thickness maps showing the difference in retinal structure between individuals with different alleles in each of the 118 lead loci (that were found to be statistically significantly associated with one or more autoencoder-derived retinal OCT phenotypes).
Left: mean depth (thickness) representation for reference:reference alleles. Middle: difference between image mean for reference:reference and image mean for reference:non-reference (heterozygous) genotypes. Right: difference between image mean for reference:reference to image mean for non-reference:non-reference (homozygous) genotypes.

### SUPPLEMENTARY TABLES

**Supplementary Table 1**. Common genetic variants that were found to be significantly associated (p-value < 5e-08) with one or more autoencoder-derived retinal OCT phenotypes (n=239; following conditional and joint multiple-variant analysis (GCTA-COJO))

| |
|---|
| (see relevant spreadsheet) |

**Supplementary Table 2.** Previous studies using self-supervised deep learning approaches to generate autoencoder-derived retinal imaging phenotypes for GWAS

| reference | material for main analysis | analytical approach | main finding |
|---|---|---|---|
| Kirchler *et al*. 2022 | retinal fundus images of 46 731 participants of European-like ancestries from the UK Biobank | (i) an ANN was trained on an independent transfer task (to prime the network); images from EyePACS and ImageNet were used for this; (ii) the trained network was combined with PCA to condense UK Biobank fundus images into low-dimensional embeddings; (iii) GWAS analysis was performed using the generate embeddings as the phenotype | 60 genomic regions associated with retinal fundus imaging features |
| Xie *et al*. 2023 | retinal fundus images (raw or segmented vessel masks) of 65,629 participants of European-like ancestries from the UK Biobank | (i) an ANN was trained on an independent transfer task (to prime the network); images from EyePACS were used for this; (ii) the trained network was used to condense UK Biobank fundus images into 128 low-dimensional embeddings; (iii) GWAS analyses was performed using the generated embeddings as the phenotype | 14+22 genomic regions associated with retinal fundus imaging features |

In contrast to the above two studies, our work utilised UK Biobank OCT scans (instead of fundus photographs). OCT has higher resolution and information content, and allows evaluation of retinal thickness, a key morphological parameter. Notably, important source of variability in fundus photographs is the significant variation in the levels of natural fundus pigmentation across the population; the impact of this on OCT scans is much less pronounced.

Another study that used self-supervised deep learning to produce imaging-derived phenotypes with a focus on performing GWAS is the one by Bonazzola *et al.* 2023; this utilised cardiac magnetic resonance (CMR) images and identified 11 significant loci.

GWAS, genome-wide association study; ANN, artificial neural network, OCT, optical coherence tomography.

### REFERENCES


- Bonazzola R, Ferrante E, Ravikumar N, Xia Y, Keavney B, Plein S, Syeda-Mahmood T, Frangi AF. Unsupervised ensemble-based phenotyping helps enhance the discoverability of genes related to heart morphology. arXiv 2023. doi: 10.48550/arXiv.2301.02916.
- Kirchler M, Konigorski S, Norden M, Meltendorf C, Kloft M, Schurmann C, Lippert C. transferGWAS: GWAS of images using deep transfer learning. Bioinformatics. 2022;38(14):3621-3628. doi: 10.1093/bioinformatics/btac369.
- Xie Z, Zhang T, Kim S, Lu J, Zhang W, Lin C, Wu M, Davis A, Channa R, Giancardo L, Chen H, Wang S, Chen R, Zhi D. iGWAS: image-based genome-wide association of self-supervised deep phenotyping of human medical images. *medRxiv* 2022. doi: 10.1101/2022.05.26.22275626.
