## Supplementary File 1 for "Autoencoder-based phenotyping of ophthalmic images highlights genetic loci influencing retinal morphology and provides informative biomarkers"

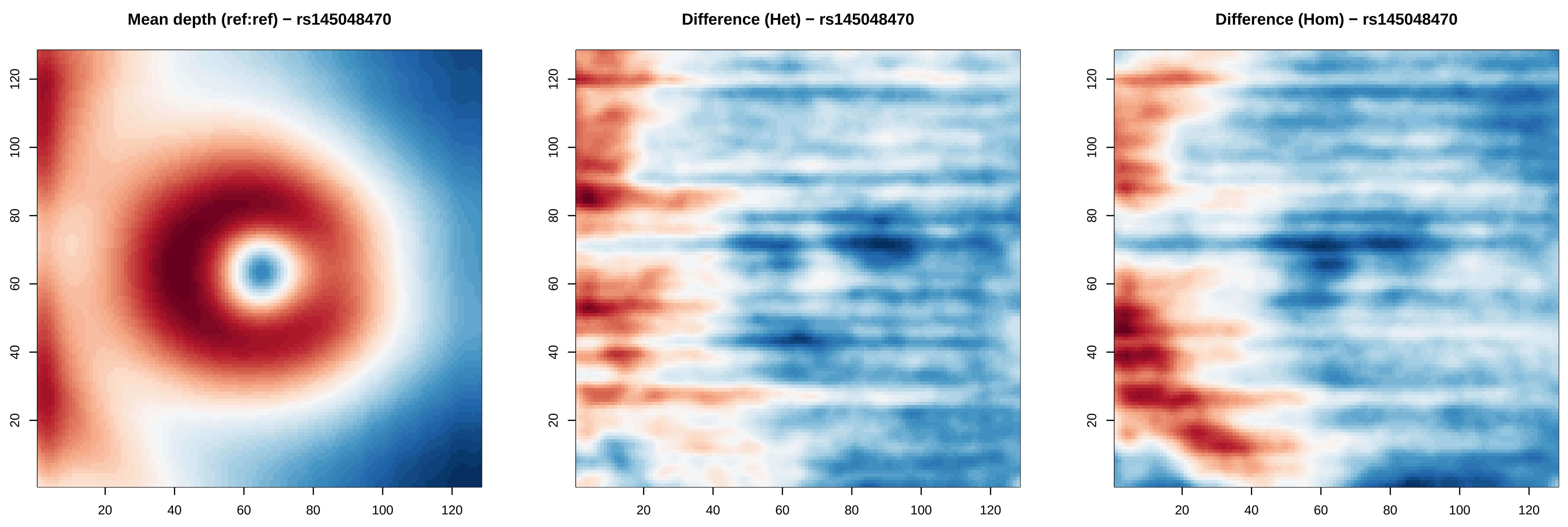
Mean depth (ref:ref) − rs145048470
Difference (Het) − rs145048470
Difference (Hom) − rs145048470
20
40
60
80
100
120

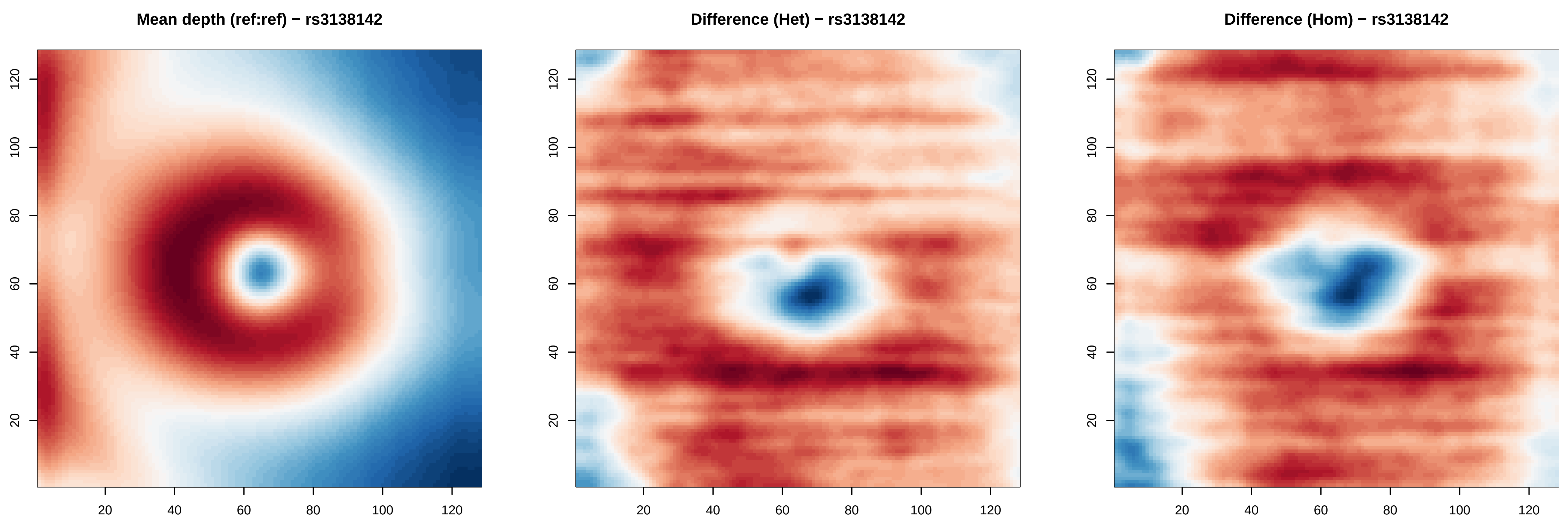

Mean depth (ref:ref) − rs3138142
Difference (Het) − rs3138142
Difference (Hom) − rs3138142
20
40
60
80
100
120

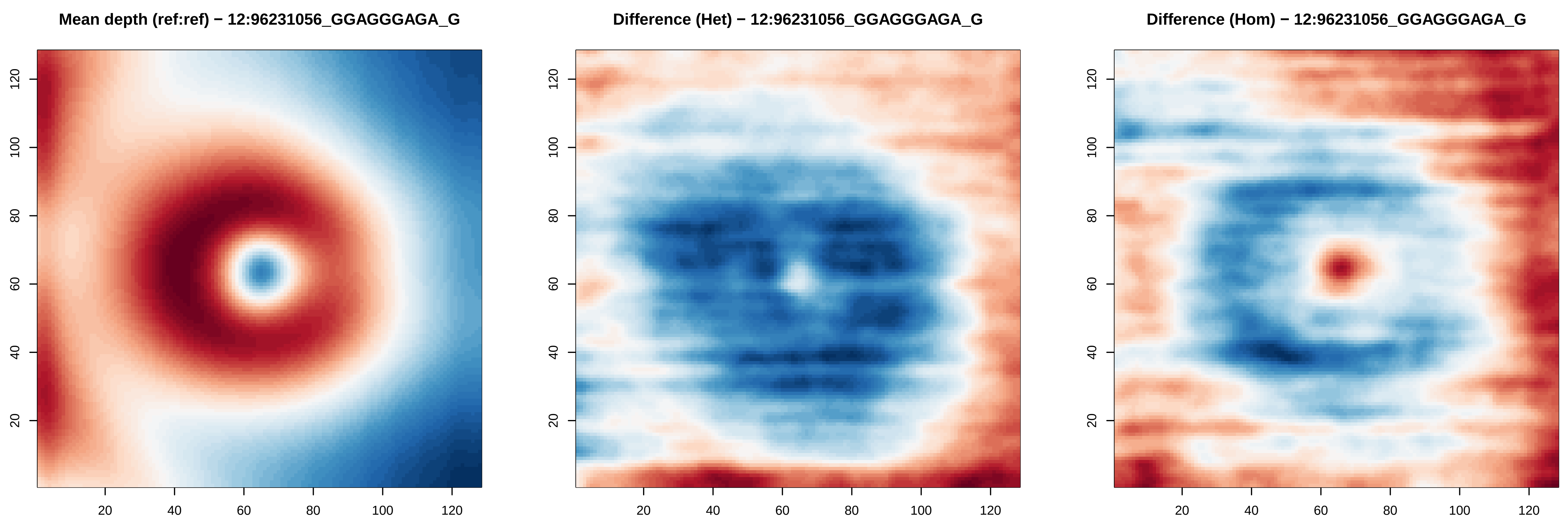
Mean depth (ref:ref) − 12:96231056_GGAGGGAGA_G
Difference (Het) − 12:96231056_GGAGGGAGA_G
Difference (Hom) − 12:96231056_GGAGGGAGA_G
20
40
60
80
100
120

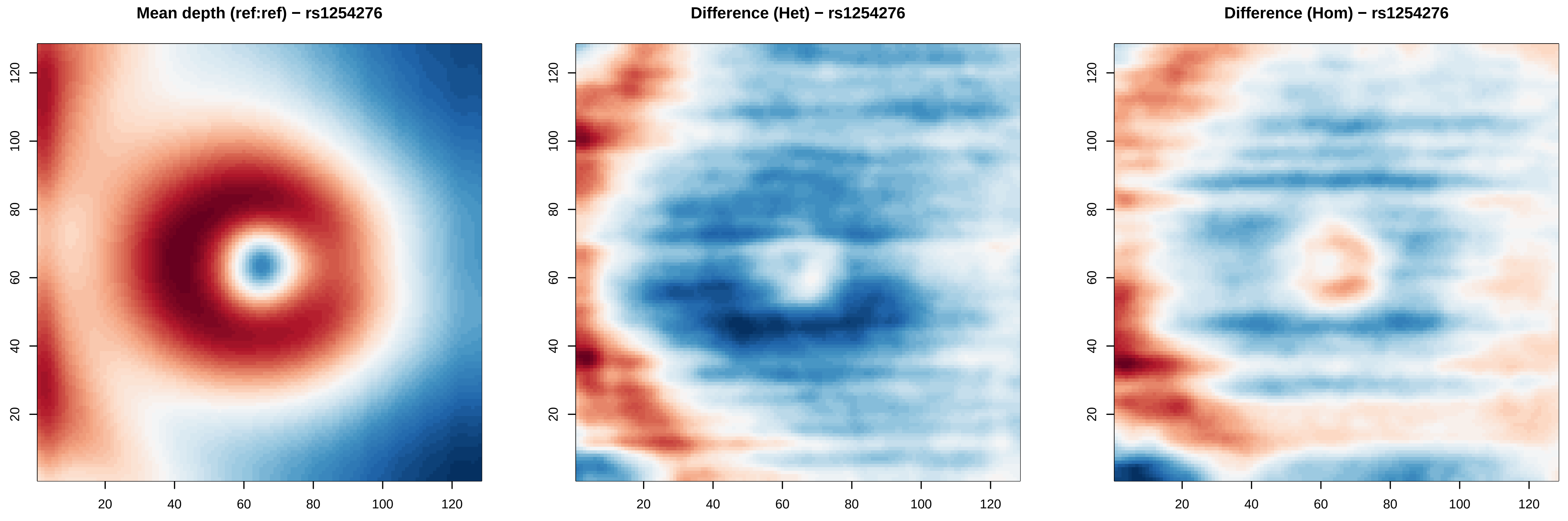

Mean depth (ref:ref) − rs1254276
Difference (Het) − rs1254276
Difference (Hom) − rs1254276
20
40
60
80
100
120

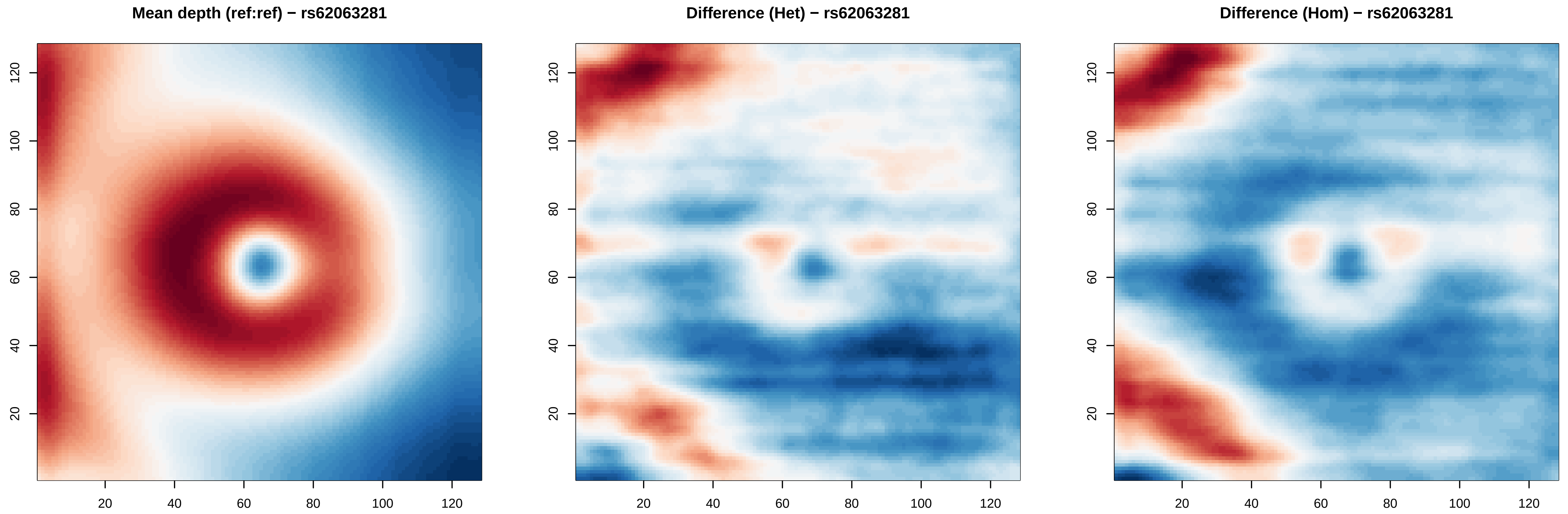
Mean depth (ref:ref) − rs62063281
Difference (Het) − rs62063281
Difference (Hom) − rs62063281
20
40
60
80
100
120

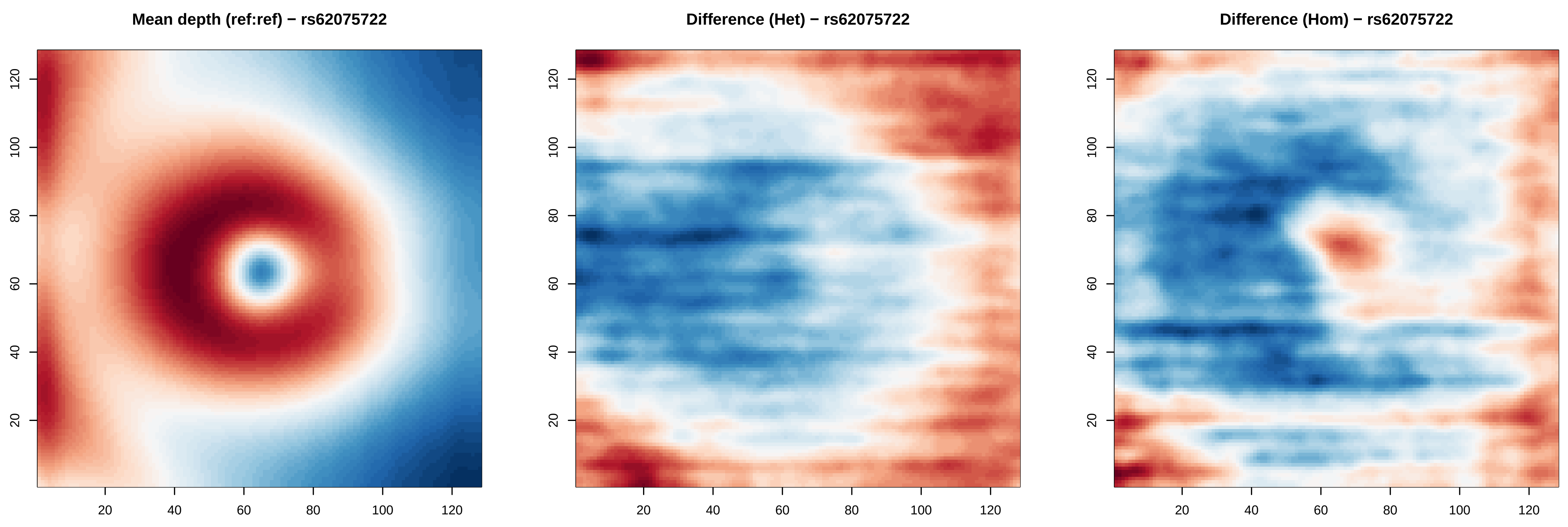

Mean depth (ref:ref) − rs62075722
Difference (Het) − rs62075722
Difference (Hom) − rs62075722
20
40
60
80
100
120

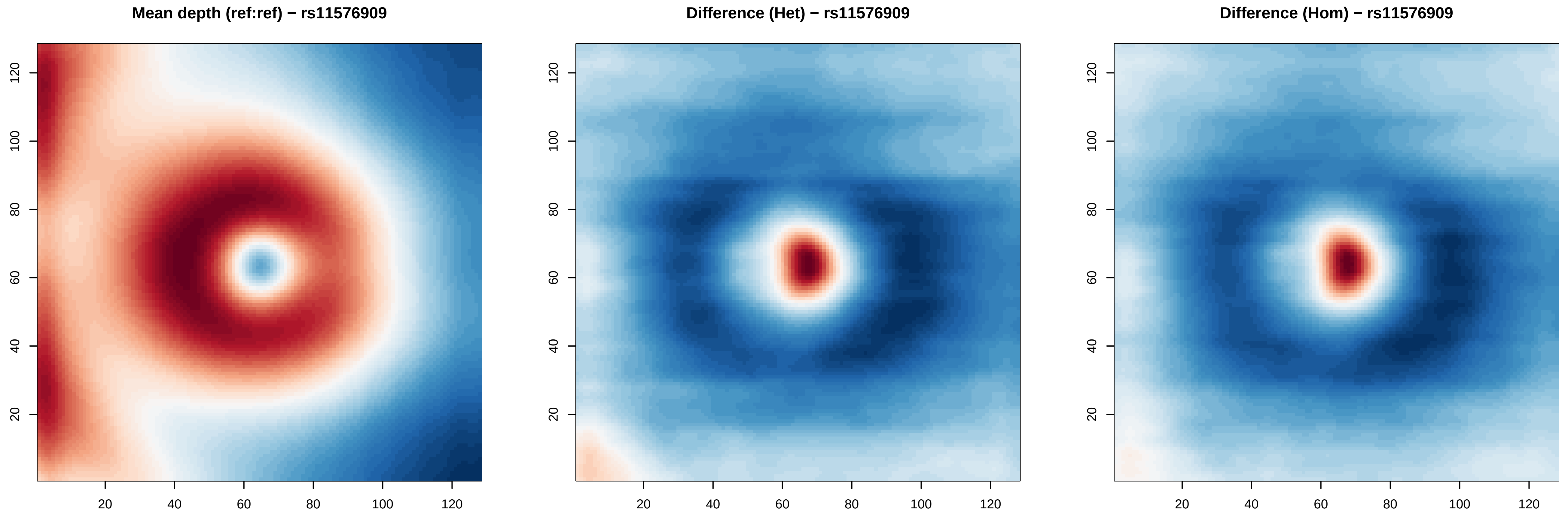

Mean depth (ref:ref) − rs11576909
Difference (Het) − rs11576909
Difference (Hom) − rs11576909
20
40
60
80
100
120

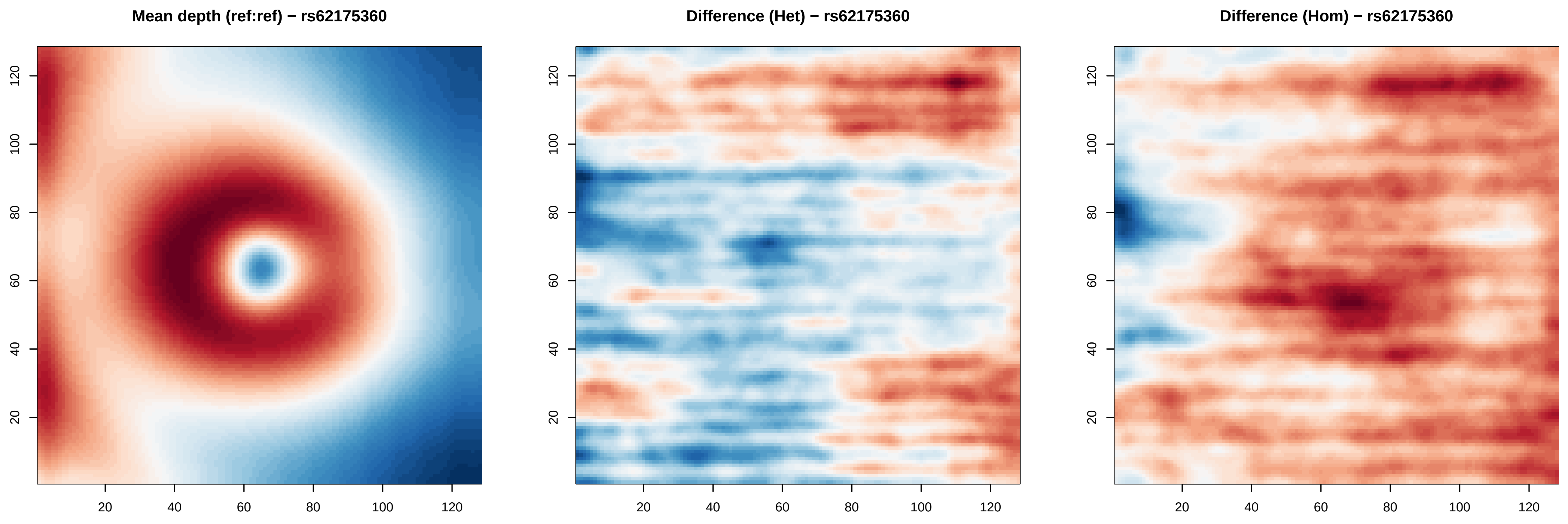

Mean depth (ref:ref) − rs62175360
Difference (Het) − rs62175360
Difference (Hom) − rs62175360
20
40
60
80
100
120

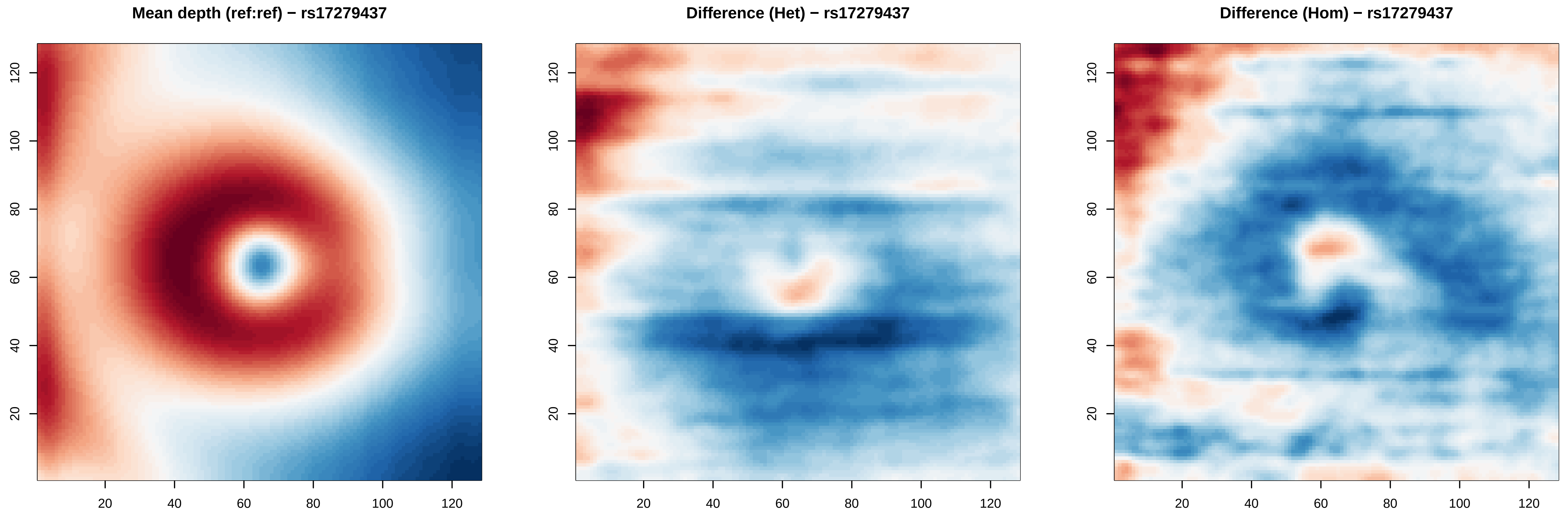
Mean depth (ref:ref) − rs17279437
Difference (Het) − rs17279437
Difference (Hom) − rs17279437
20
40
60
80
100
120

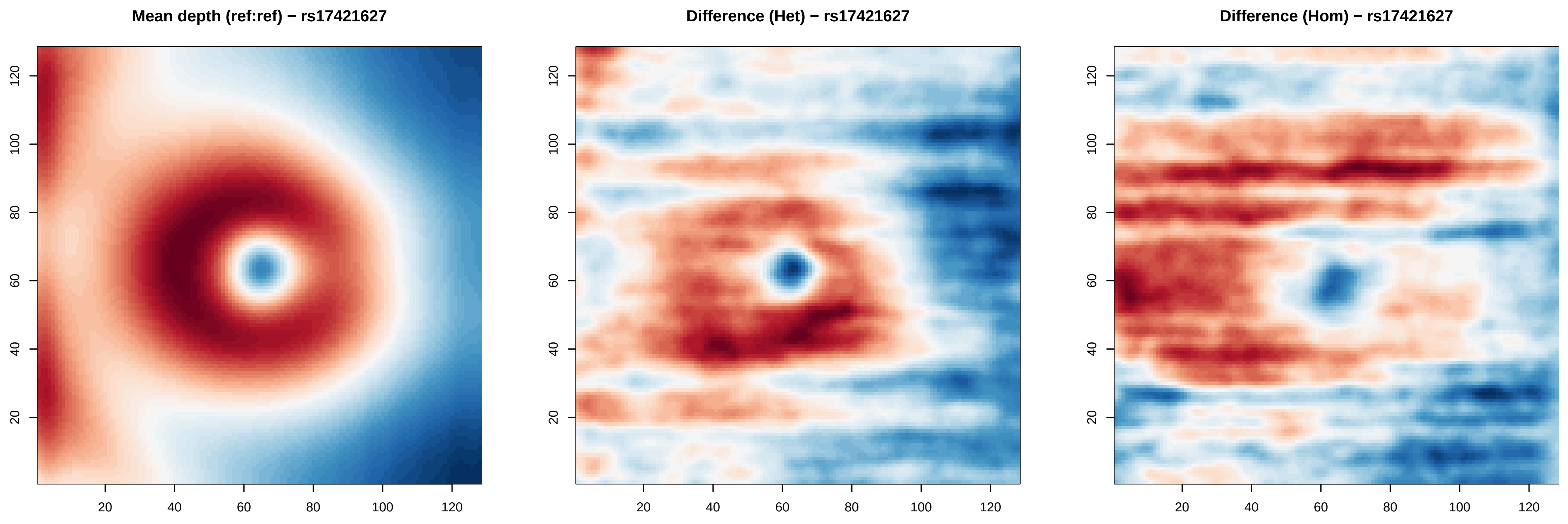
Mean depth (ref:ref) − rs17421627
Difference (Het) − rs17421627
Difference (Hom) − rs17421627
20
40
60
80
100
120

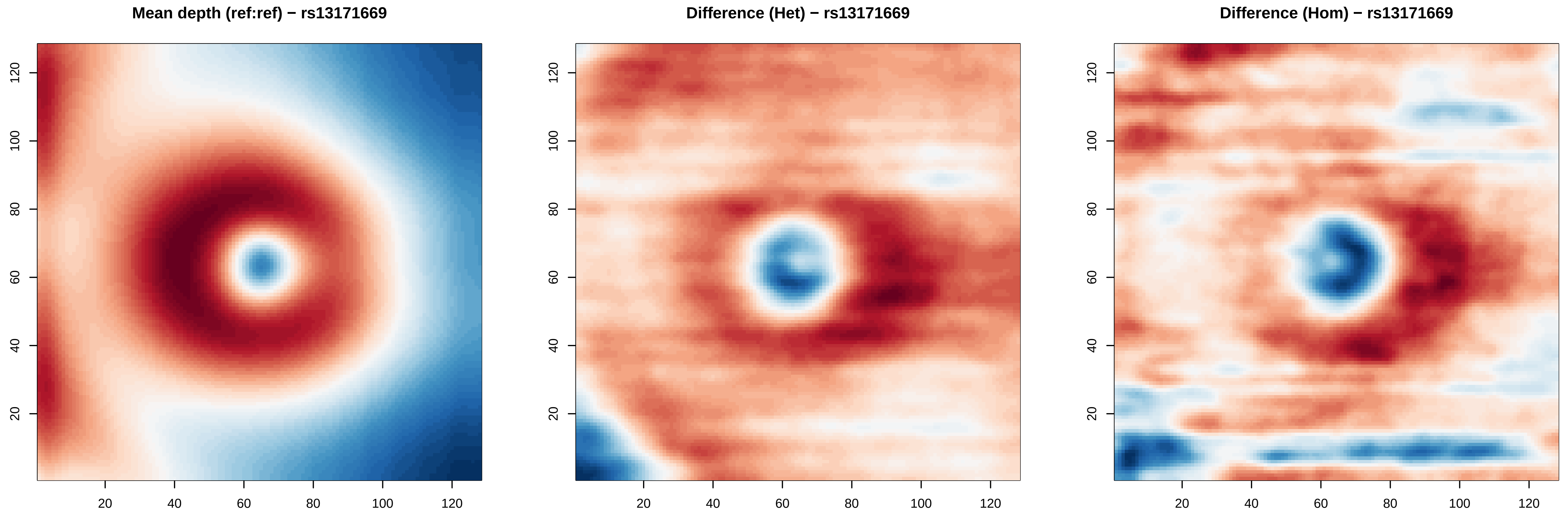
Mean depth (ref:ref) − rs13171669
Difference (Het) − rs13171669
Difference (Hom) − rs13171669
20
40
60
80
100
120

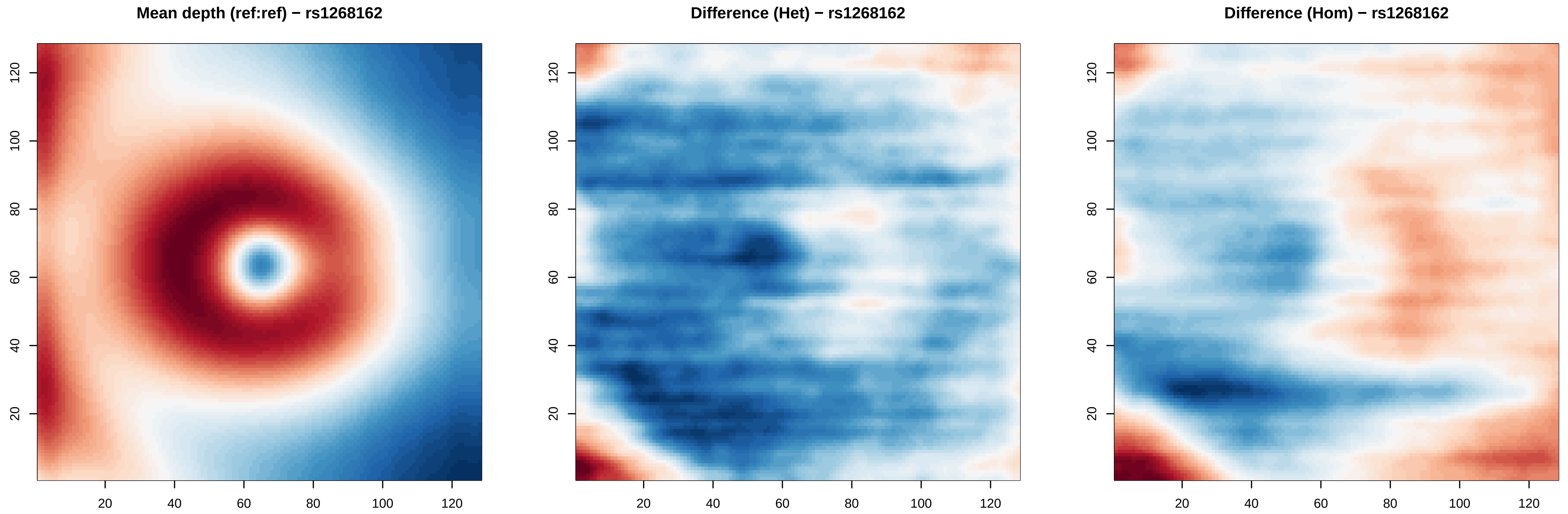

Mean depth (ref:ref) − rs1268162
Difference (Het) − rs1268162
Difference (Hom) − rs1268162
20
40
60
80
100
120

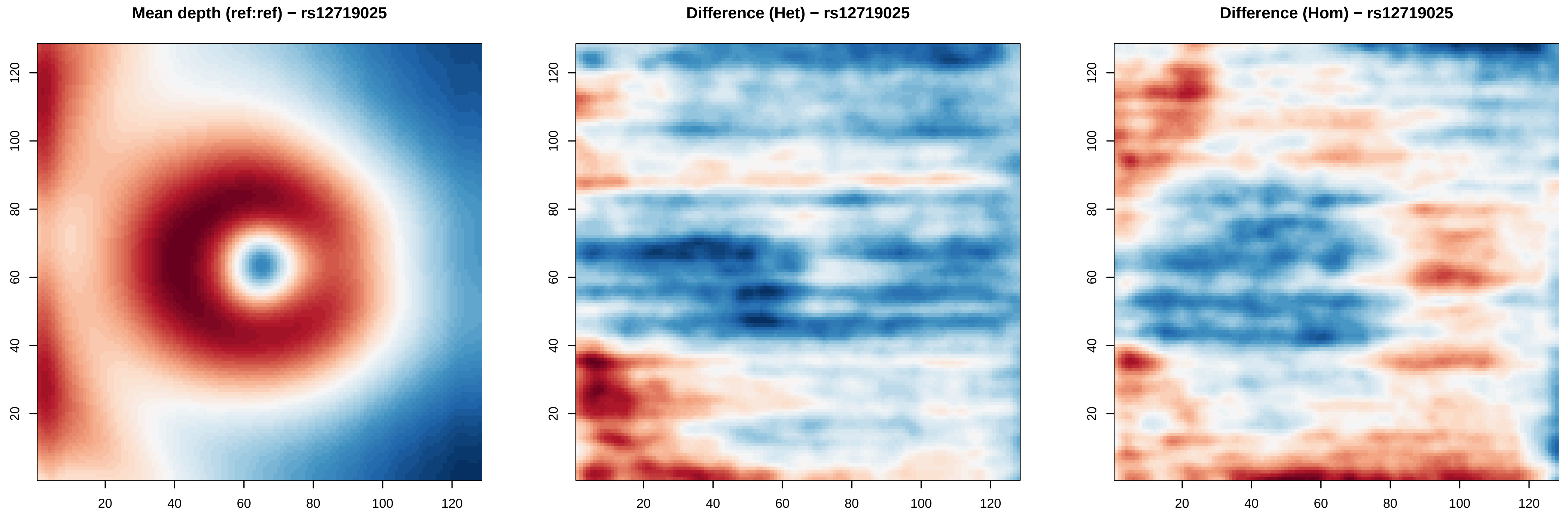
Mean depth (ref:ref) − rs12719025
Difference (Het) − rs12719025
Difference (Hom) − rs12719025
20
40
60
80
100
120

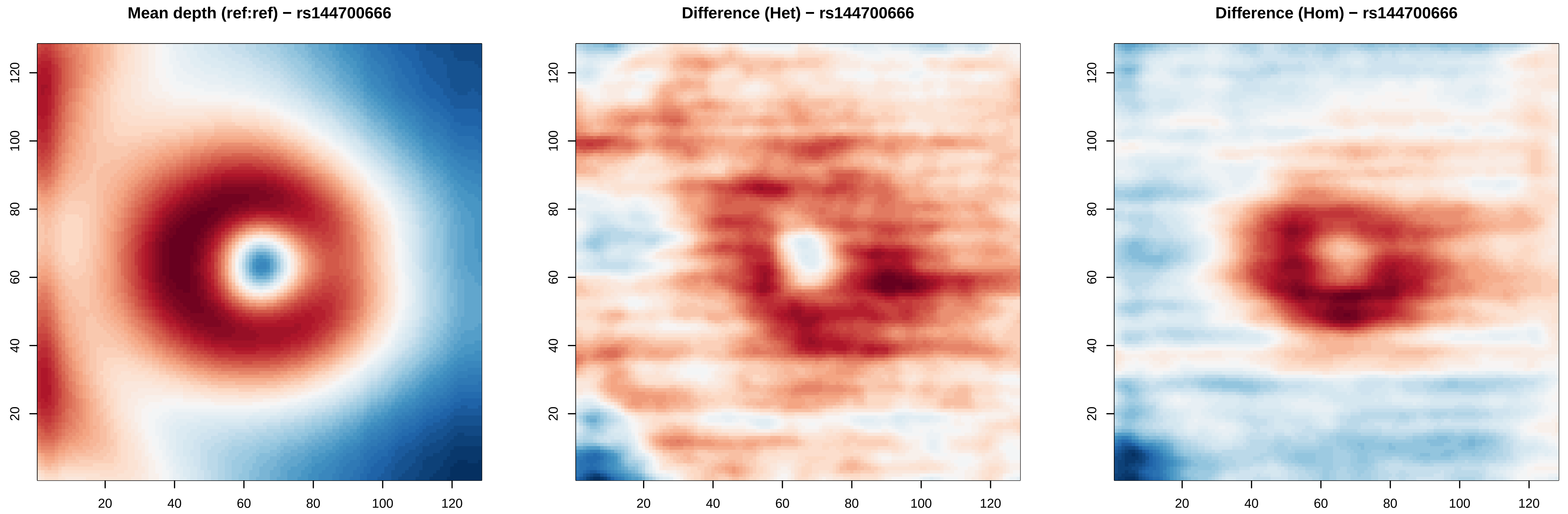

Mean depth (ref:ref) − rs144700666
Difference (Het) − rs144700666
Difference (Hom) − rs144700666
20
40
60
80
100
120

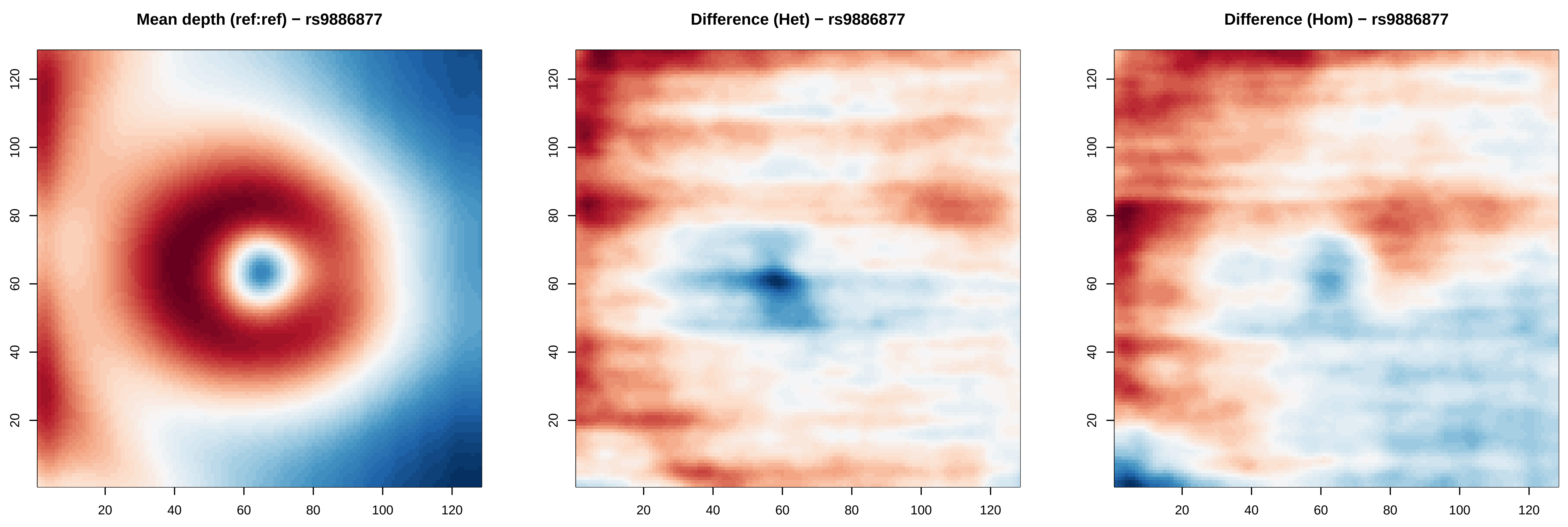
Mean depth (ref:ref) − rs9886877
Difference (Het) − rs9886877
Difference (Hom) − rs9886877
20
40
60
80
100
120

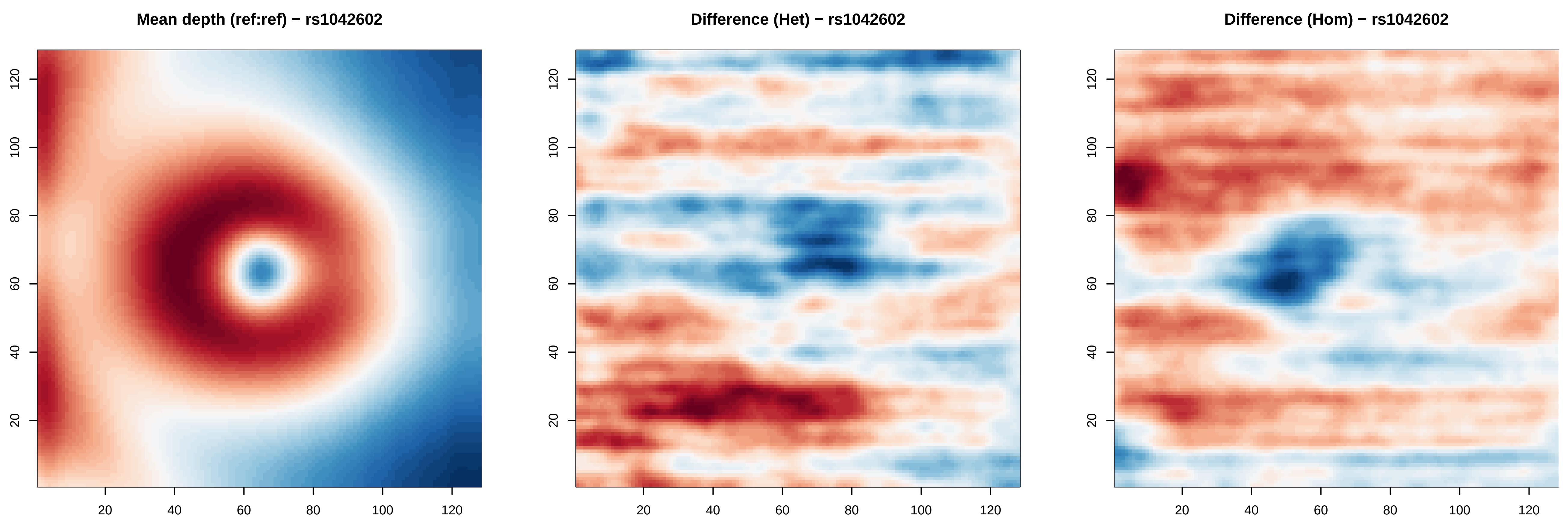
Mean depth (ref:ref) − rs1042602
Difference (Het) − rs1042602
Difference (Hom) − rs1042602
20
40
60
80
100
120

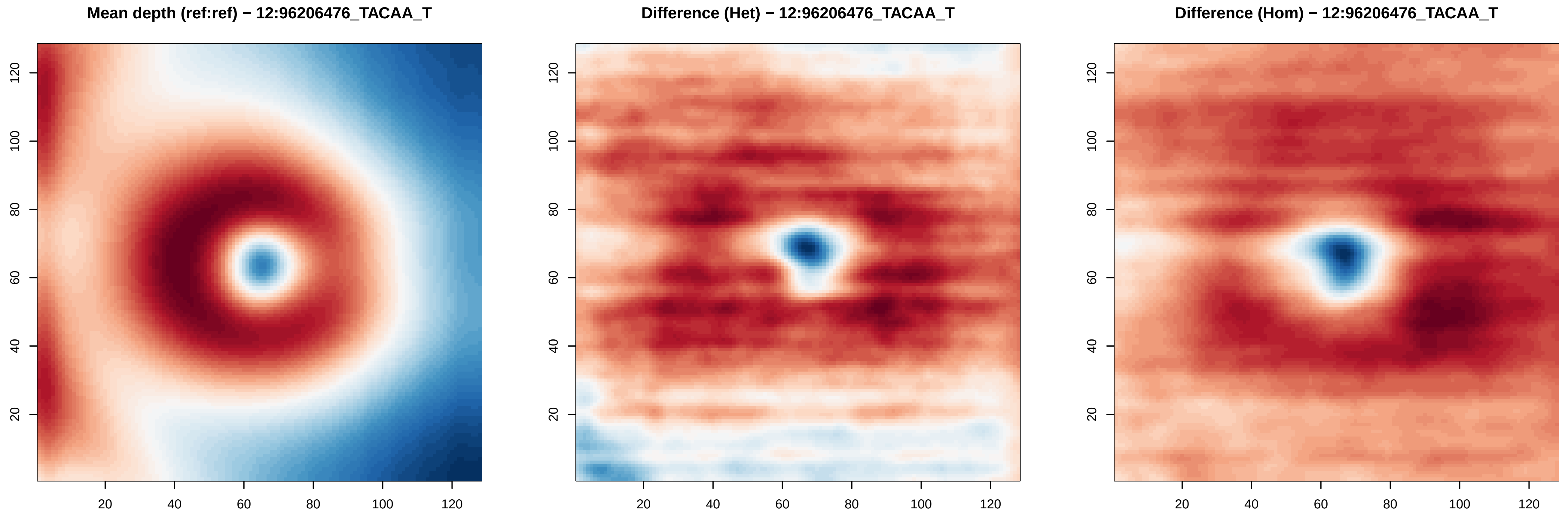

Mean depth (ref:ref) − 12:96206476_TACAA_T
Difference (Het) − 12:96206476_TACAA_T
Difference (Hom) − 12:96206476_TACAA_T
20 40 60 80 100 120
20 40 60 80 100 120

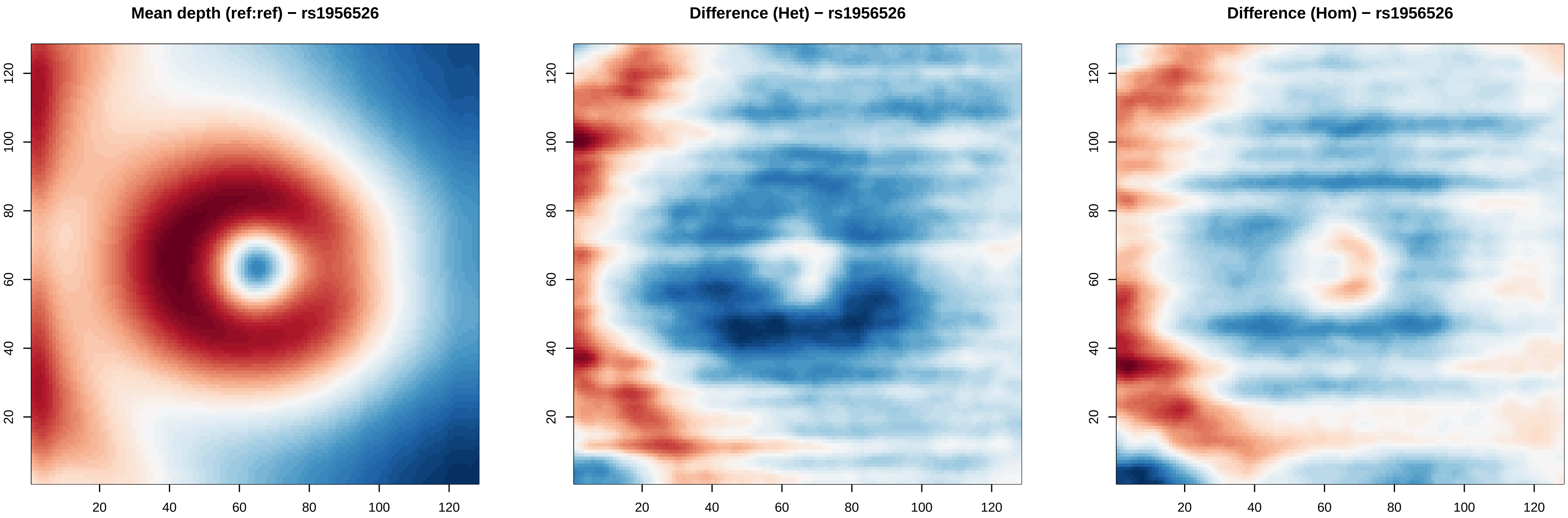
Mean depth (ref:ref) − rs1956526
Difference (Het) − rs1956526
Difference (Hom) − rs1956526
20
40
60
80
100
120

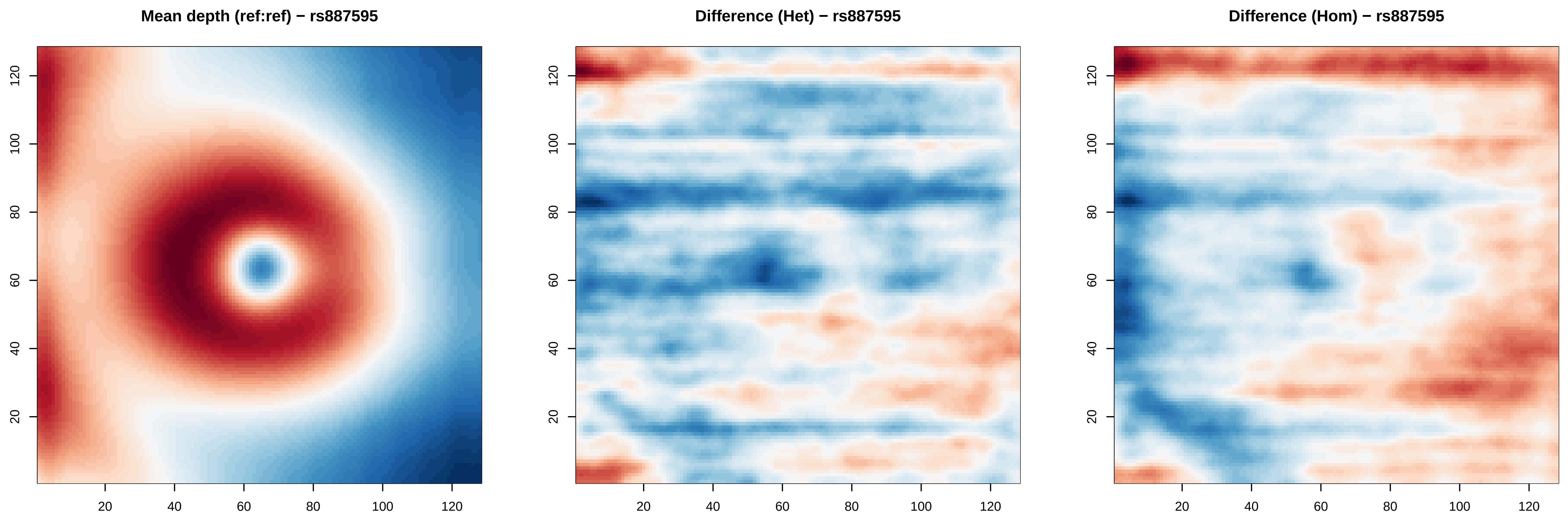
Mean depth (ref:ref) − rs887595
Difference (Het) − rs887595
Difference (Hom) − rs887595
20
40
60
80
100
120
20
40
60
80
100
120

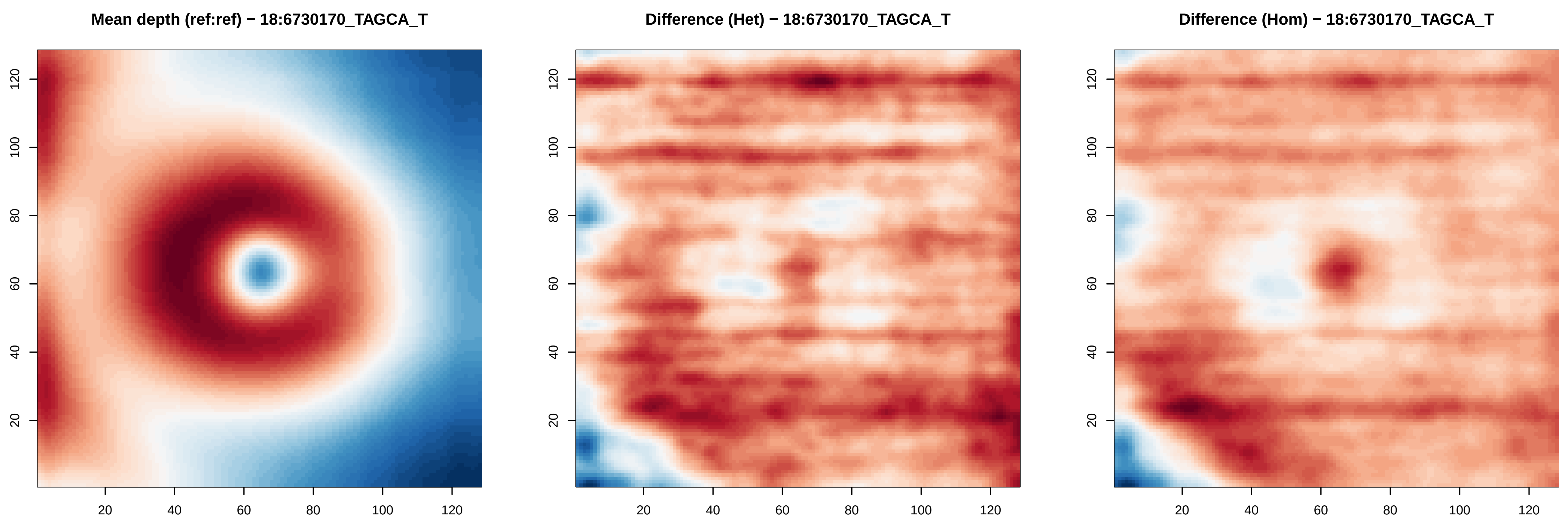

Mean depth (ref:ref) − 18:6730170_TAGCA_T
Difference (Het) − 18:6730170_TAGCA_T
Difference (Hom) − 18:6730170_TAGCA_T
20
40
60
80
100
120

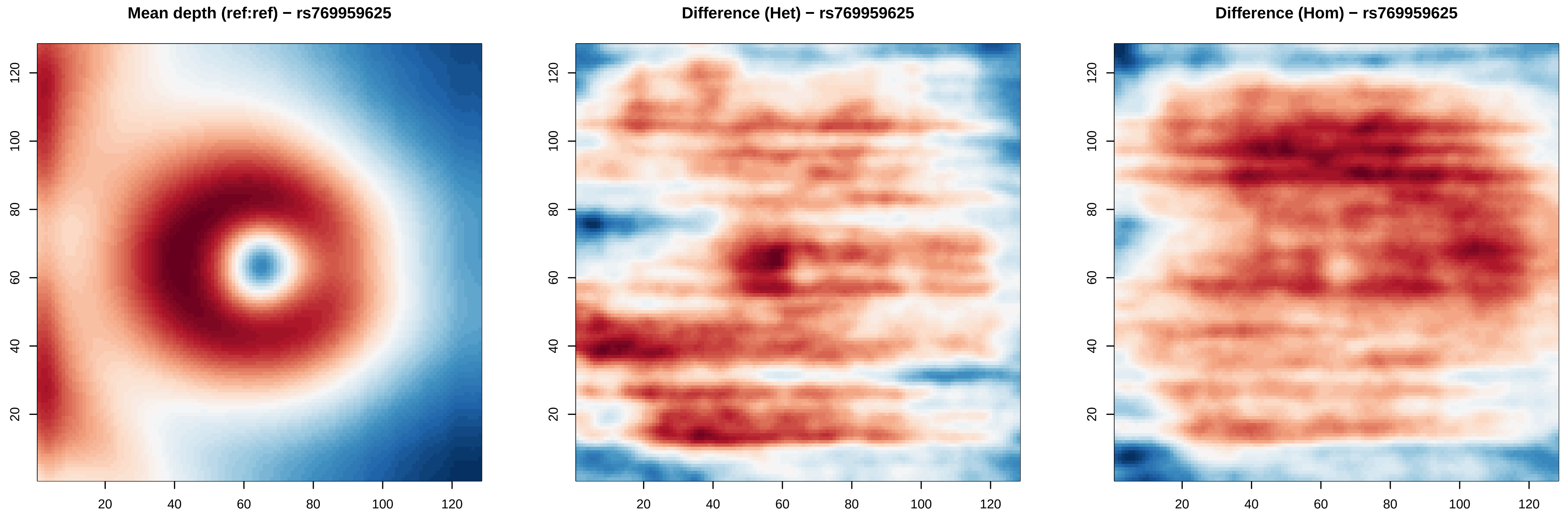
Mean depth (ref:ref) − rs769959625
Difference (Het) − rs769959625
Difference (Hom) − rs769959625
20
40
60
80
100
120

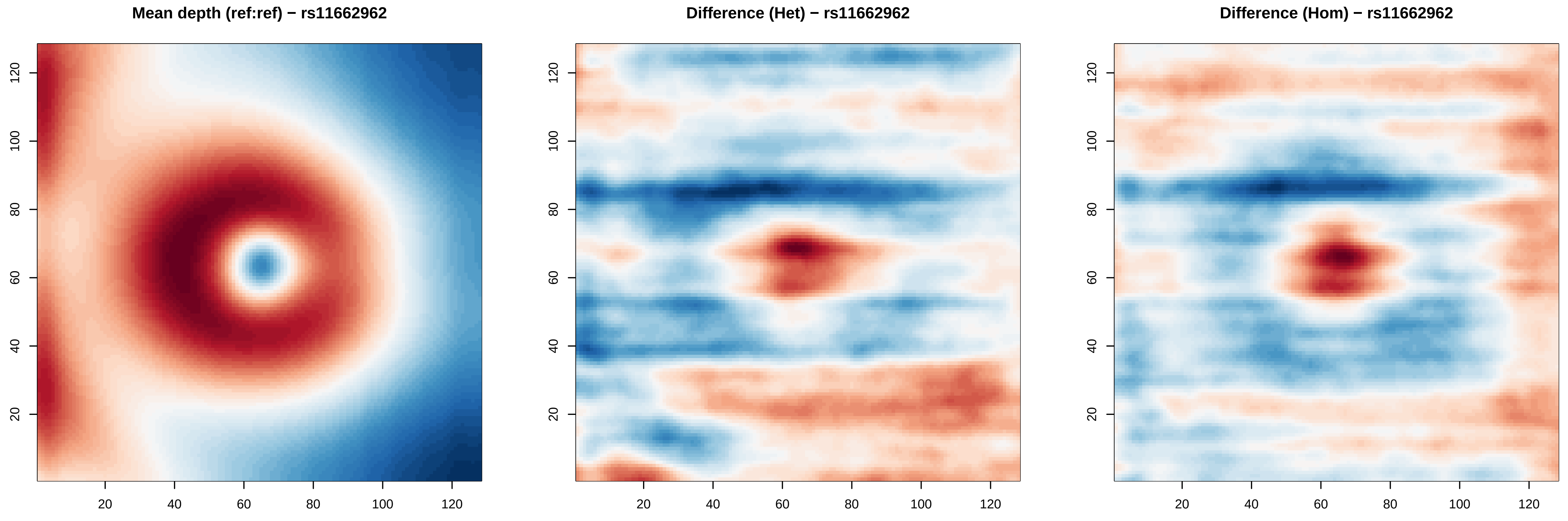

Mean depth (ref:ref) − rs11662962
Difference (Het) − rs11662962
Difference (Hom) − rs11662962
20
40
60
80
100
120

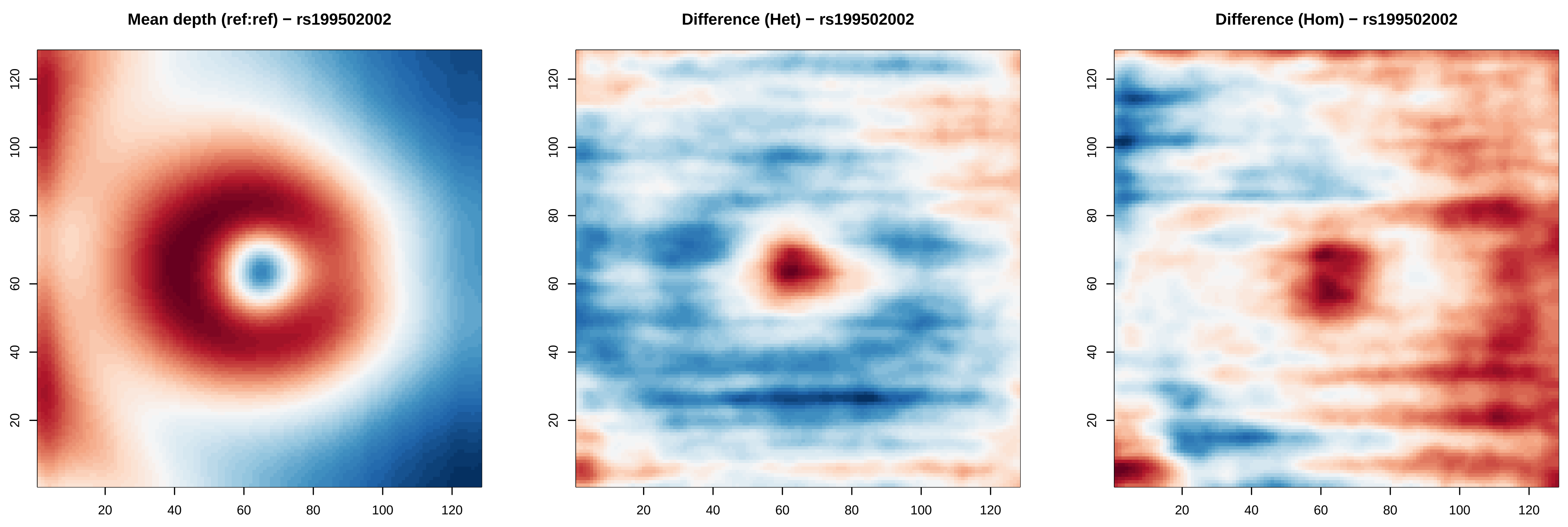
Mean depth (ref:ref) − rs199502002
Difference (Het) − rs199502002
Difference (Hom) − rs199502002
20
40
60
80
100
120

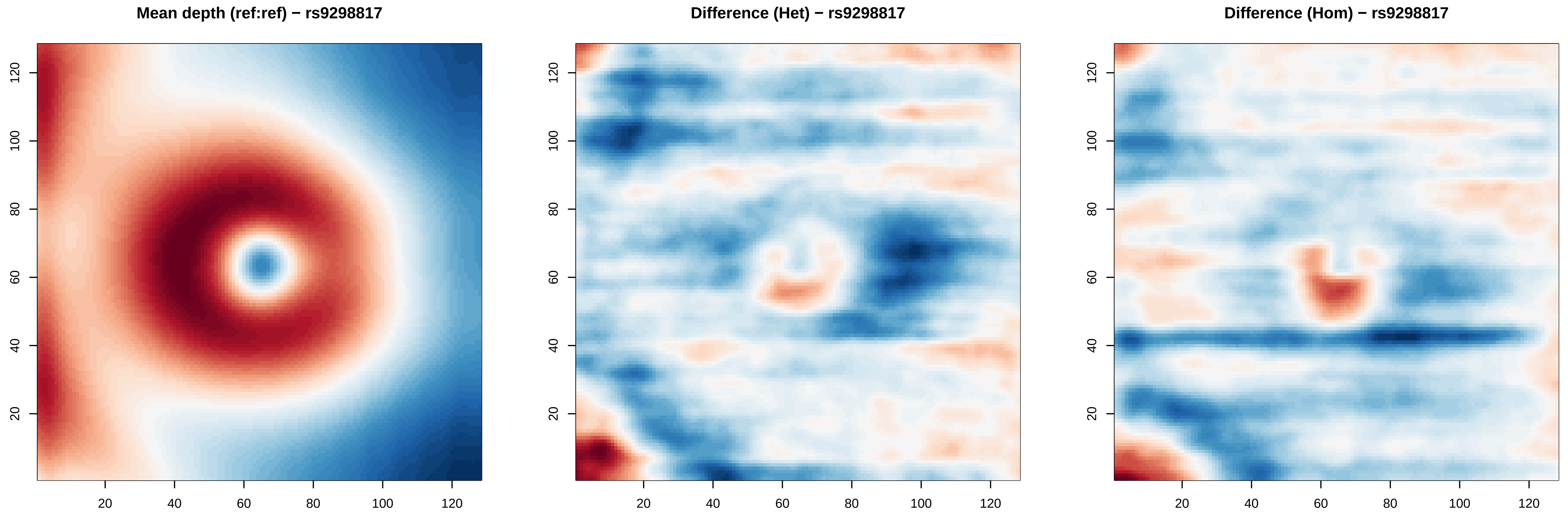

Mean depth (ref:ref) − rs9298817
Difference (Het) − rs9298817
Difference (Hom) − rs9298817
20
40
60
80
100
120

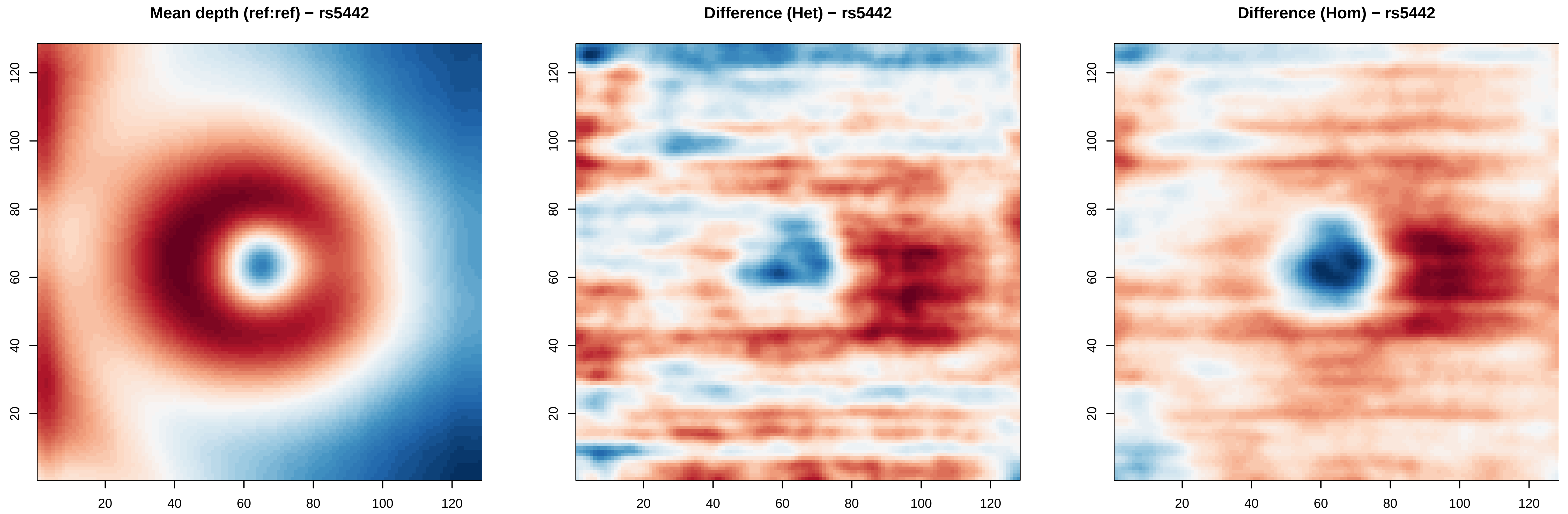
Mean depth (ref:ref) − rs5442
Difference (Het) − rs5442
Difference (Hom) − rs5442
20
40
60
80
100
120

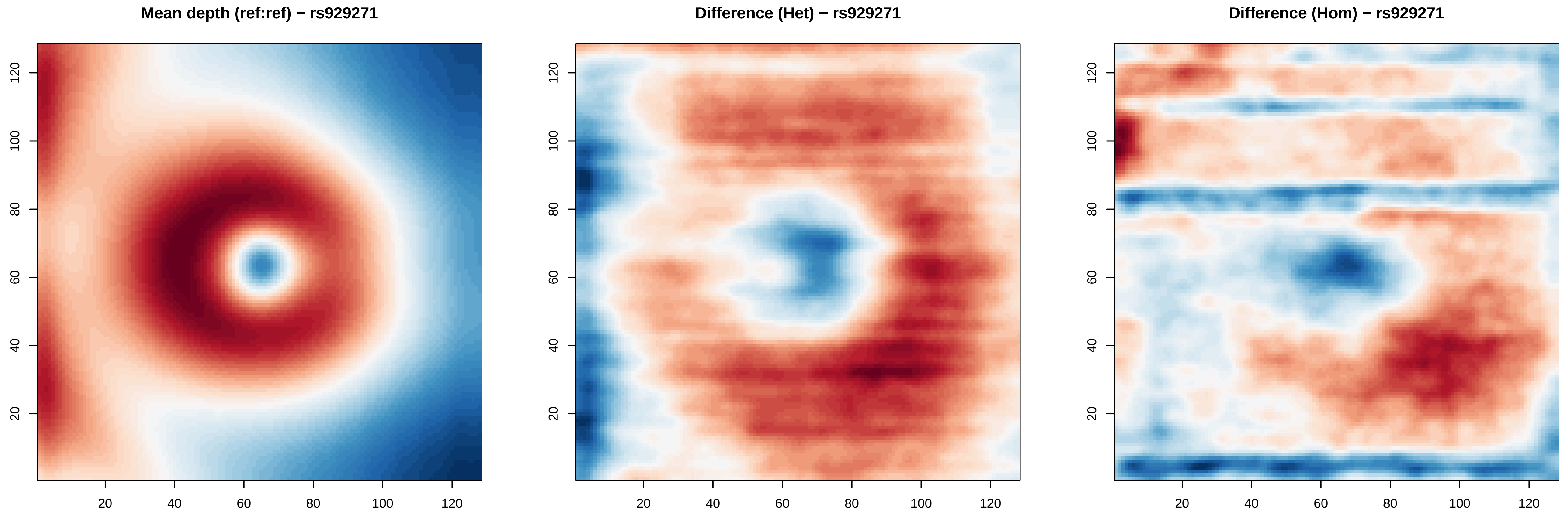
Mean depth (ref:ref) − rs929271
Difference (Het) − rs929271
Difference (Hom) − rs929271
20
40
60
80
100
120

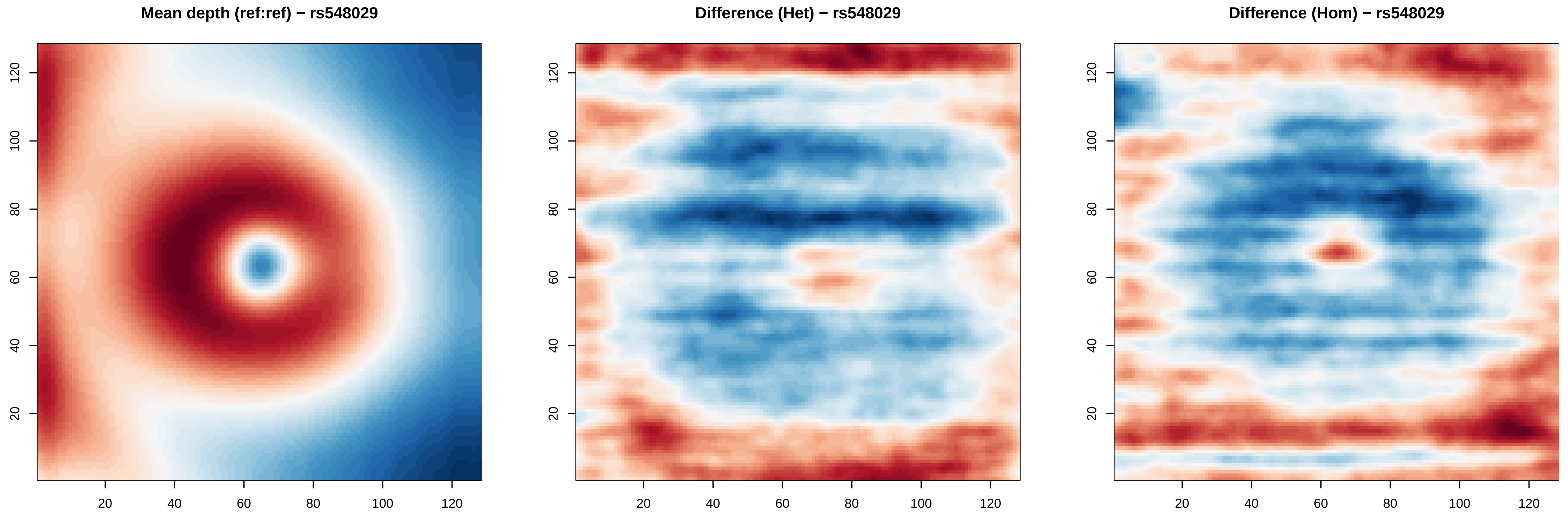
Mean depth (ref:ref) − rs548029
Difference (Het) − rs548029
Difference (Hom) − rs548029
20
40
60
80
100
120

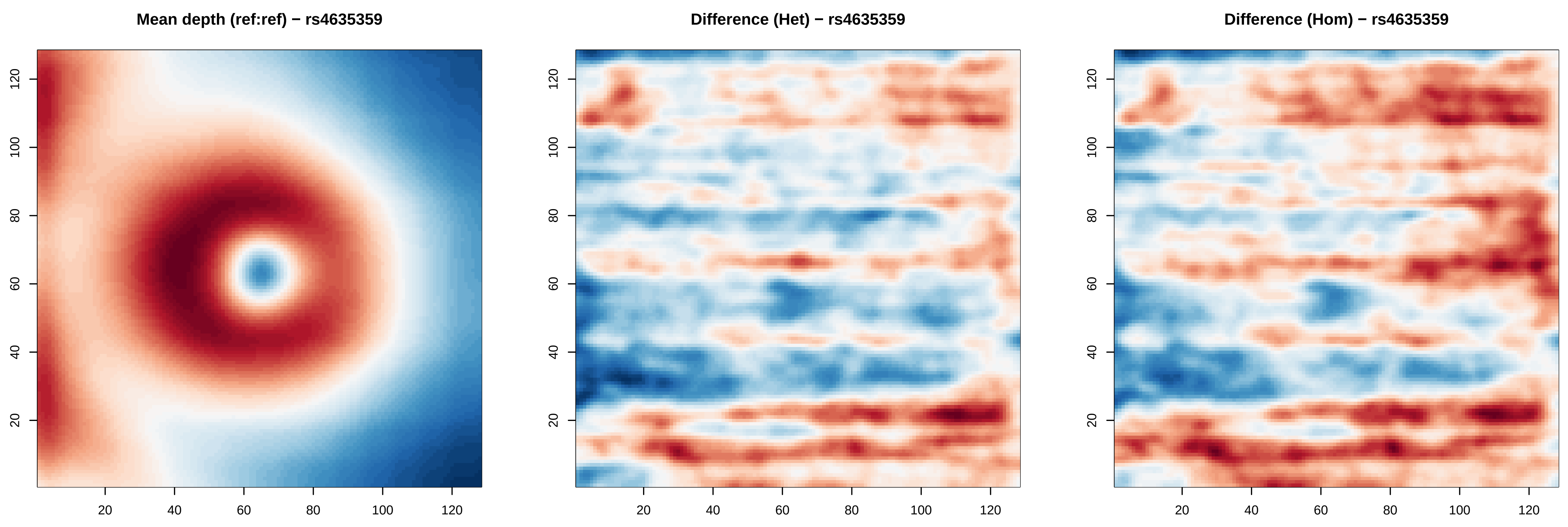
Mean depth (ref:ref) − rs4635359
Difference (Het) − rs4635359
Difference (Hom) − rs4635359
20
40
60
80
100
120

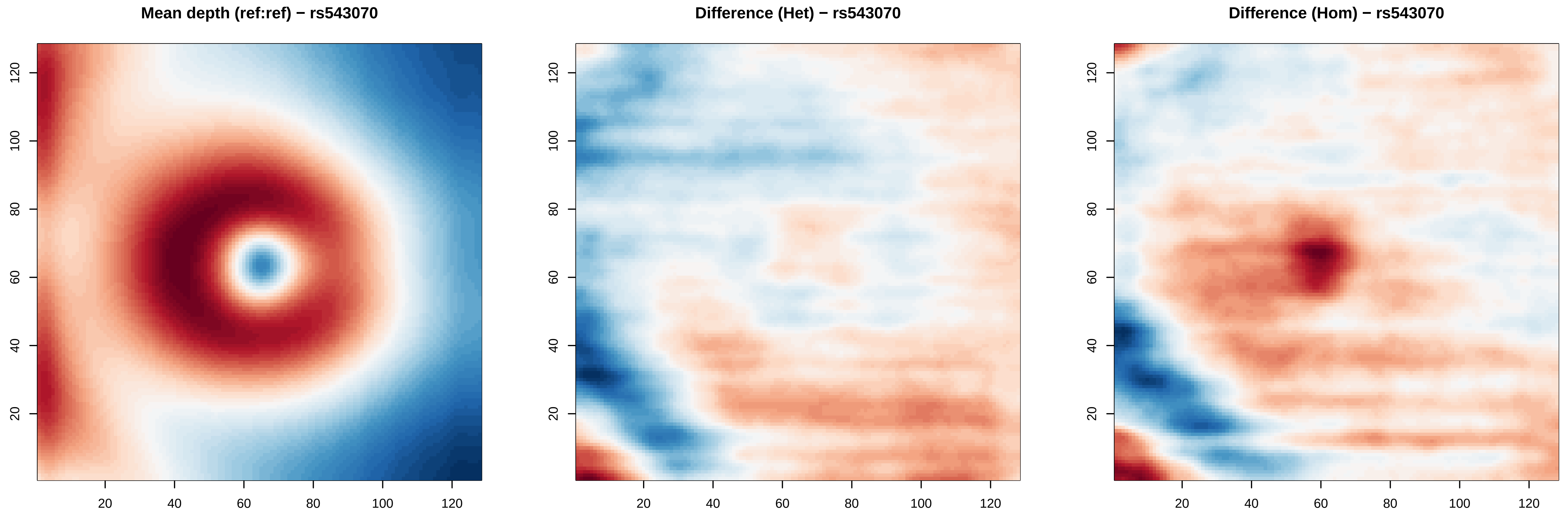
Mean depth (ref:ref) − rs543070
Difference (Het) − rs543070
Difference (Hom) − rs543070
20
40
60
80
100
120

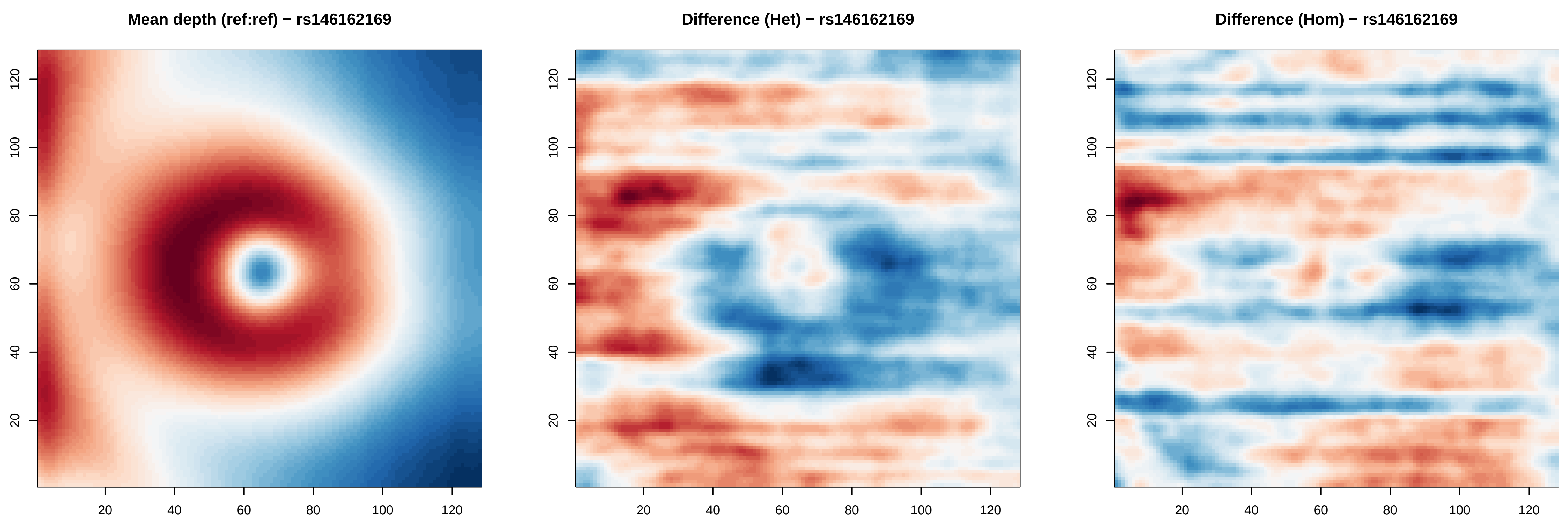

Mean depth (ref:ref) − rs146162169
Difference (Het) − rs146162169
Difference (Hom) − rs146162169
20
40
60
80
100
120

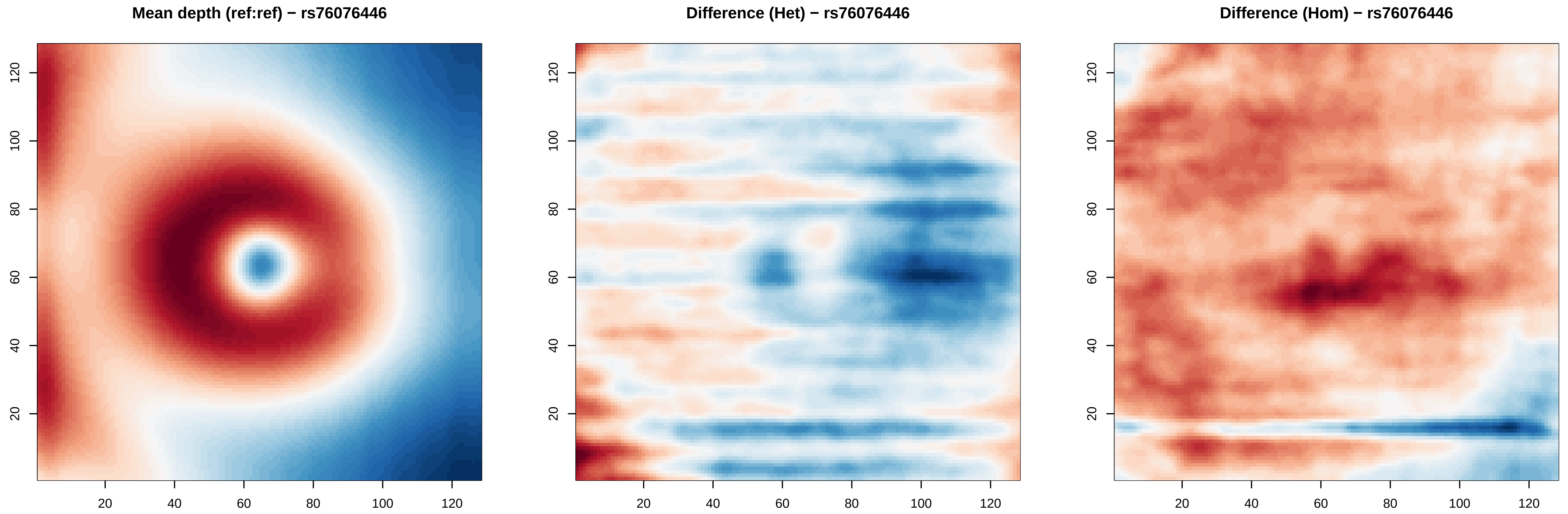

Mean depth (ref:ref) − rs76076446
Difference (Het) − rs76076446
Difference (Hom) − rs76076446
20
40
60
80
100
120

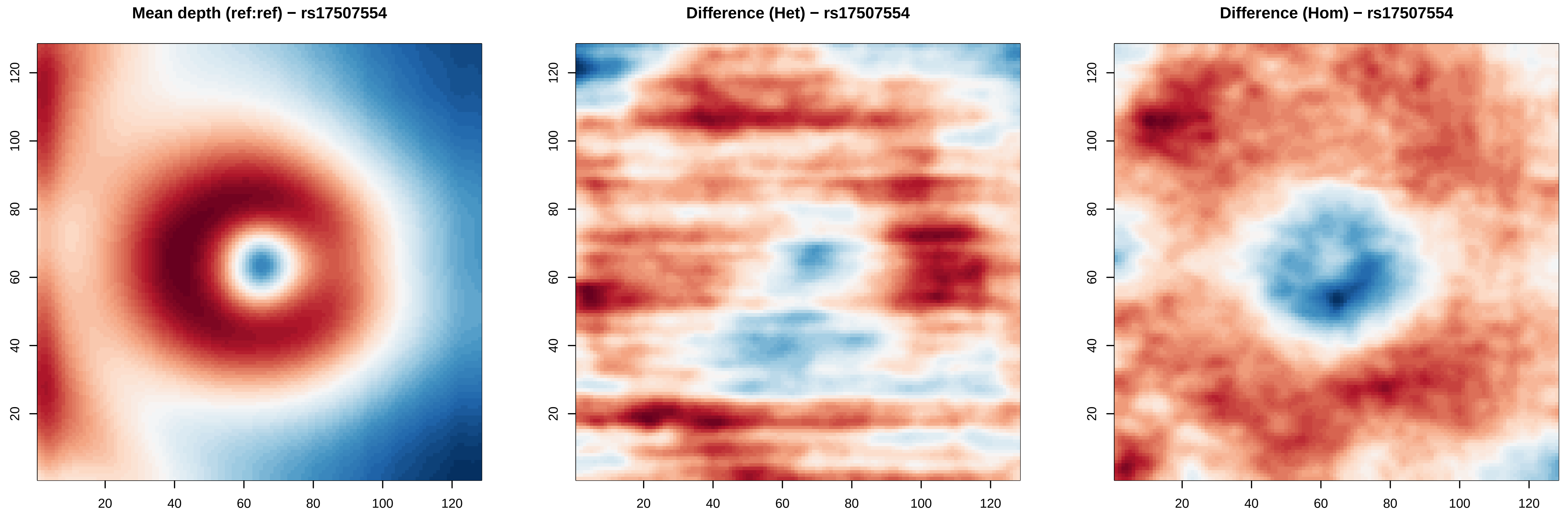
Mean depth (ref:ref) − rs17507554
Difference (Het) − rs17507554
Difference (Hom) − rs17507554
20
40
60
80
100
120

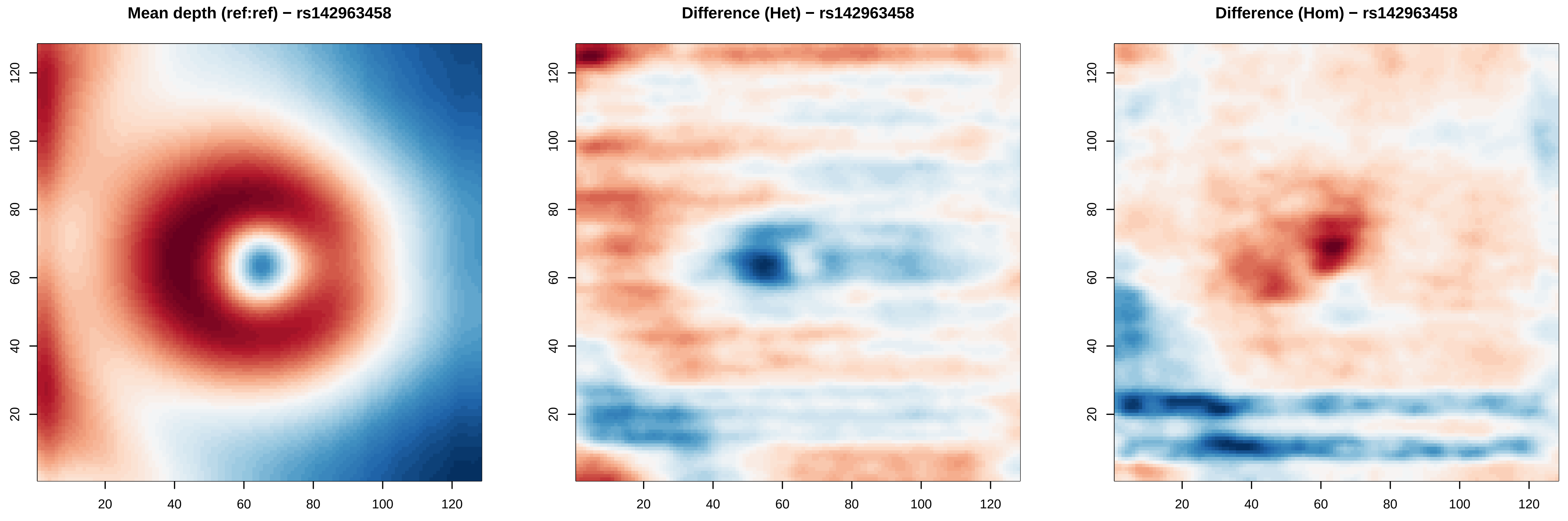
Mean depth (ref:ref) − rs142963458
Difference (Het) − rs142963458
Difference (Hom) − rs142963458
20
40
60
80
100
120

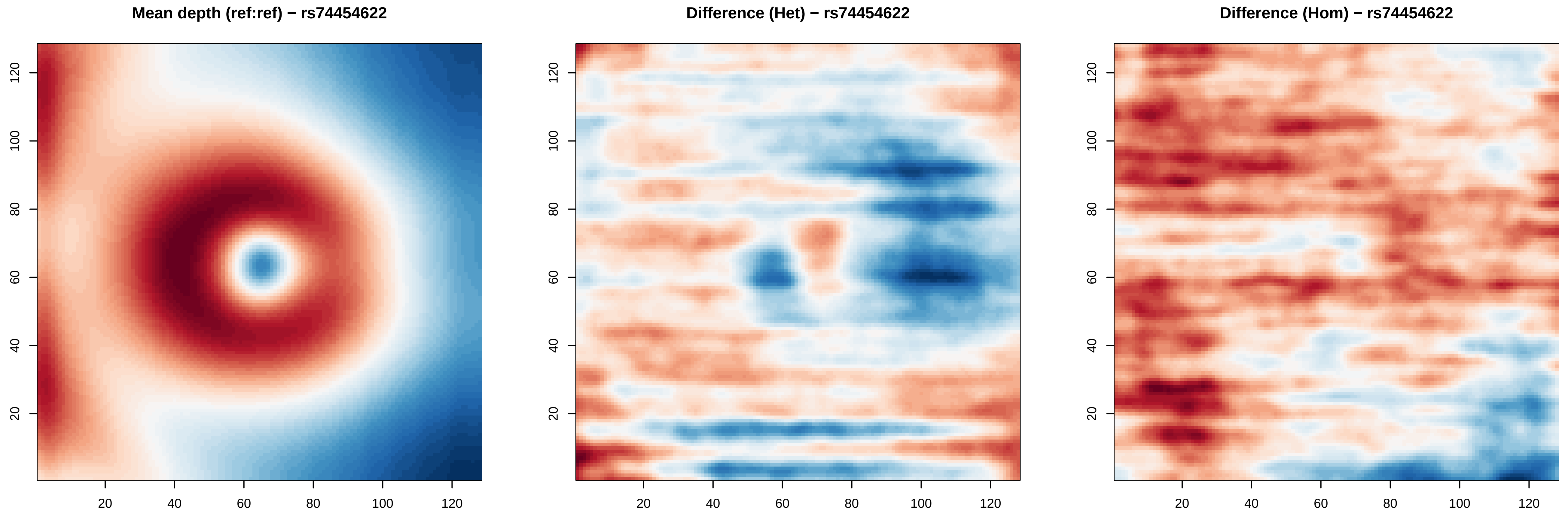

Mean depth (ref:ref) − rs74454622
Difference (Het) − rs74454622
Difference (Hom) − rs74454622
20
40
60
80
100
120

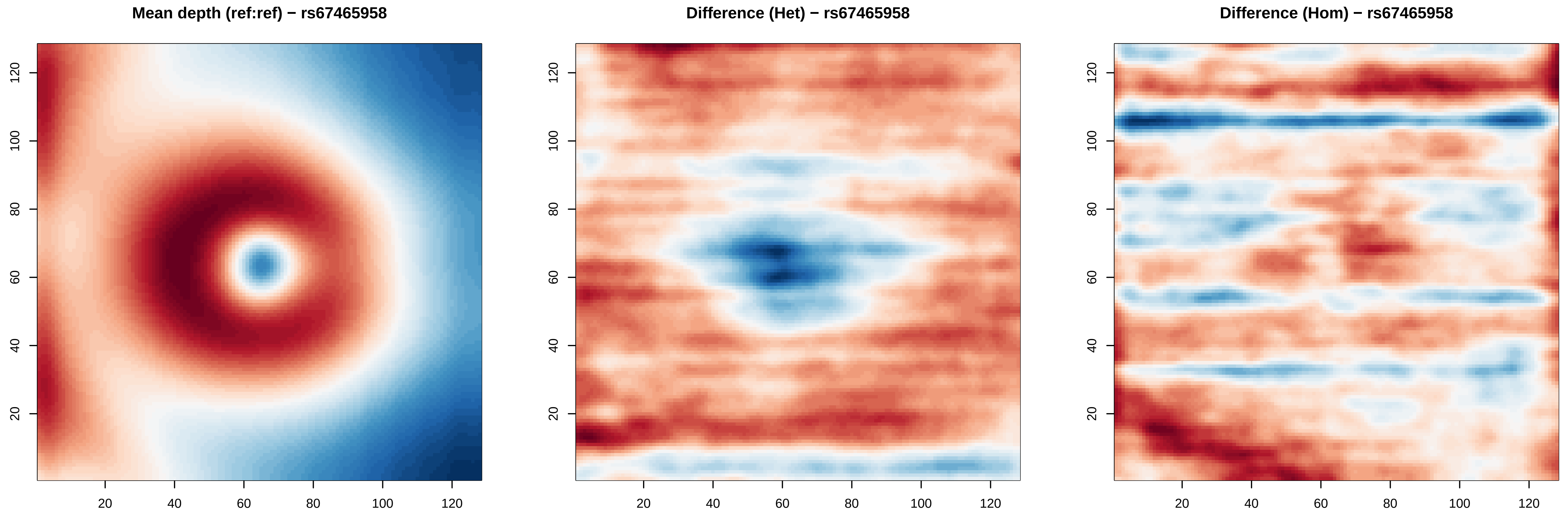

Mean depth (ref:ref) − rs67465958
Difference (Het) − rs67465958
Difference (Hom) − rs67465958
20
40
60
80
100
120

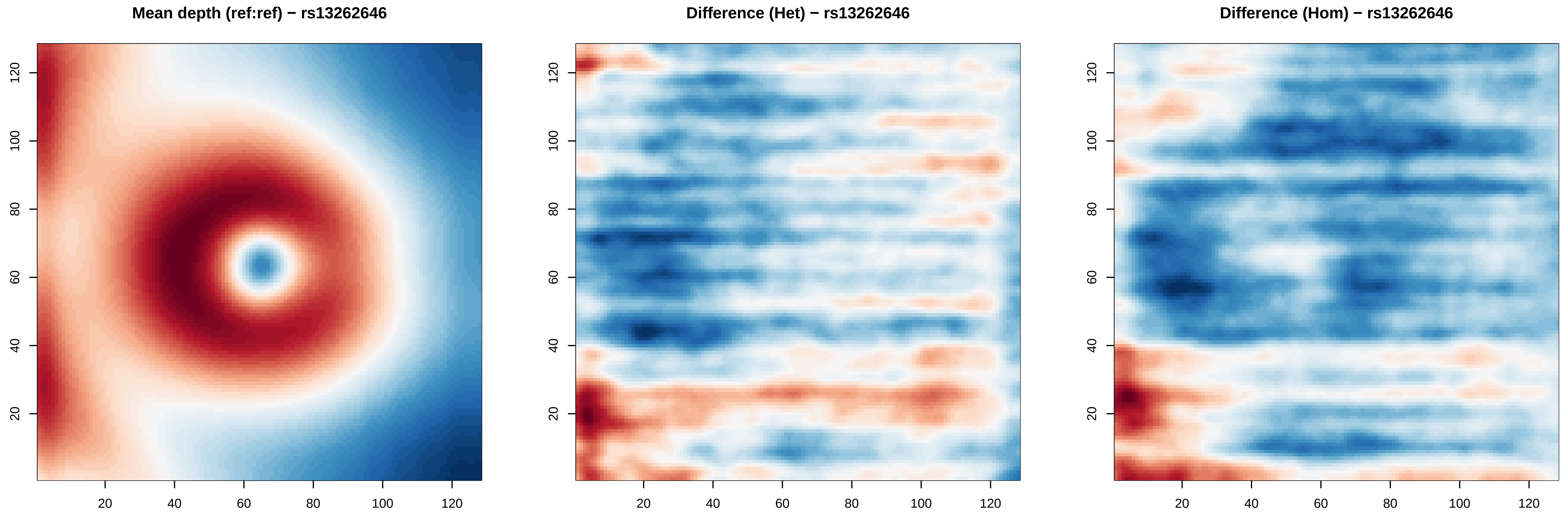

Mean depth (ref:ref) − rs13262646
Difference (Het) − rs13262646
Difference (Hom) − rs13262646
20
40
60
80
100
120

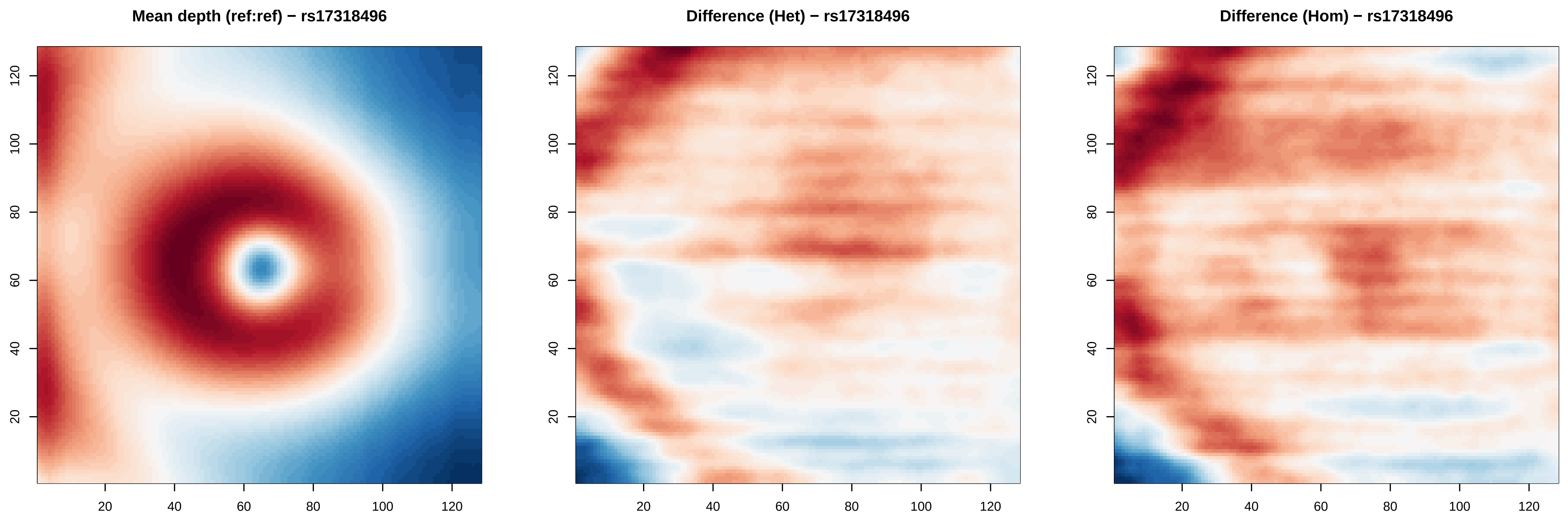

Mean depth (ref:ref) − rs17318496
Difference (Het) − rs17318496
Difference (Hom) − rs17318496
20
40
60
80
100
120

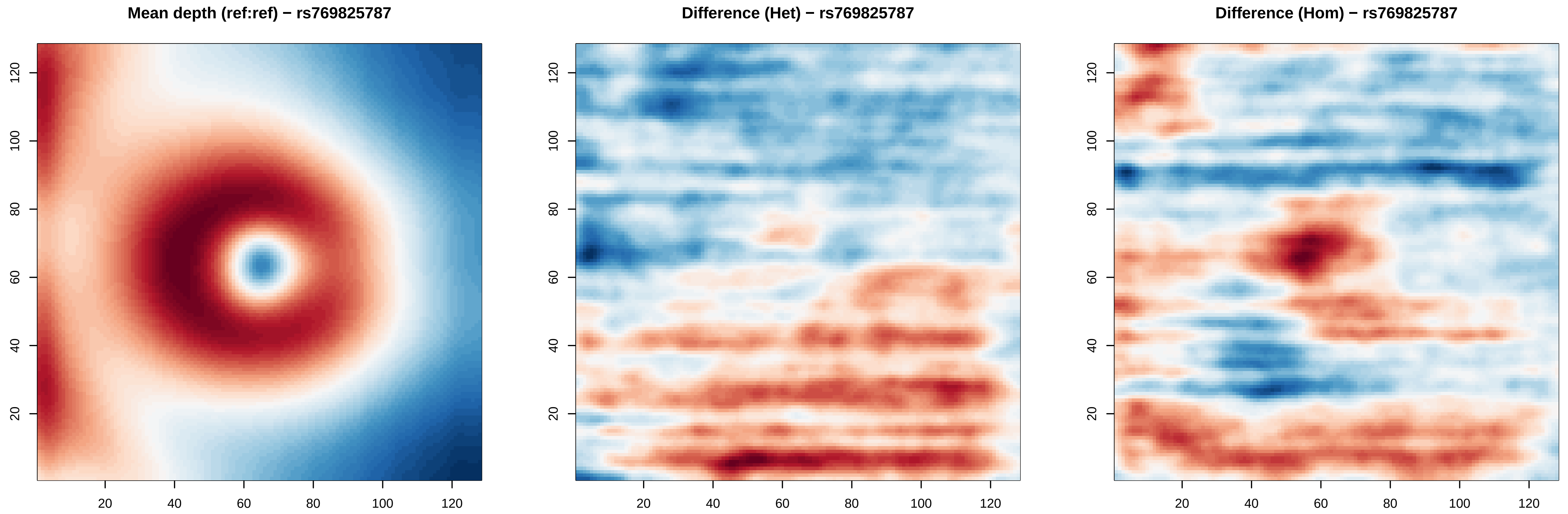

Mean depth (ref:ref) − rs769825787
Difference (Het) − rs769825787
Difference (Hom) − rs769825787
20
40
60
80
100
120

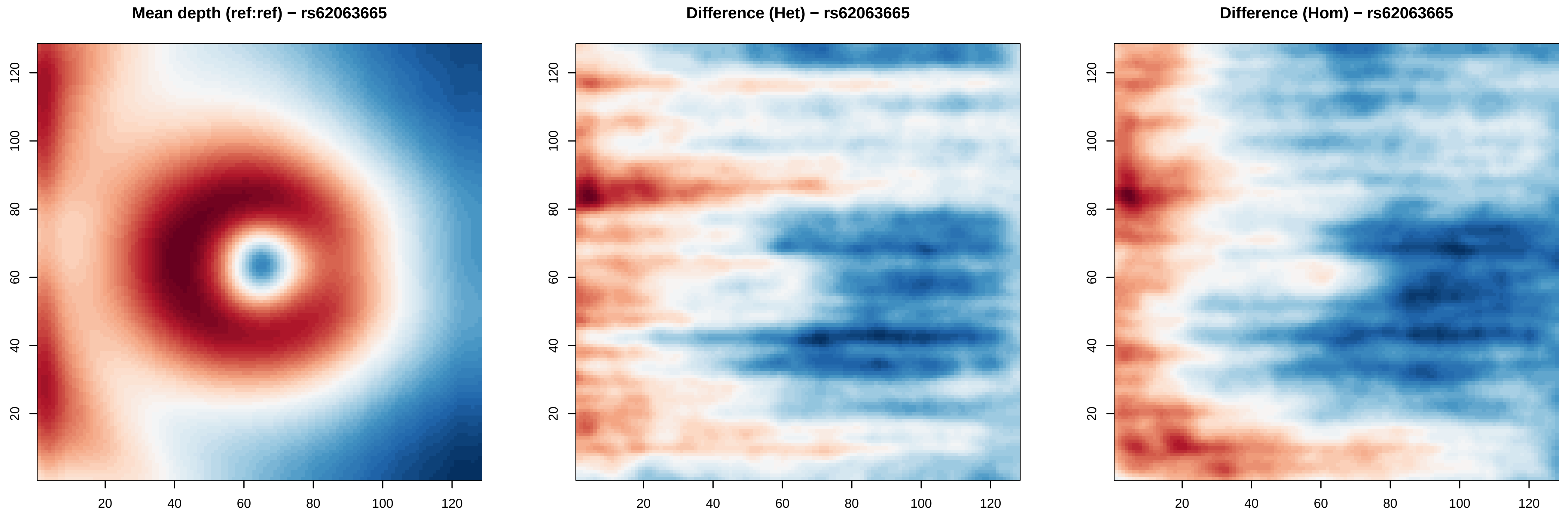

Mean depth (ref:ref) − rs62063665
Difference (Het) − rs62063665
Difference (Hom) − rs62063665
20
40
60
80
100
120

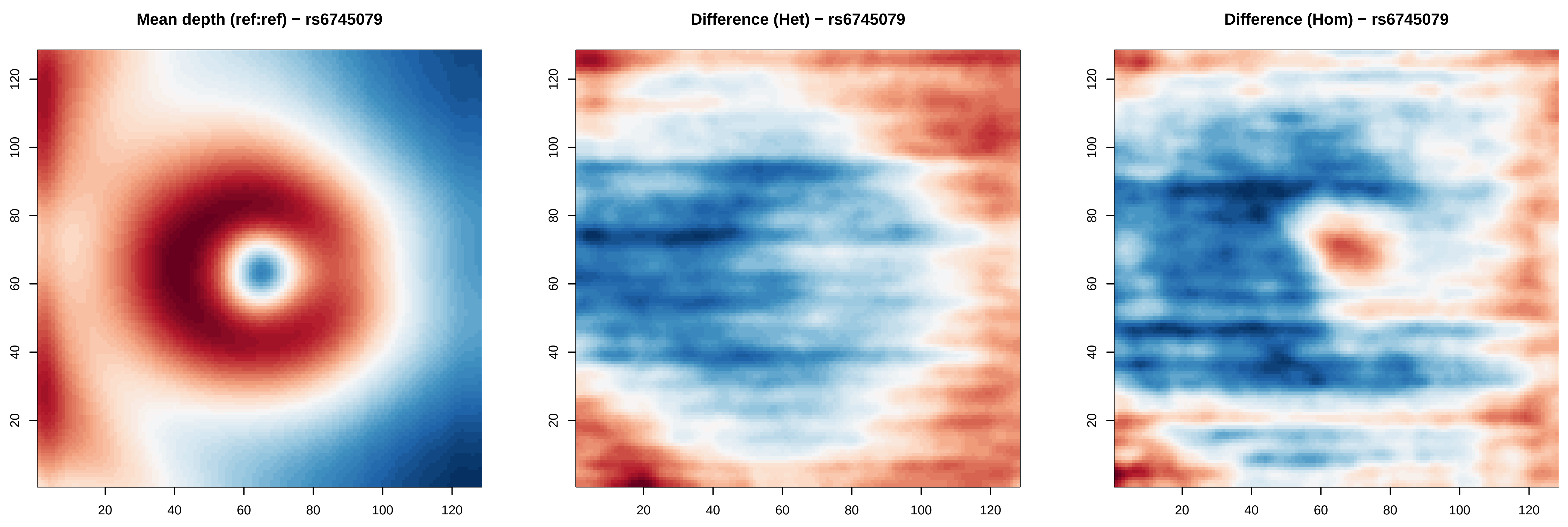

Mean depth (ref:ref) − rs6745079
Difference (Het) − rs6745079
Difference (Hom) − rs6745079
20
40
60
80
100
120

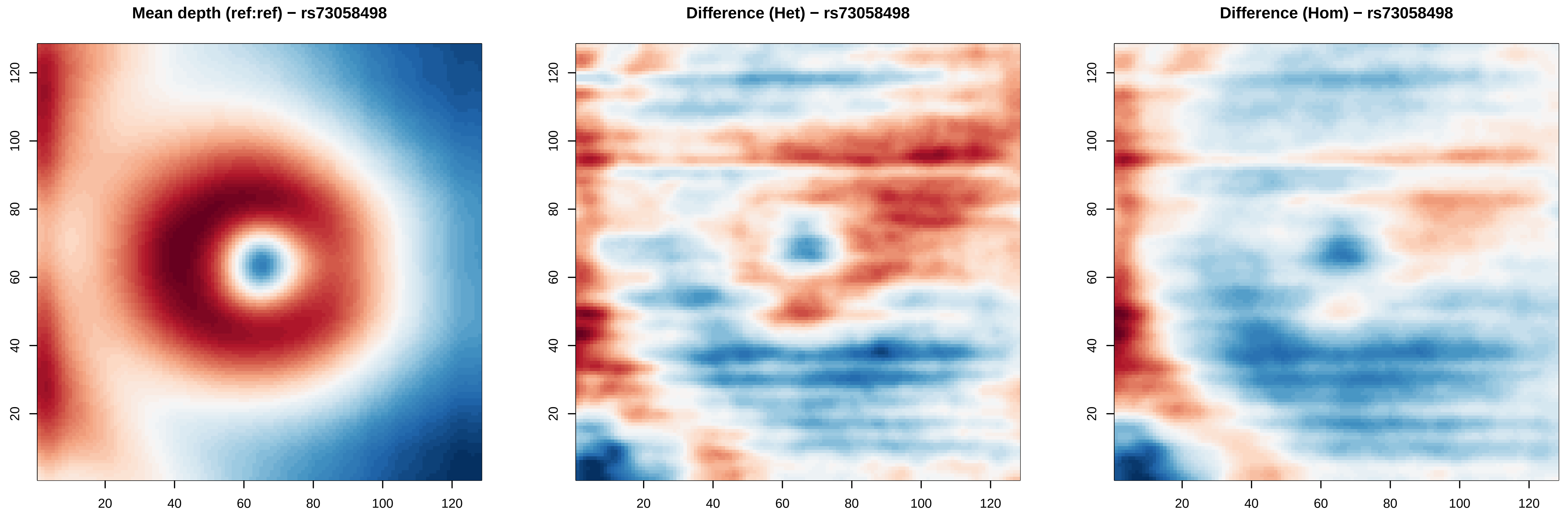
Mean depth (ref:ref) − rs73058498
Difference (Het) − rs73058498
Difference (Hom) − rs73058498
20
40
60
80
100
120

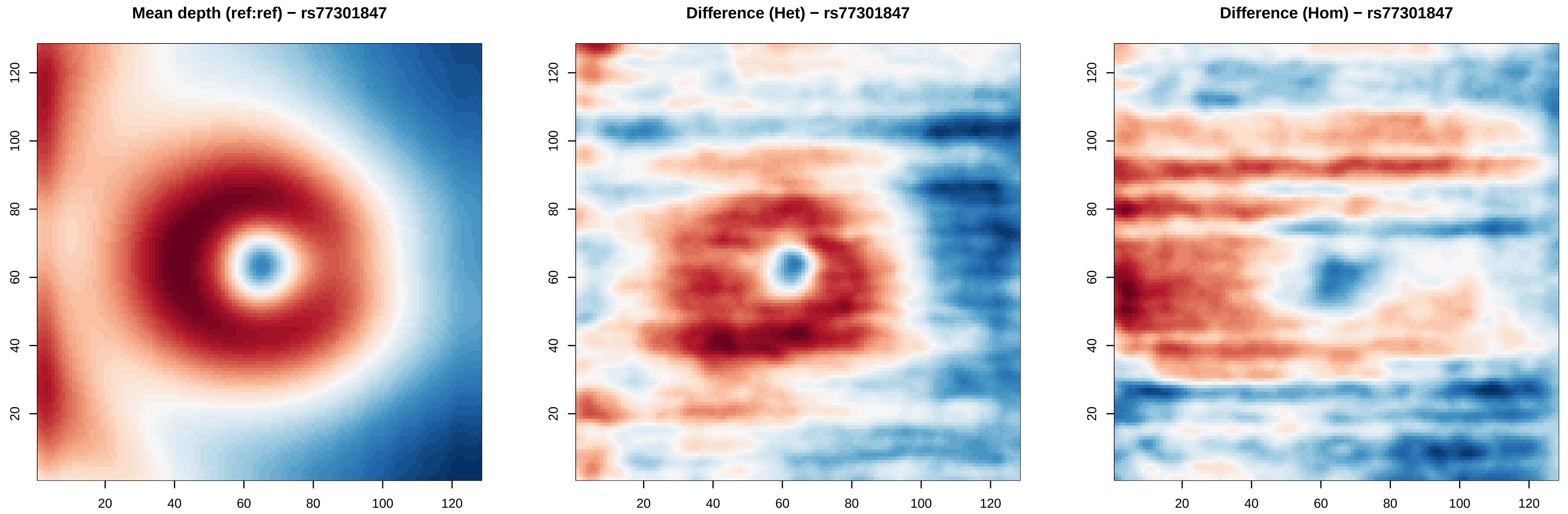
Mean depth (ref:ref) − rs77301847
Difference (Het) − rs77301847
Difference (Hom) − rs77301847
20
40
60
80
100
120

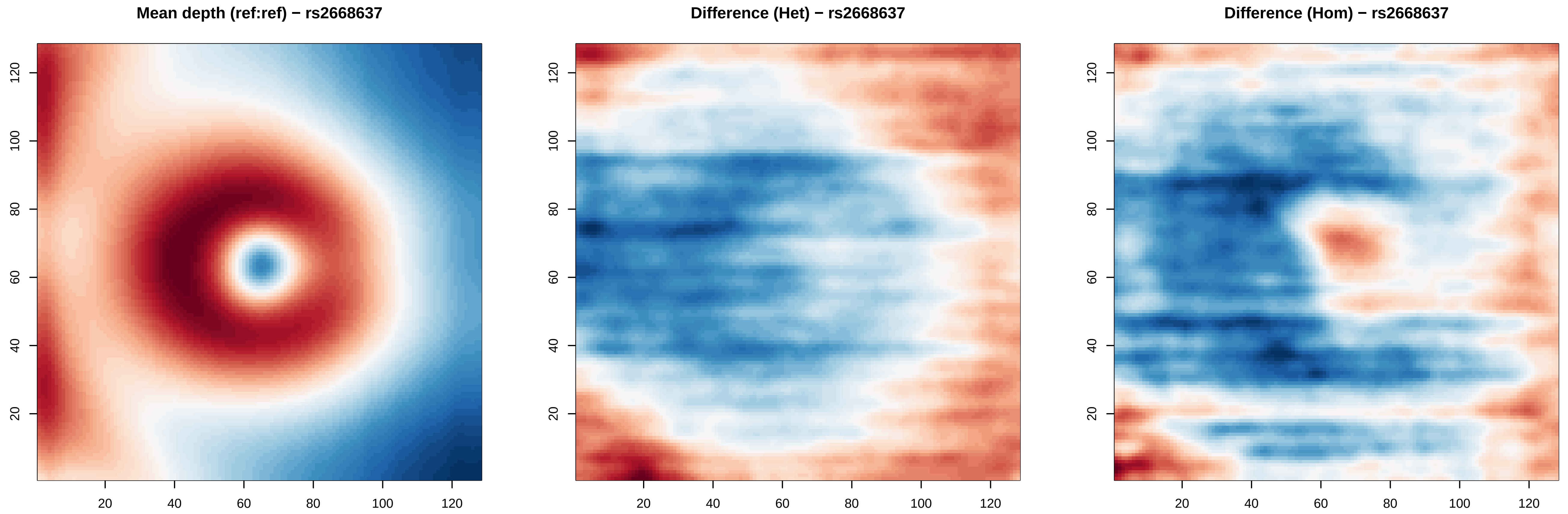
Mean depth (ref:ref) − rs2668637
Difference (Het) − rs2668637
Difference (Hom) − rs2668637
20
40
60
80
100
120

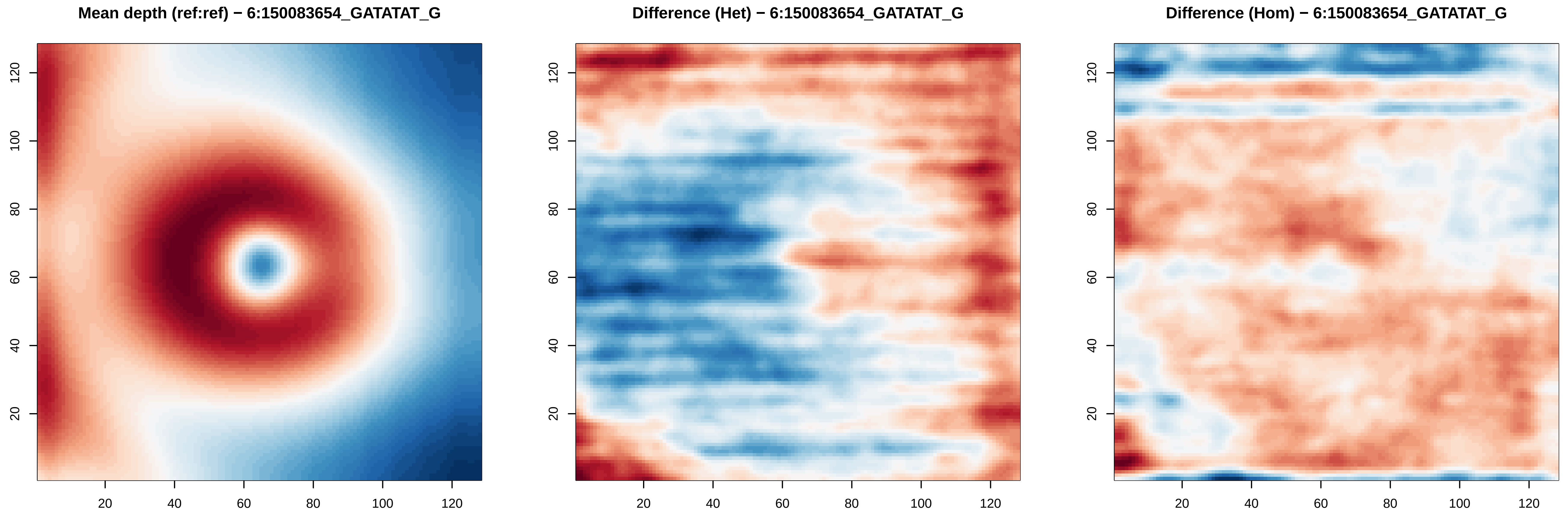
Mean depth (ref:ref) − 6:150083654_GATATAT_G
Difference (Het) − 6:150083654_GATATAT_G
Difference (Hom) − 6:150083654_GATATAT_G
20 40 60 80 100 120
20 40 60 80 100 120

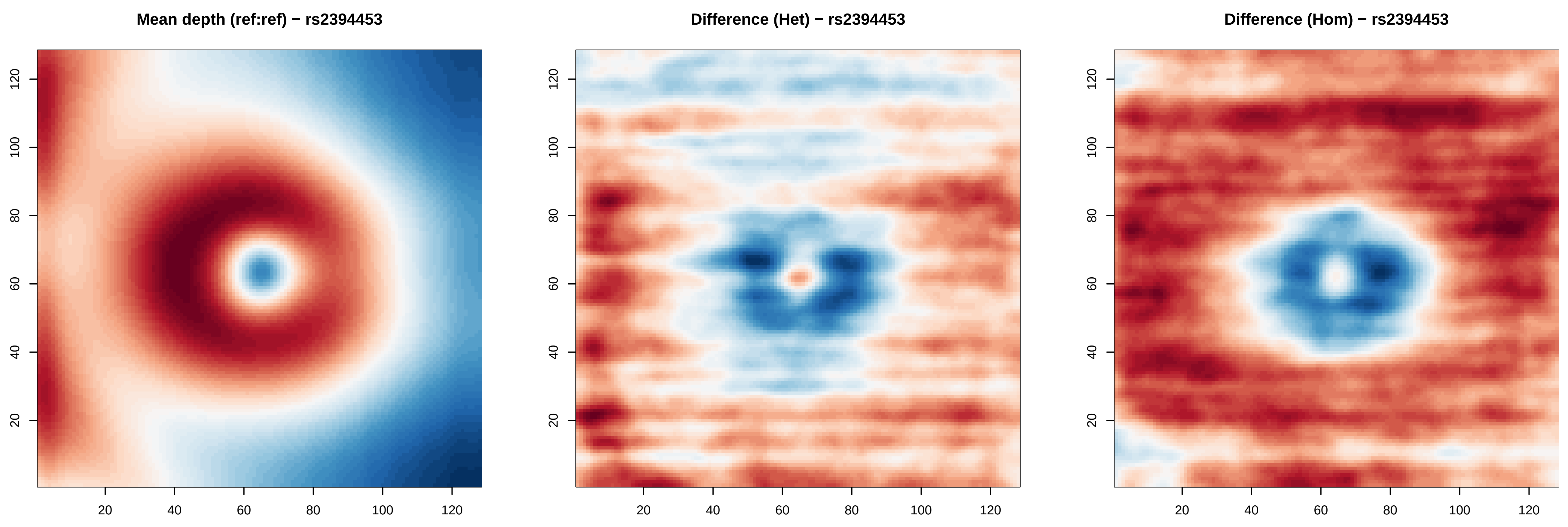

Mean depth (ref:ref) − rs2394453
Difference (Het) − rs2394453
Difference (Hom) − rs2394453
20
40
60
80
100
120

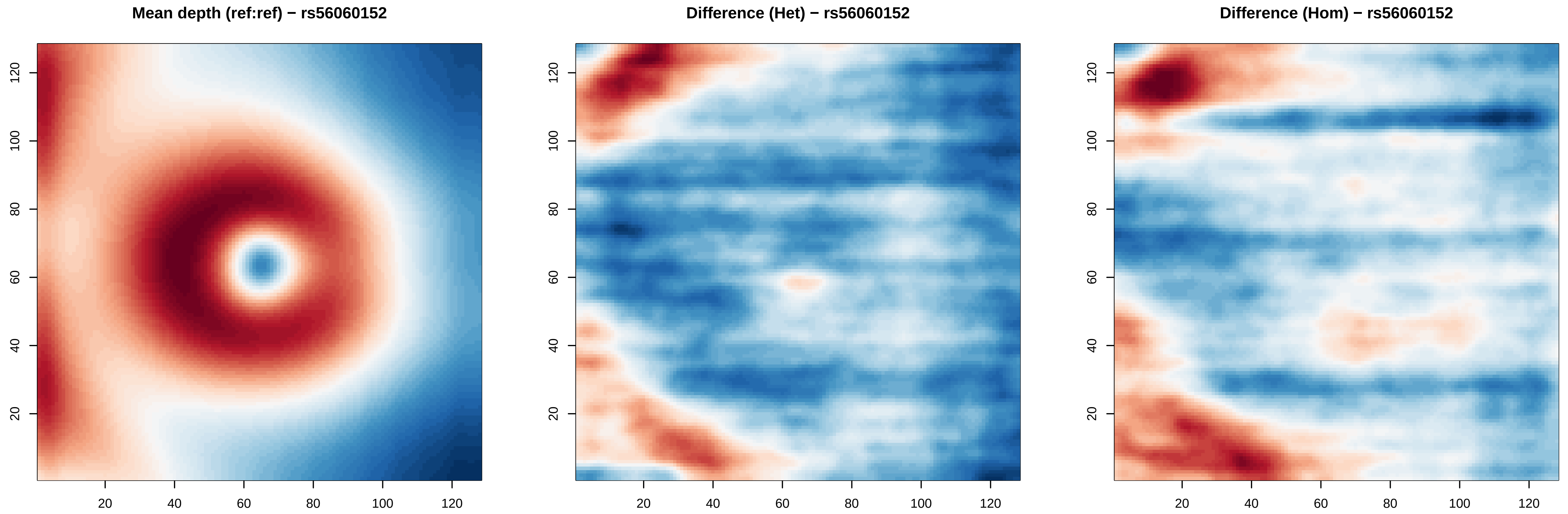

Mean depth (ref:ref) − rs56060152
Difference (Het) − rs56060152
Difference (Hom) − rs56060152
20
40
60
80
100
120

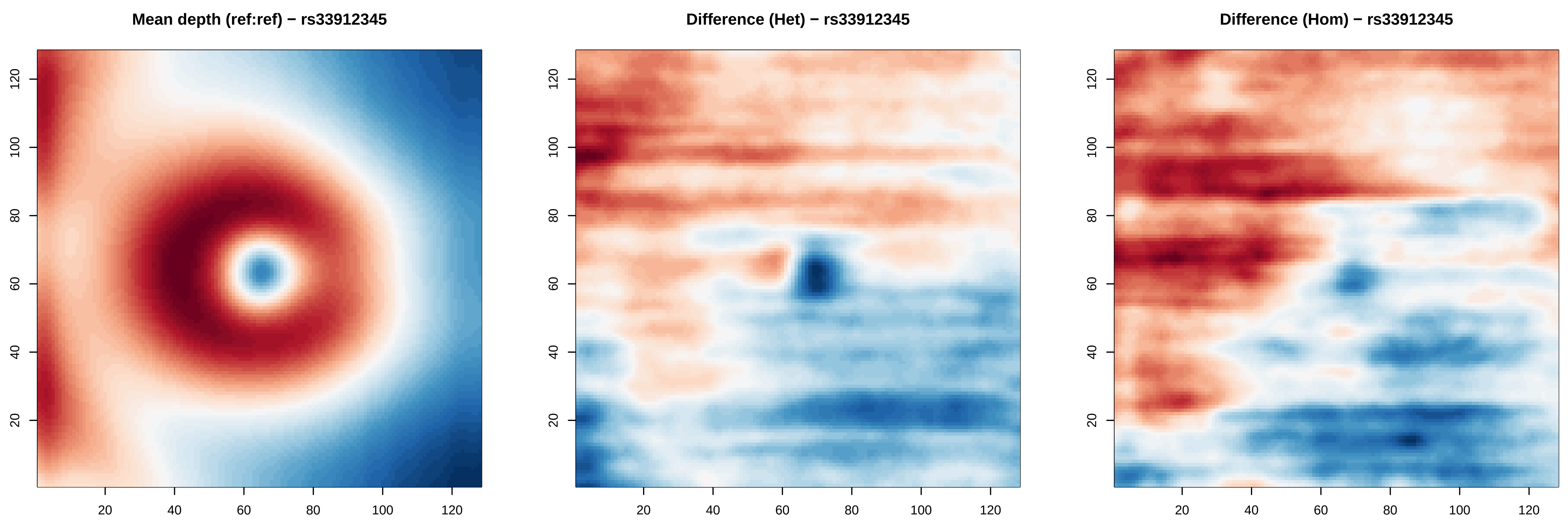

Mean depth (ref:ref) − rs33912345
Difference (Het) − rs33912345
Difference (Hom) − rs33912345
20
40
60
80
100
120

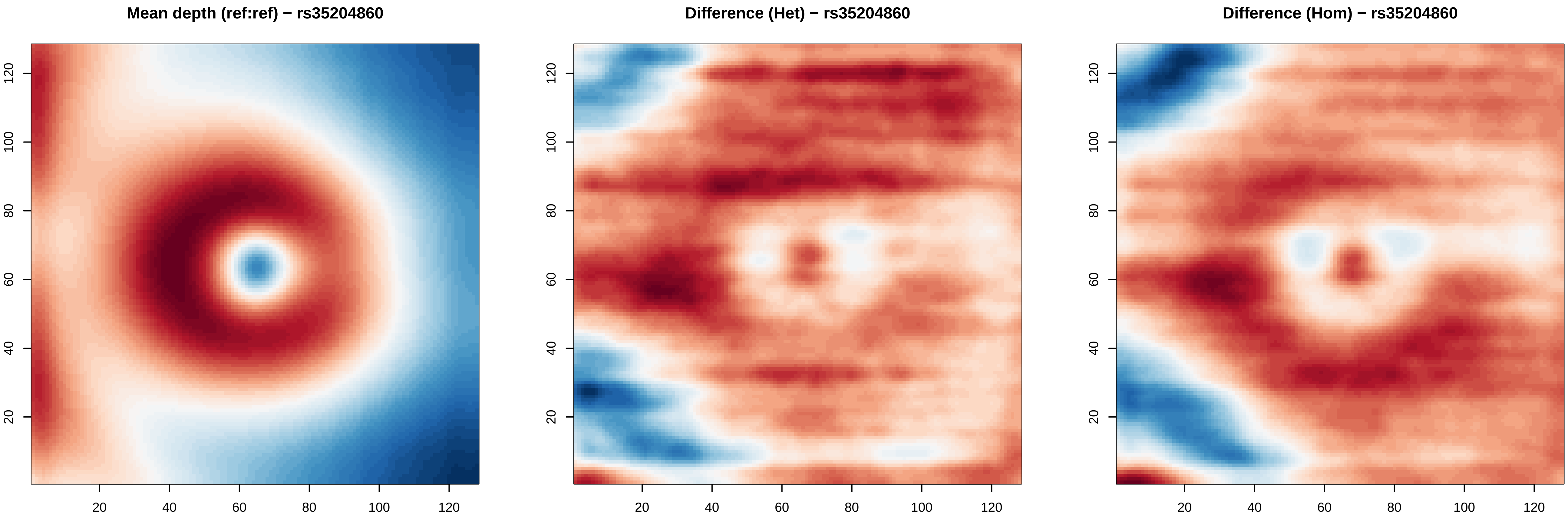

Mean depth (ref:ref) − rs35204860
Difference (Het) − rs35204860
Difference (Hom) − rs35204860
20
40
60
80
100
120

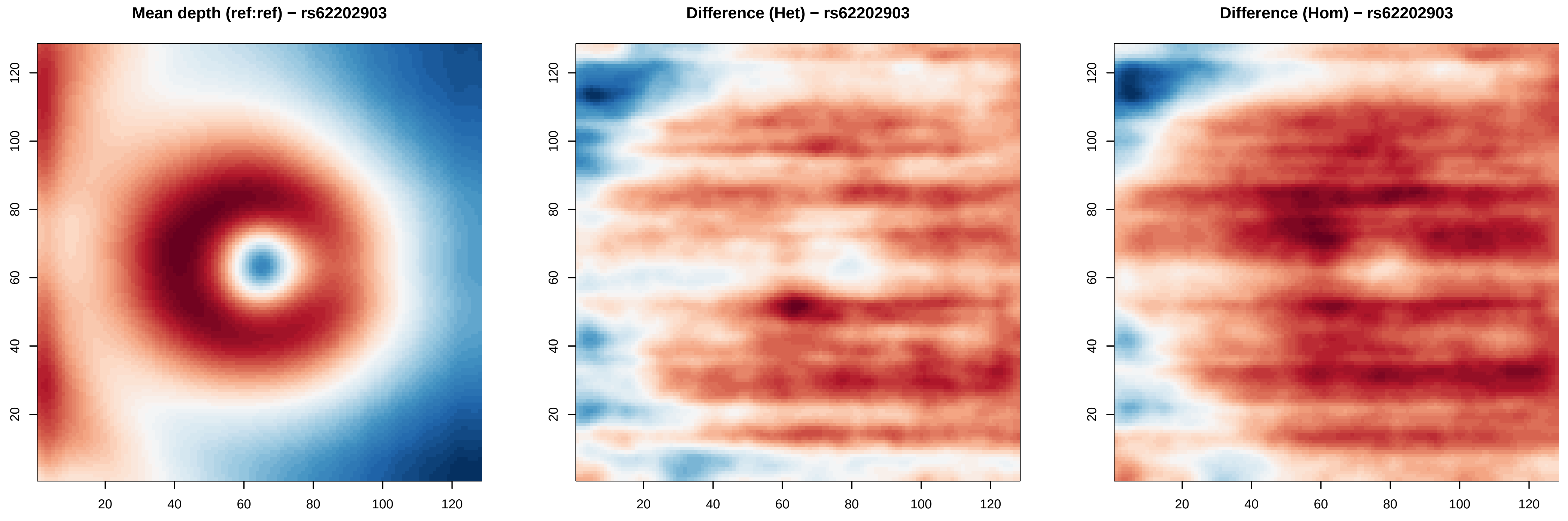

Mean depth (ref:ref) − rs62202903
Difference (Het) − rs62202903
Difference (Hom) − rs62202903
20
40
60
80
100
120

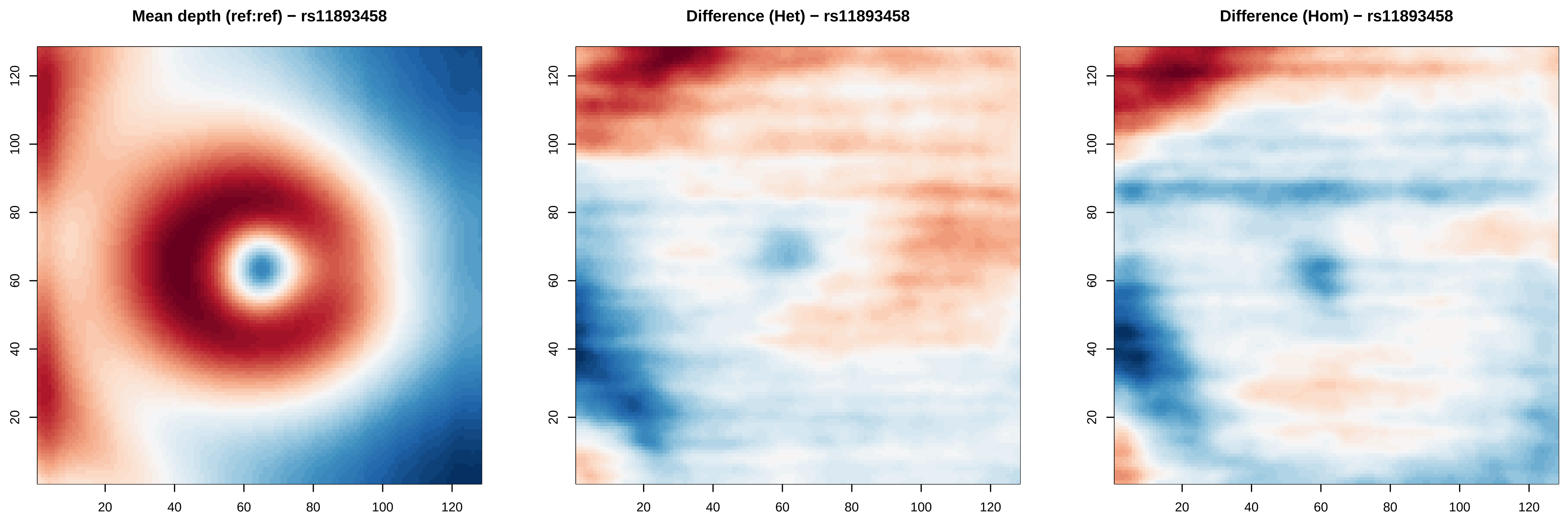

Mean depth (ref:ref) − rs11893458
Difference (Het) − rs11893458
Difference (Hom) − rs11893458
20
40
60
80
100
120

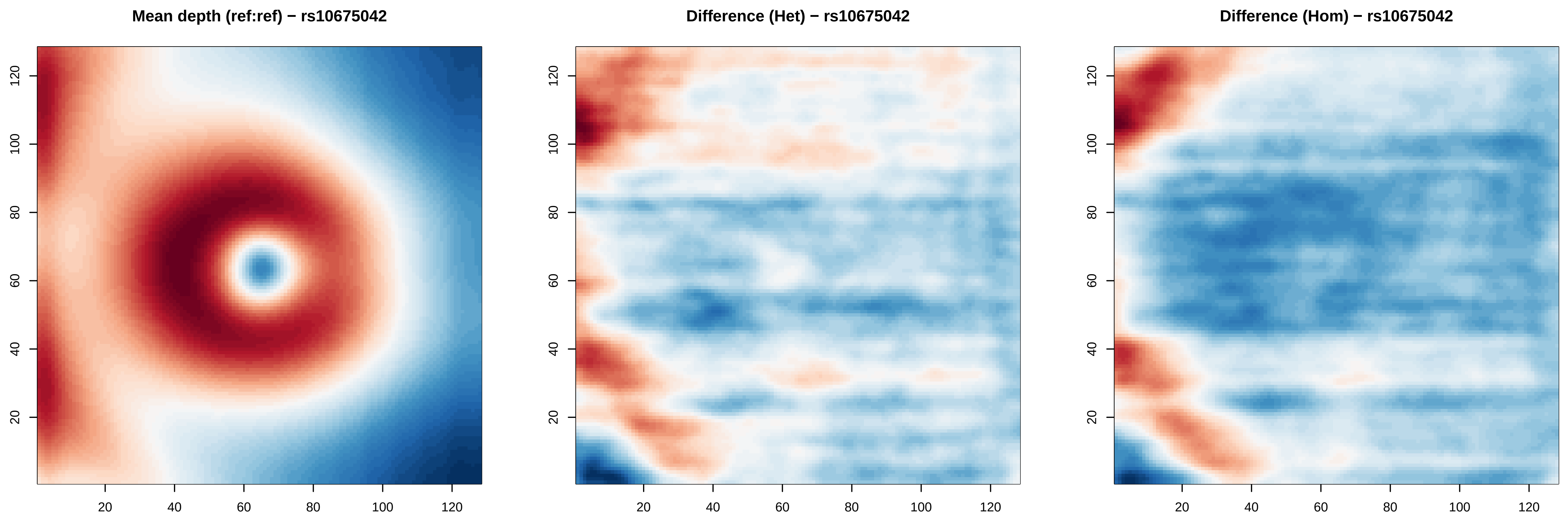
Mean depth (ref:ref) − rs10675042
Difference (Het) − rs10675042
Difference (Hom) − rs10675042
20
40
60
80
100
120

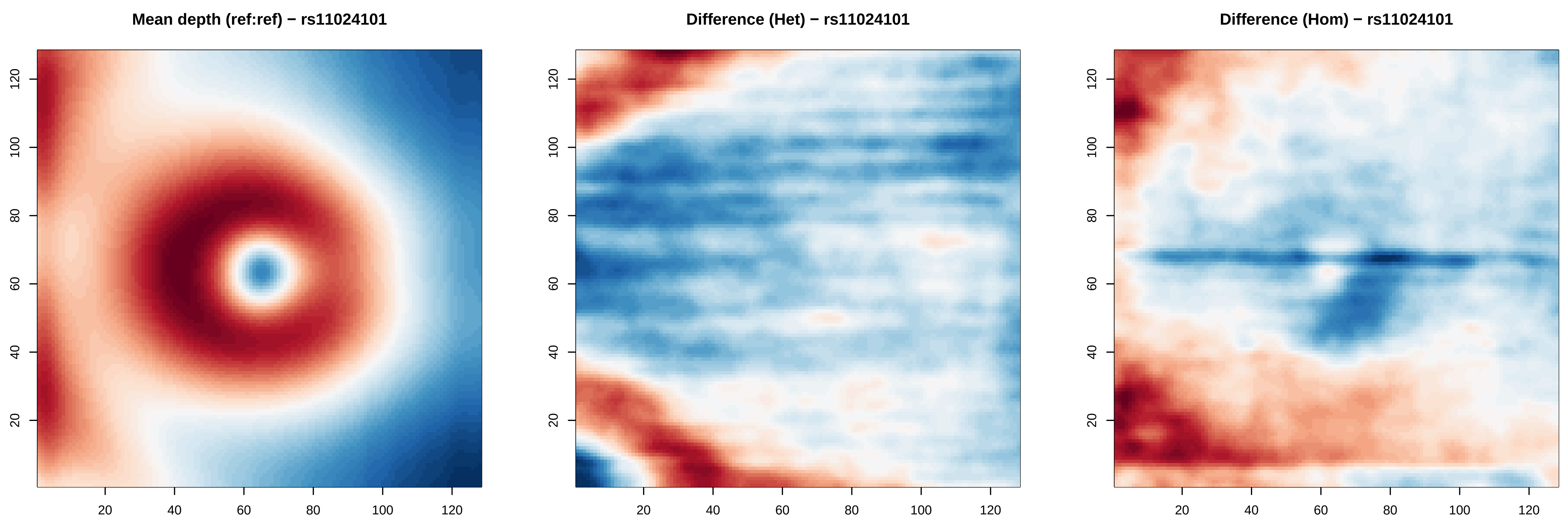
Mean depth (ref:ref) − rs11024101
Difference (Het) − rs11024101
Difference (Hom) − rs11024101
20
40
60
80
100
120

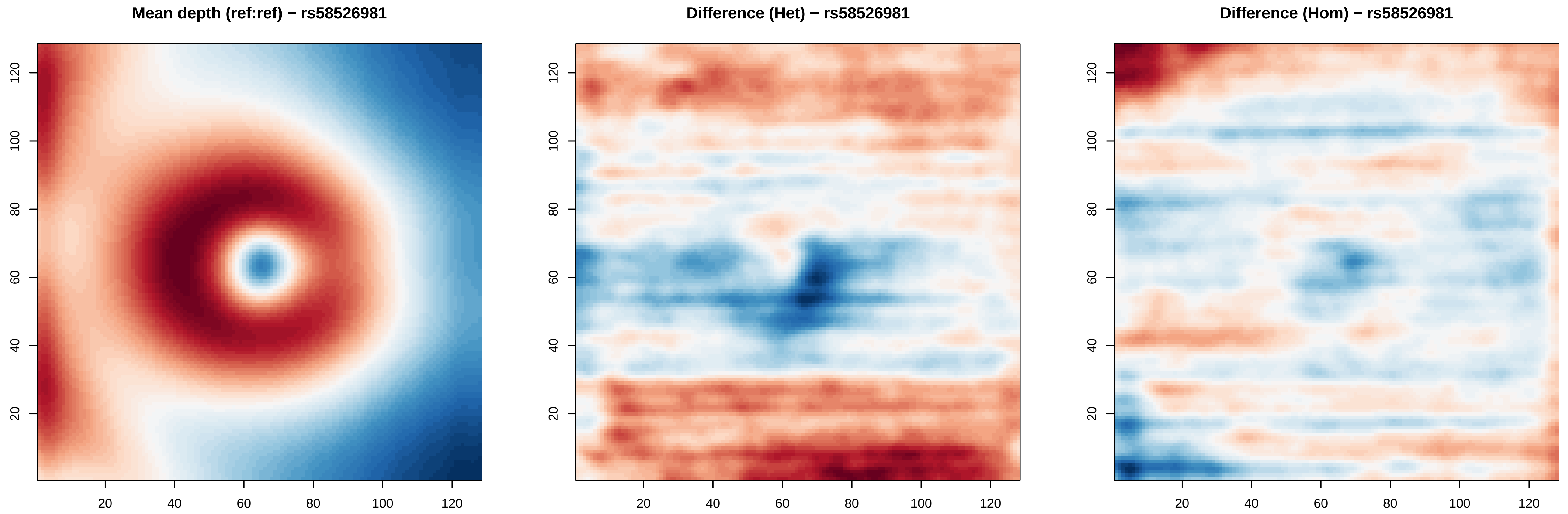

Mean depth (ref:ref) − rs58526981
Difference (Het) − rs58526981
Difference (Hom) − rs58526981
20
40
60
80
100
120

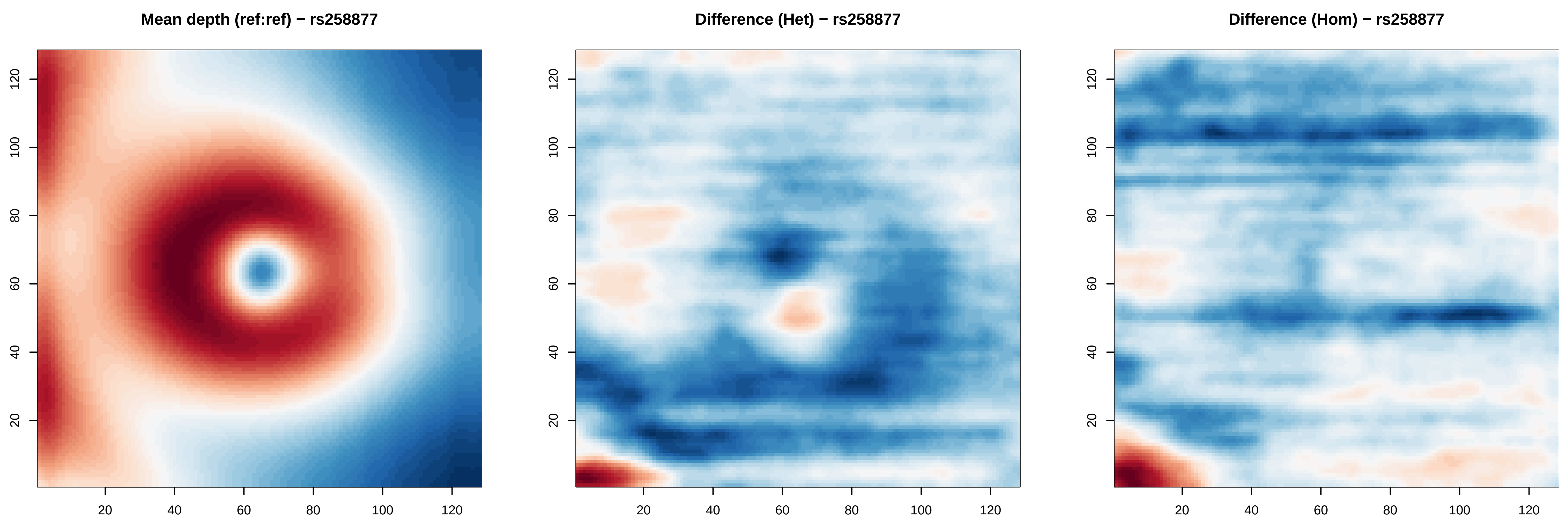
Mean depth (ref:ref) − rs258877
Difference (Het) − rs258877
Difference (Hom) − rs258877
20
40
60
80
100
120

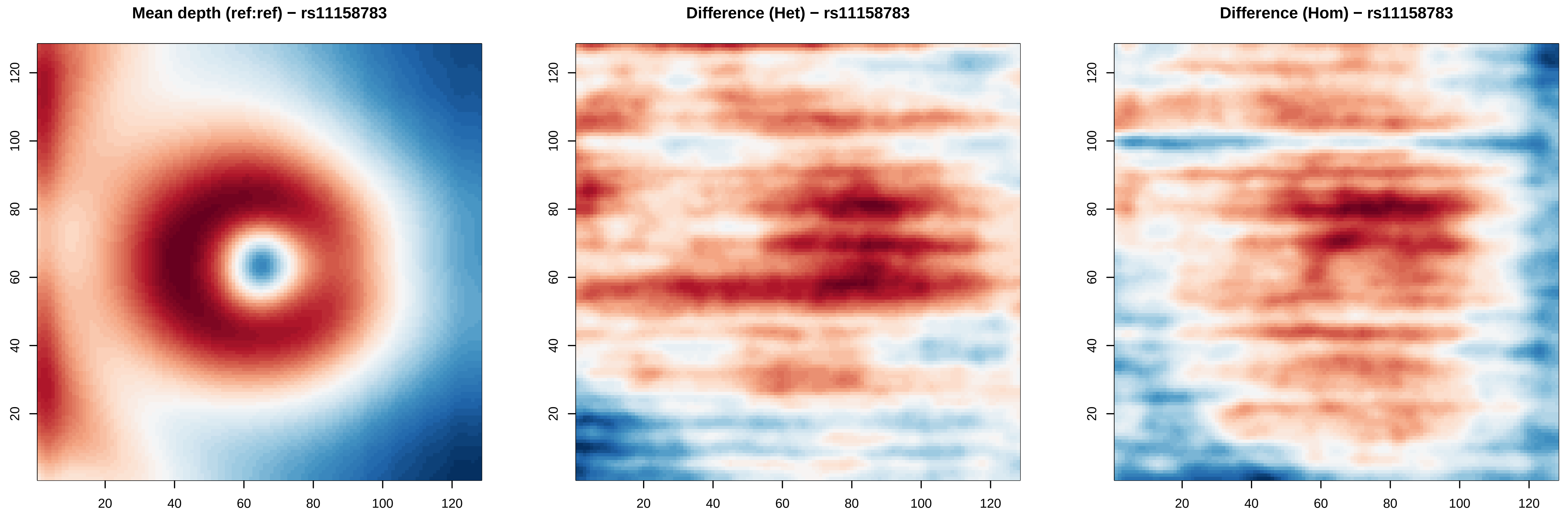

Mean depth (ref:ref) − rs11158783
Difference (Het) − rs11158783
Difference (Hom) − rs11158783
20
40
60
80
100
120

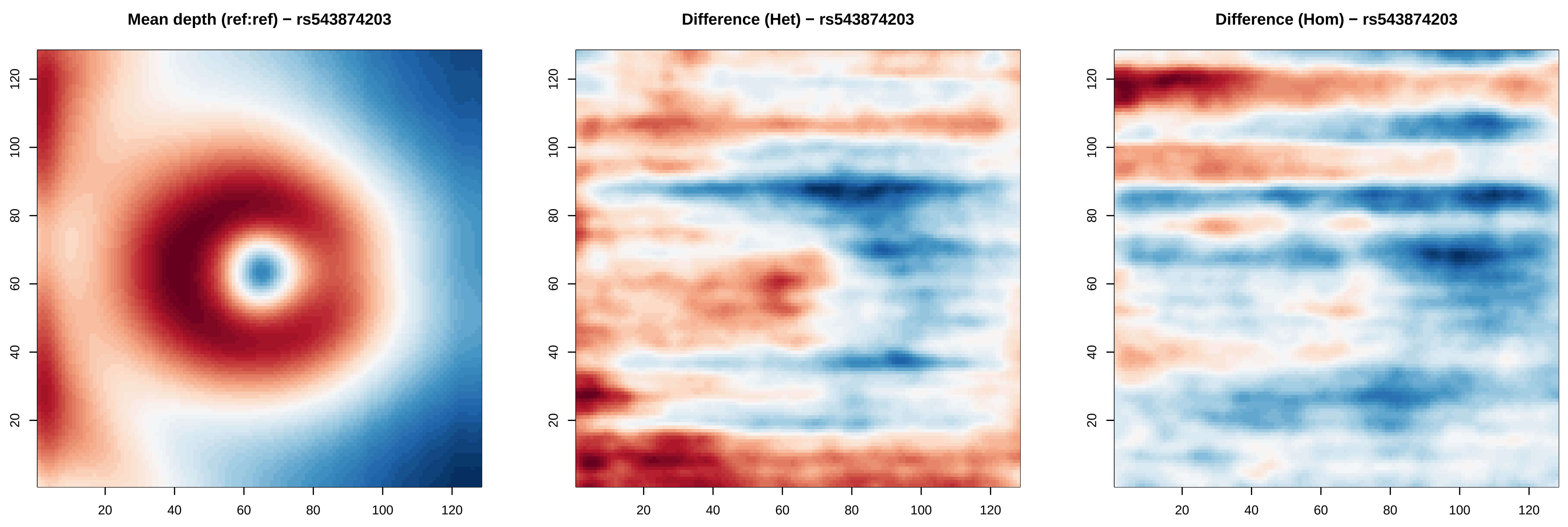
Mean depth (ref:ref) − rs543874203
Difference (Het) − rs543874203
Difference (Hom) − rs543874203
20
40
60
80
100
120

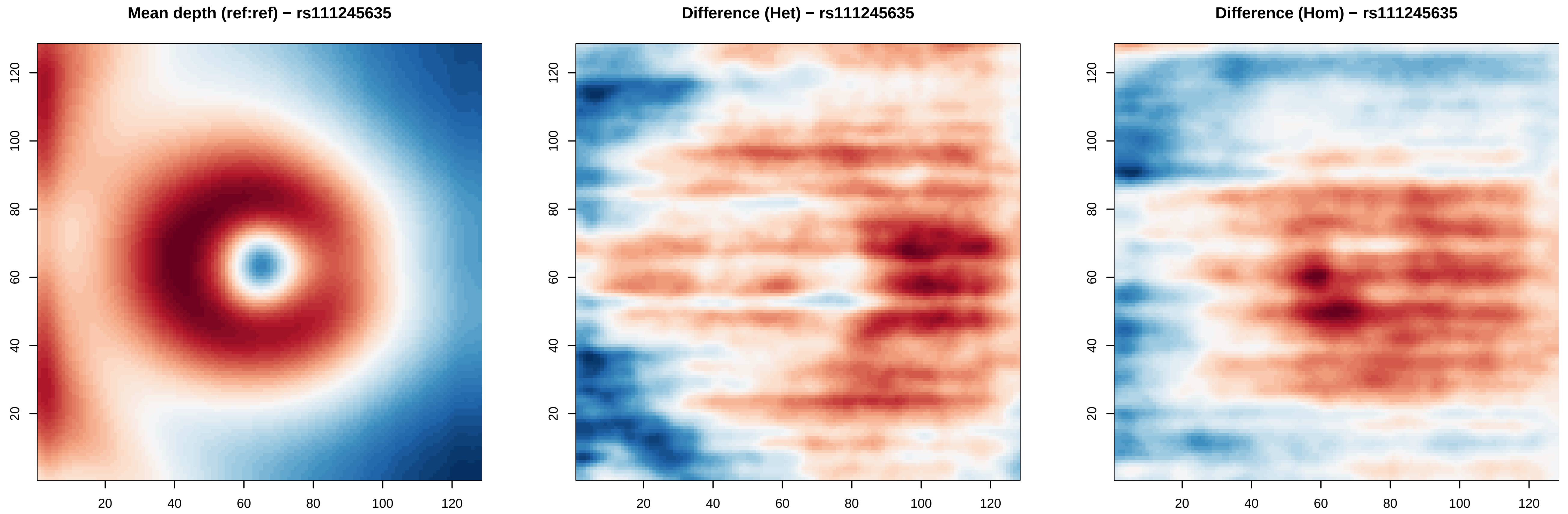
Mean depth (ref:ref) − rs111245635
Difference (Het) − rs111245635
Difference (Hom) − rs111245635
20
40
60
80
100
120

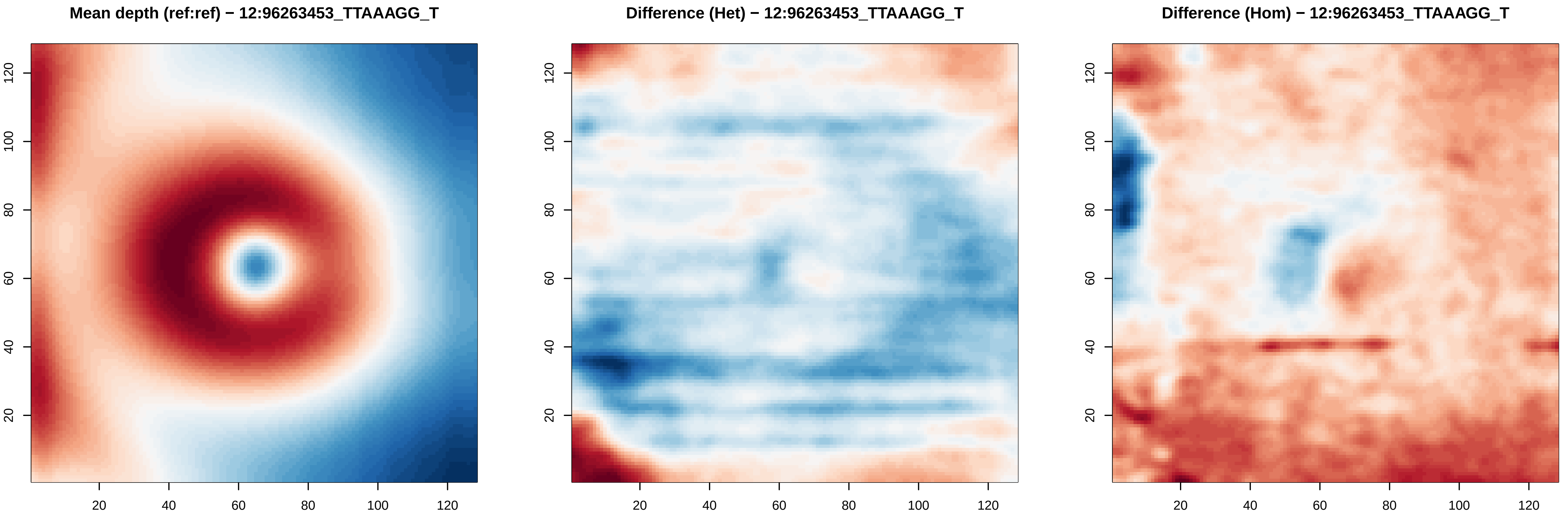
Mean depth (ref:ref) − 12:96263453_TTAAAGG_T
Difference (Het) − 12:96263453_TTAAAGG_T
Difference (Hom) − 12:96263453_TTAAAGG_T
20
40
60
80
100
120

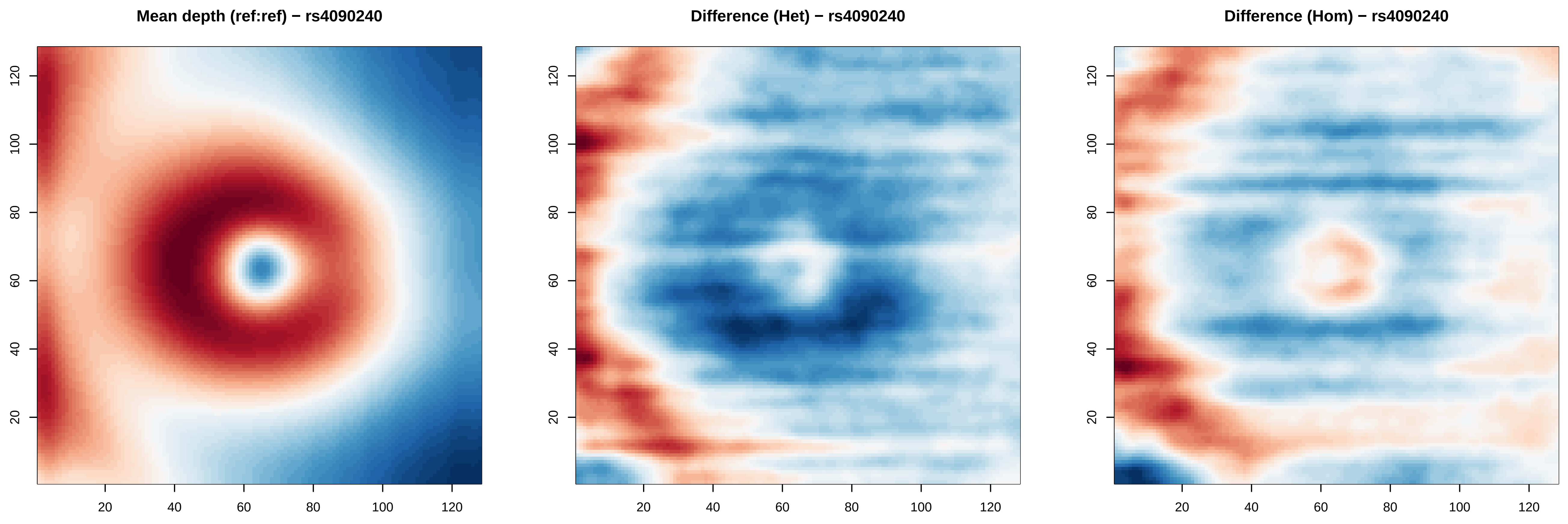

Mean depth (ref:ref) − rs4090240
Difference (Het) − rs4090240
Difference (Hom) − rs4090240
20
40
60
80
100
120

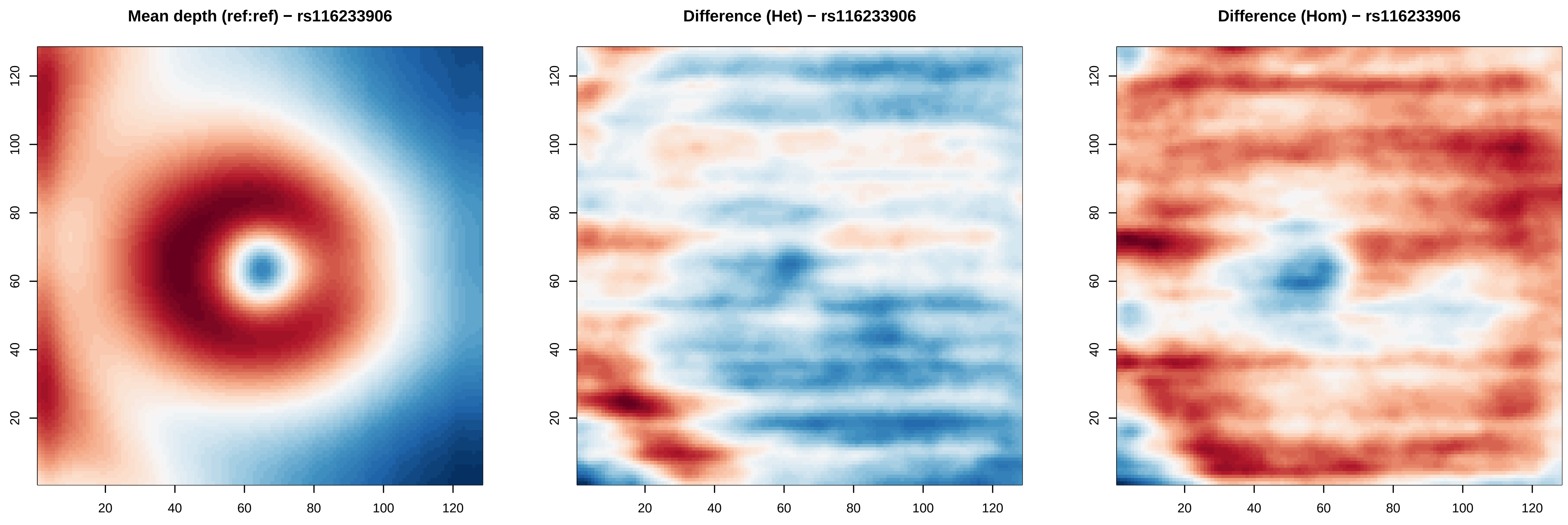
Mean depth (ref:ref) − rs116233906
Difference (Het) − rs116233906
Difference (Hom) − rs116233906
20
40
60
80
100
120

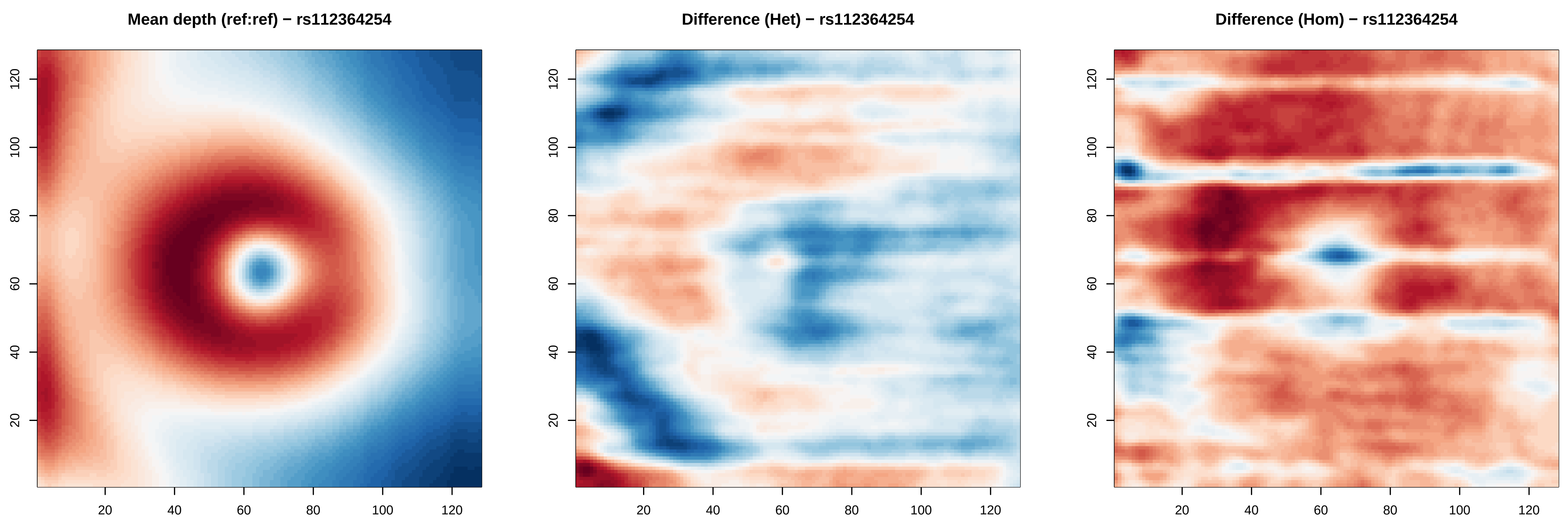

Mean depth (ref:ref) − rs112364254
Difference (Het) − rs112364254
Difference (Hom) − rs112364254
20
40
60
80
100
120

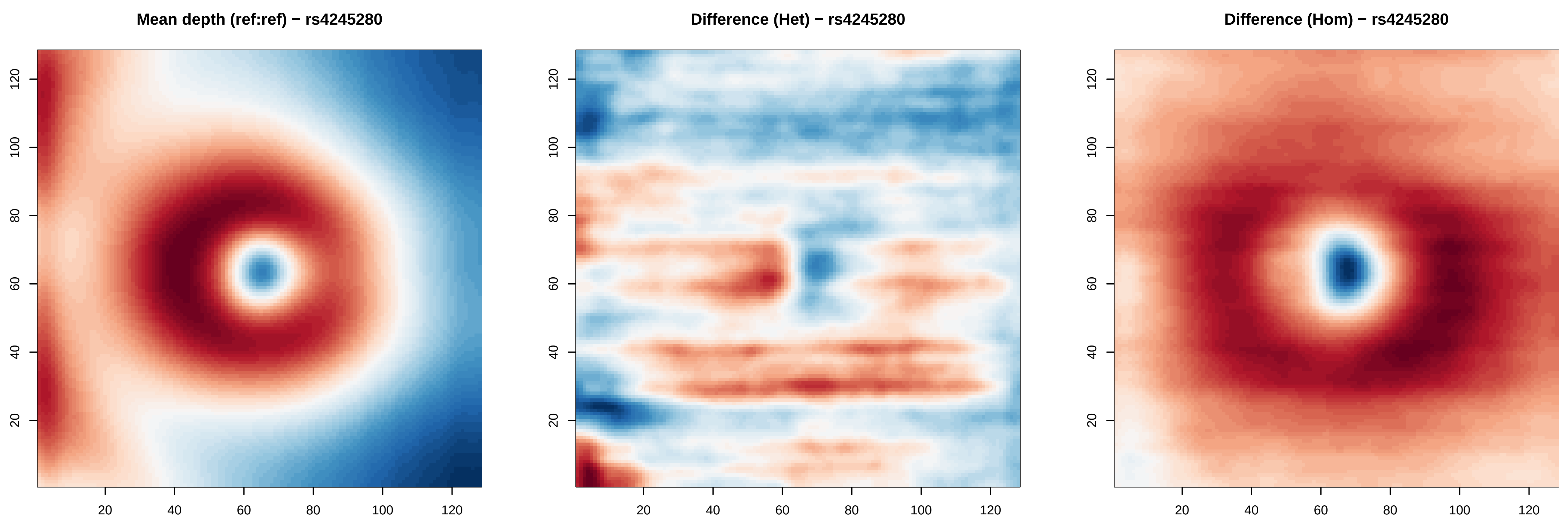
Mean depth (ref:ref) − rs4245280
Difference (Het) − rs4245280
Difference (Hom) − rs4245280
20
40
60
80
100
120

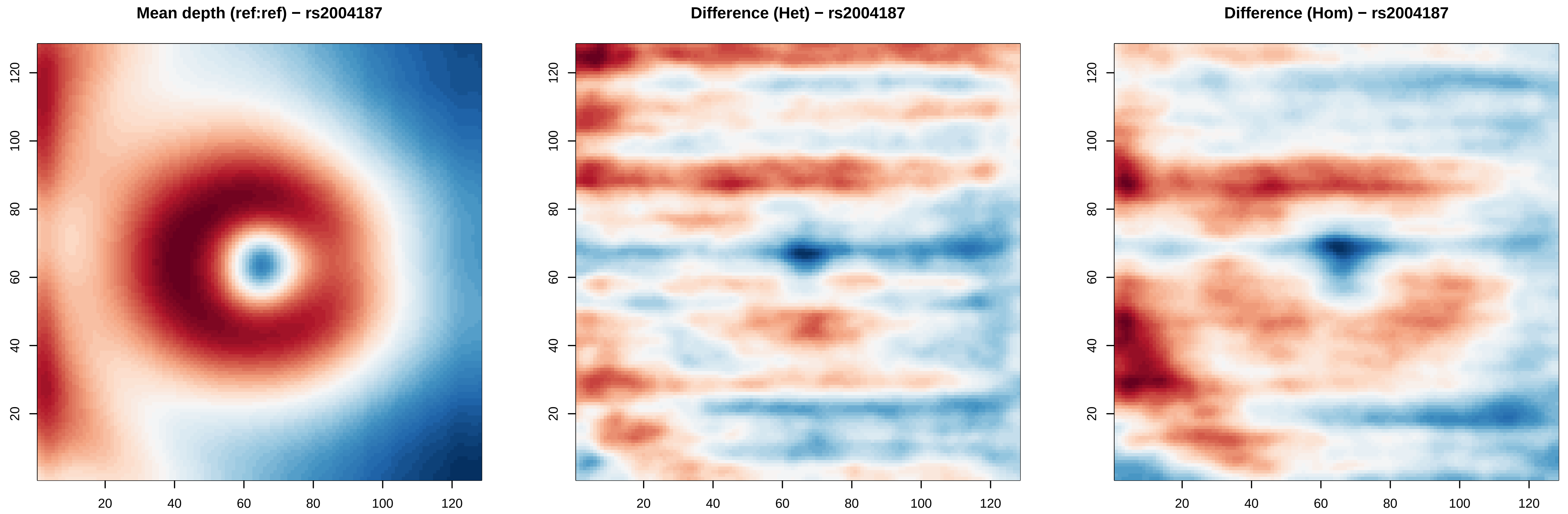

Mean depth (ref:ref) − rs2004187
Difference (Het) − rs2004187
Difference (Hom) − rs2004187
20
40
60
80
100
120

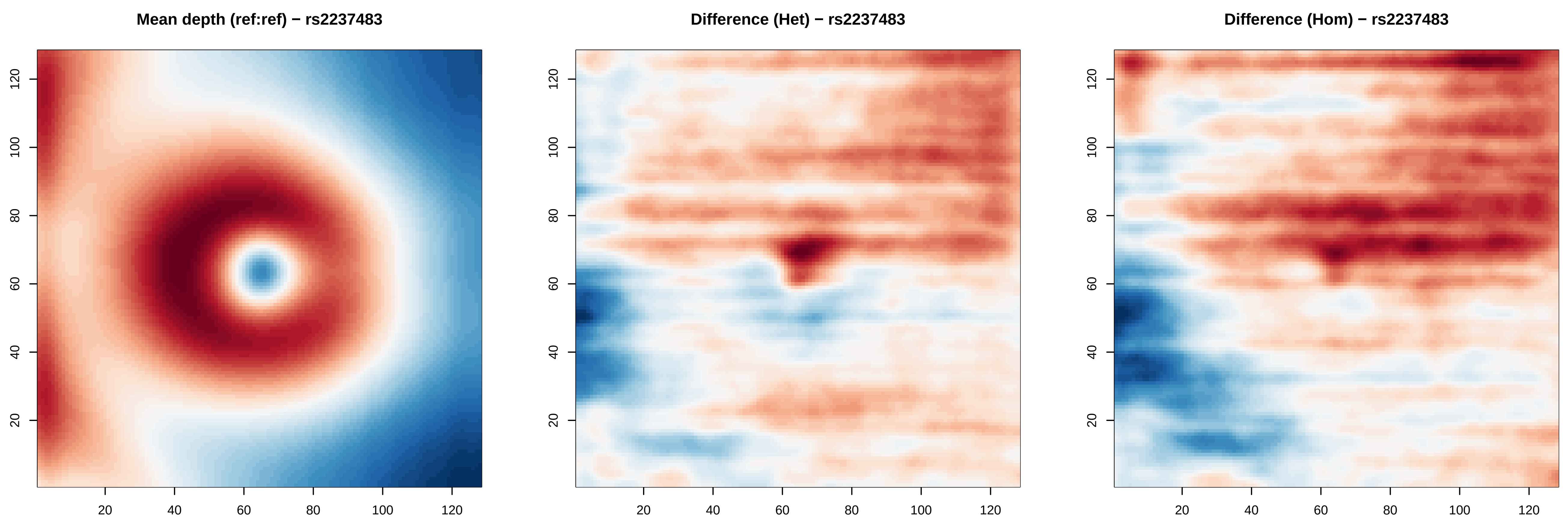

Mean depth (ref:ref) − rs2237483
Difference (Het) − rs2237483
Difference (Hom) − rs2237483
20
40
60
80
100
120

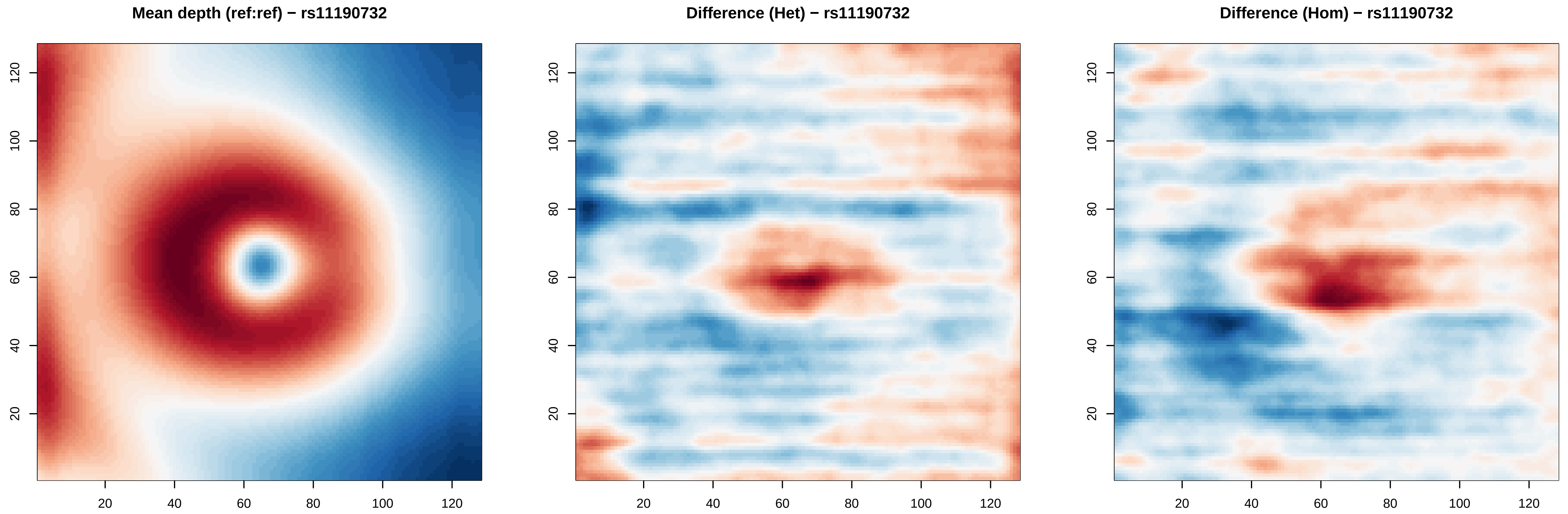
Mean depth (ref:ref) − rs11190732
Difference (Het) − rs11190732
Difference (Hom) − rs11190732
20
40
60
80
100
120

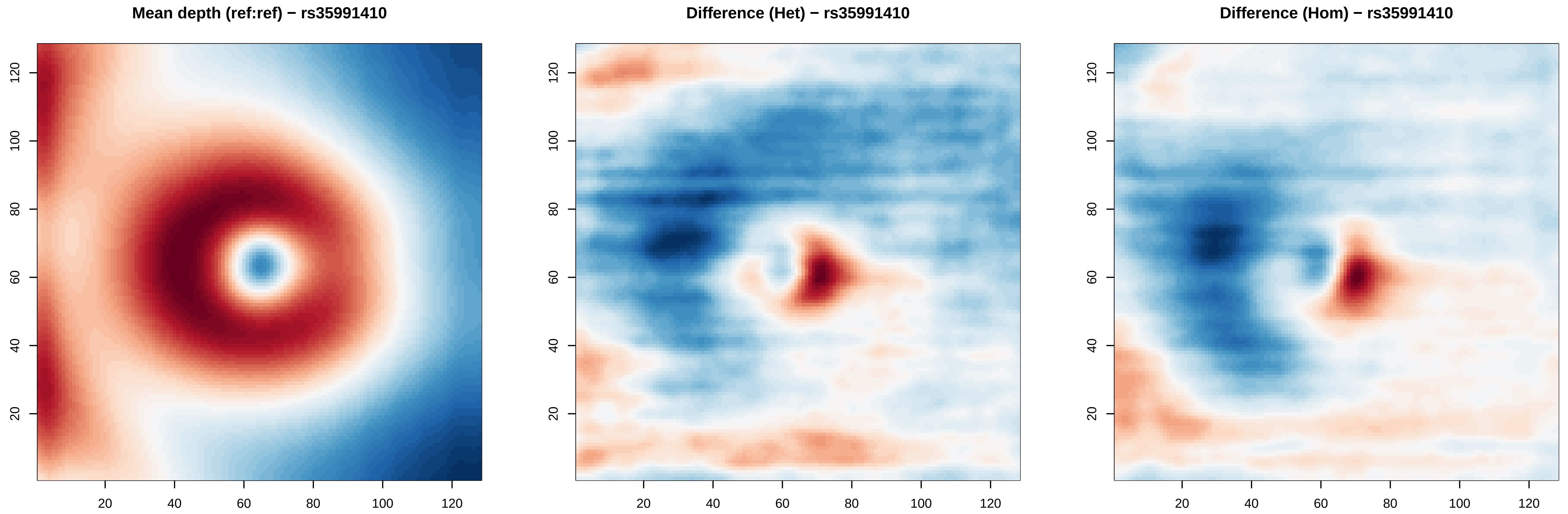
Mean depth (ref:ref) − rs35991410
Difference (Het) − rs35991410
Difference (Hom) − rs35991410
20
40
60
80
100
120

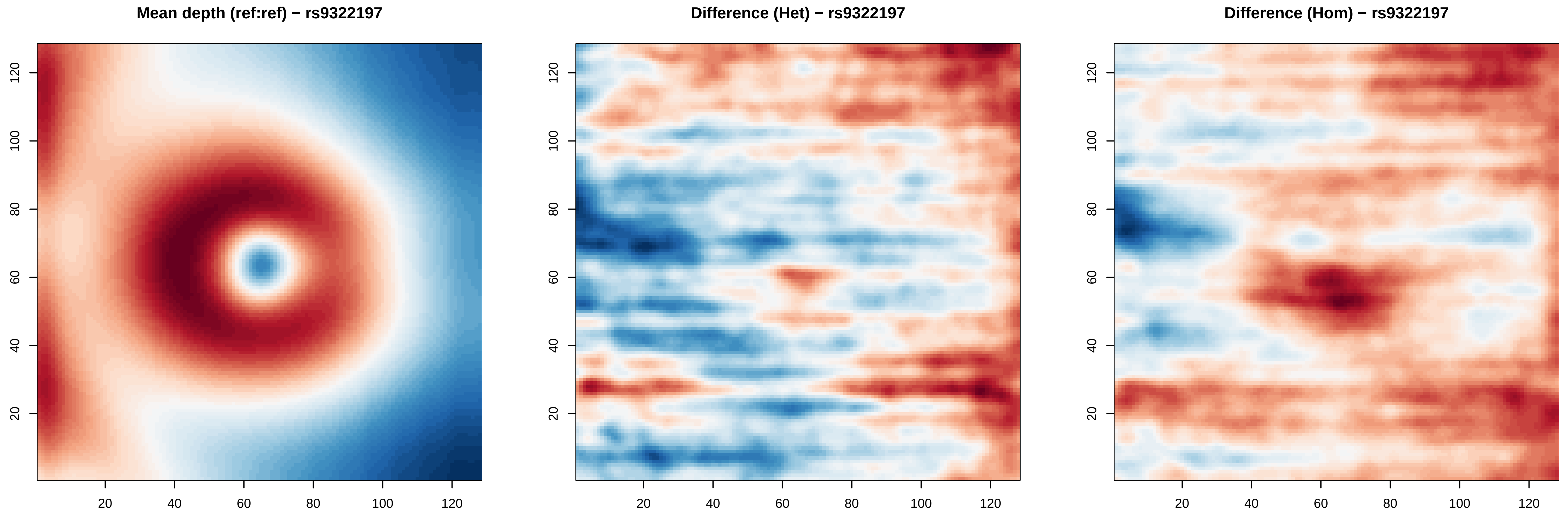

Mean depth (ref:ref) − rs9322197
Difference (Het) − rs9322197
Difference (Hom) − rs9322197
20
40
60
80
100
120

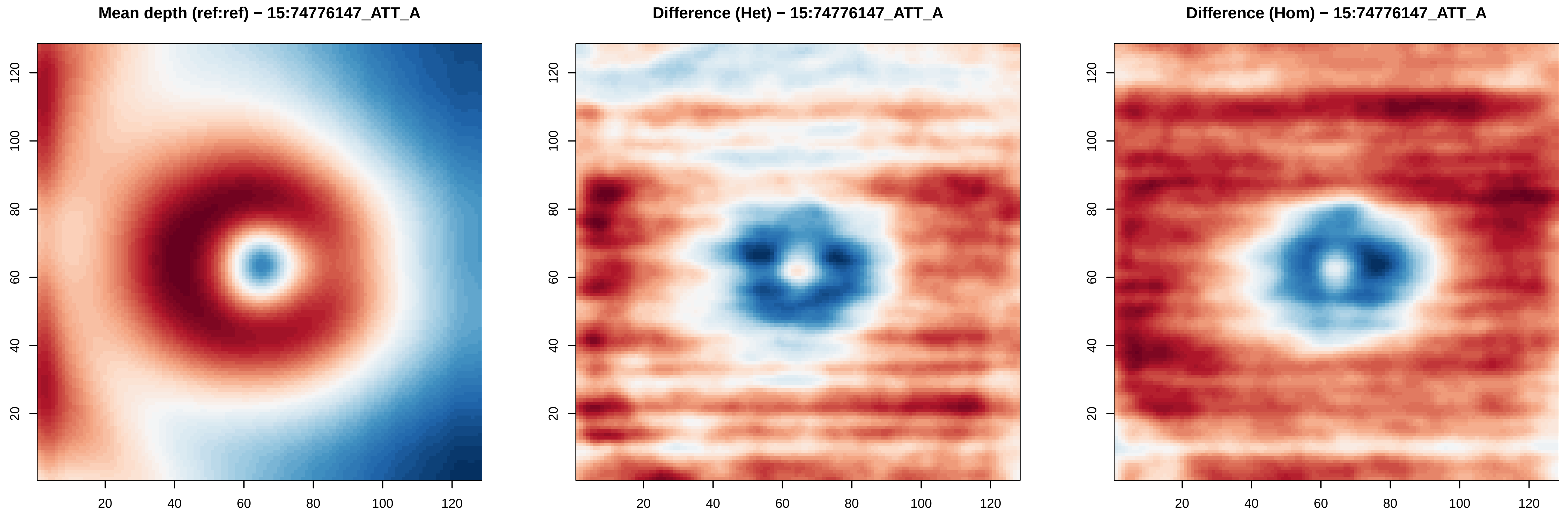
Mean depth (ref:ref) − 15:74776147_ATT_A
Difference (Het) − 15:74776147_ATT_A
Difference (Hom) − 15:74776147_ATT_A
20
40
60
80
100
120

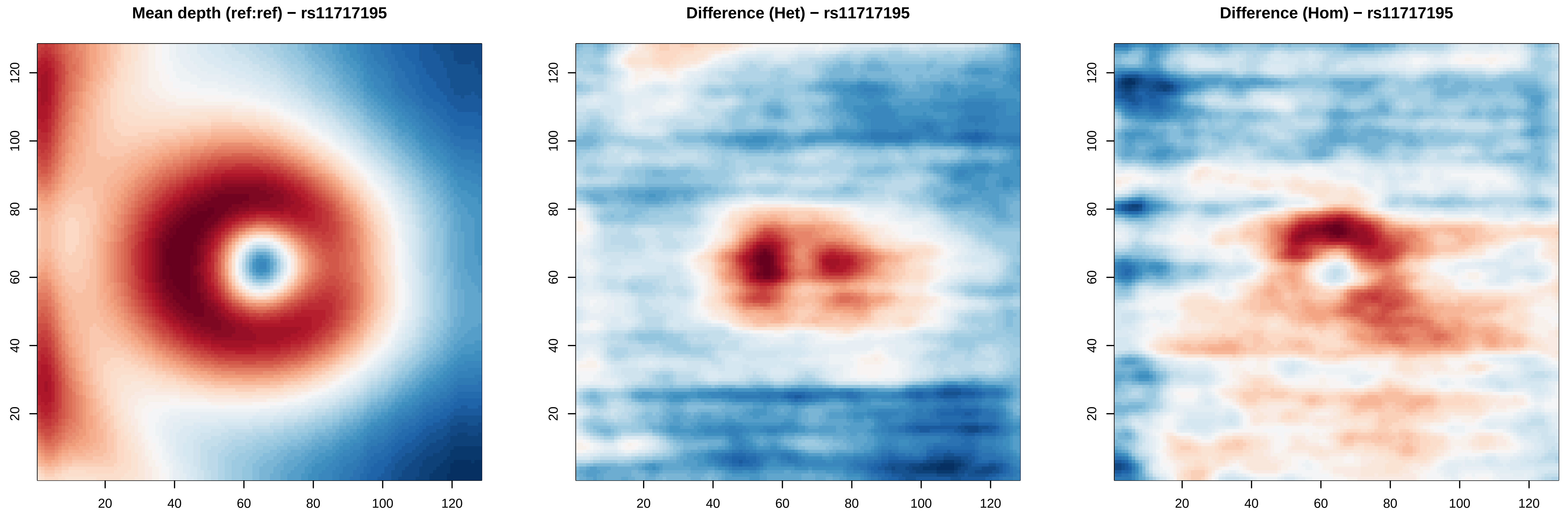
Mean depth (ref:ref) − rs11717195
Difference (Het) − rs11717195
Difference (Hom) − rs11717195
20
40
60
80
100
120

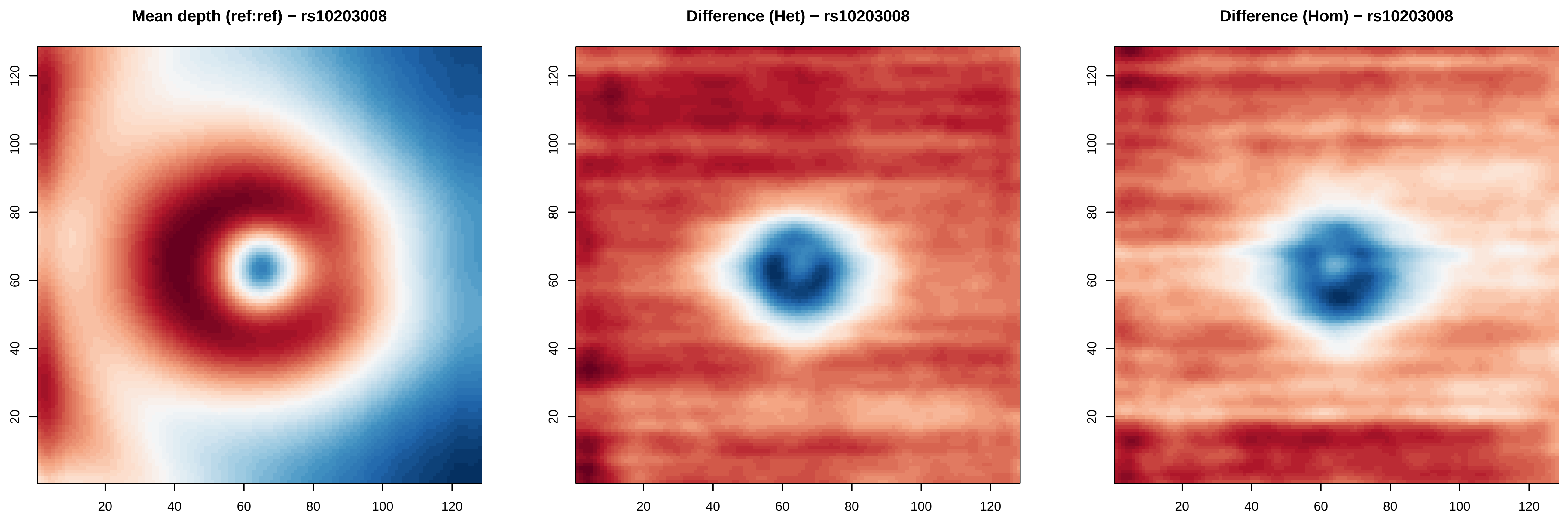
Mean depth (ref:ref) − rs10203008
Difference (Het) − rs10203008
Difference (Hom) − rs10203008
20
40
60
80
100
120

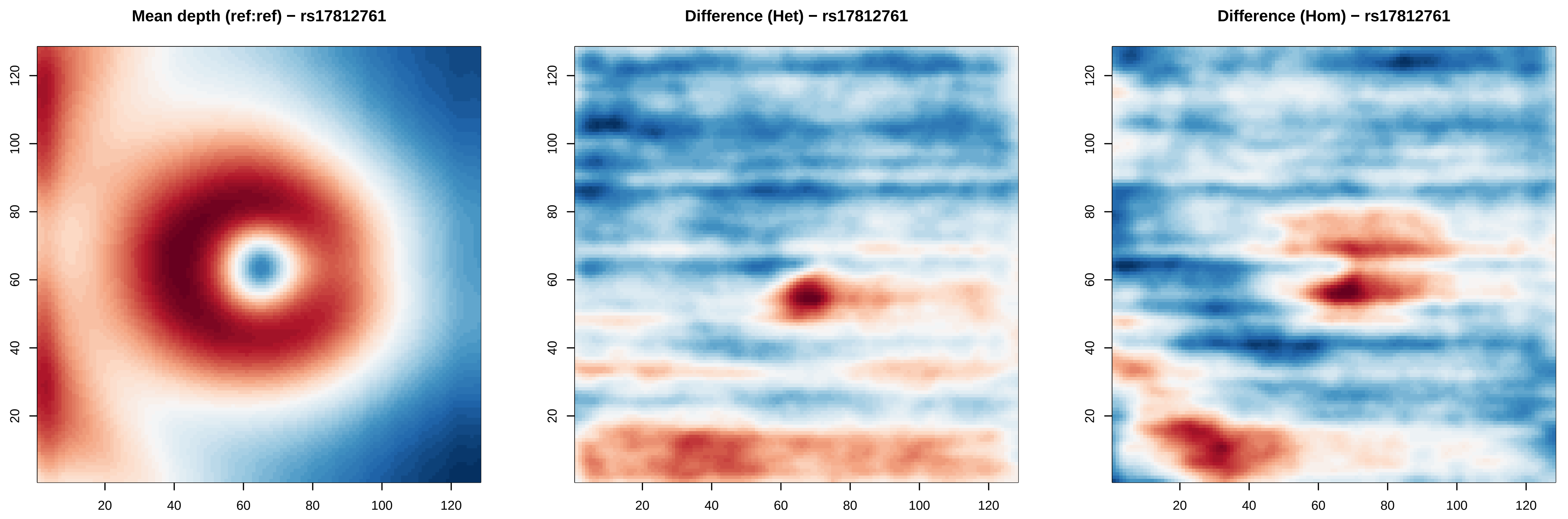

Mean depth (ref:ref) − rs17812761
Difference (Het) − rs17812761
Difference (Hom) − rs17812761
20
40
60
80
100
120

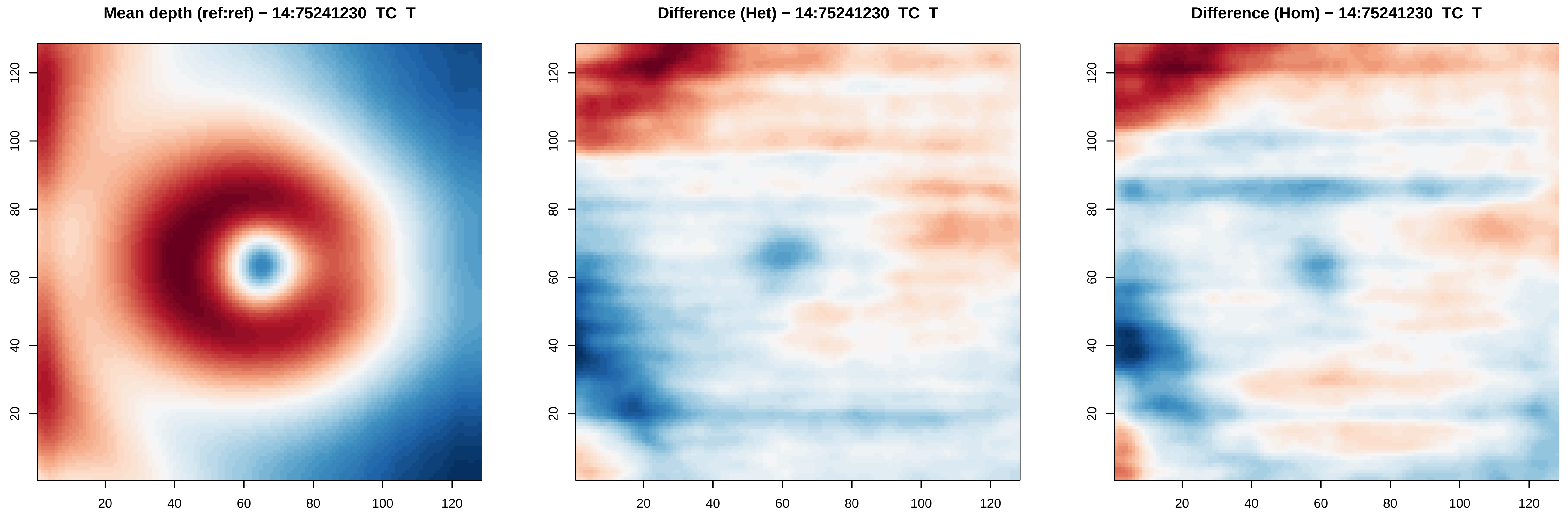
Mean depth (ref:ref) − 14:75241230_TC_T
Difference (Het) − 14:75241230_TC_T
Difference (Hom) − 14:75241230_TC_T
20
40
60
80
100
120

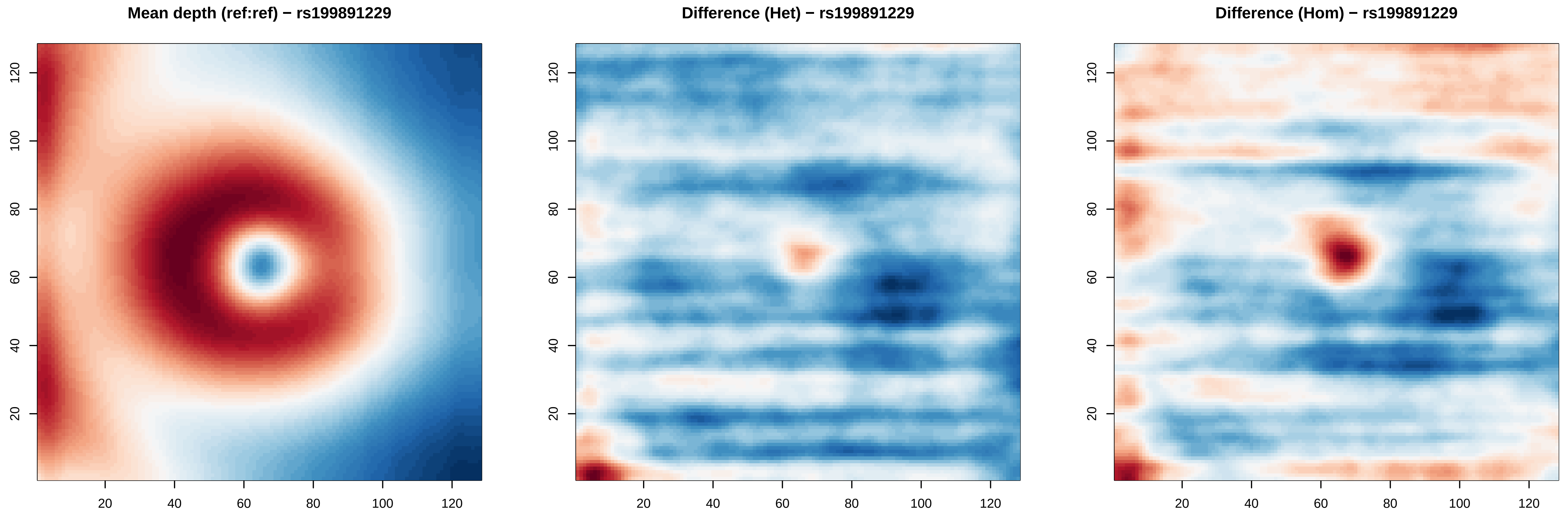
Mean depth (ref:ref) − rs199891229
Difference (Het) − rs199891229
Difference (Hom) − rs199891229
20
40
60
80
100
120

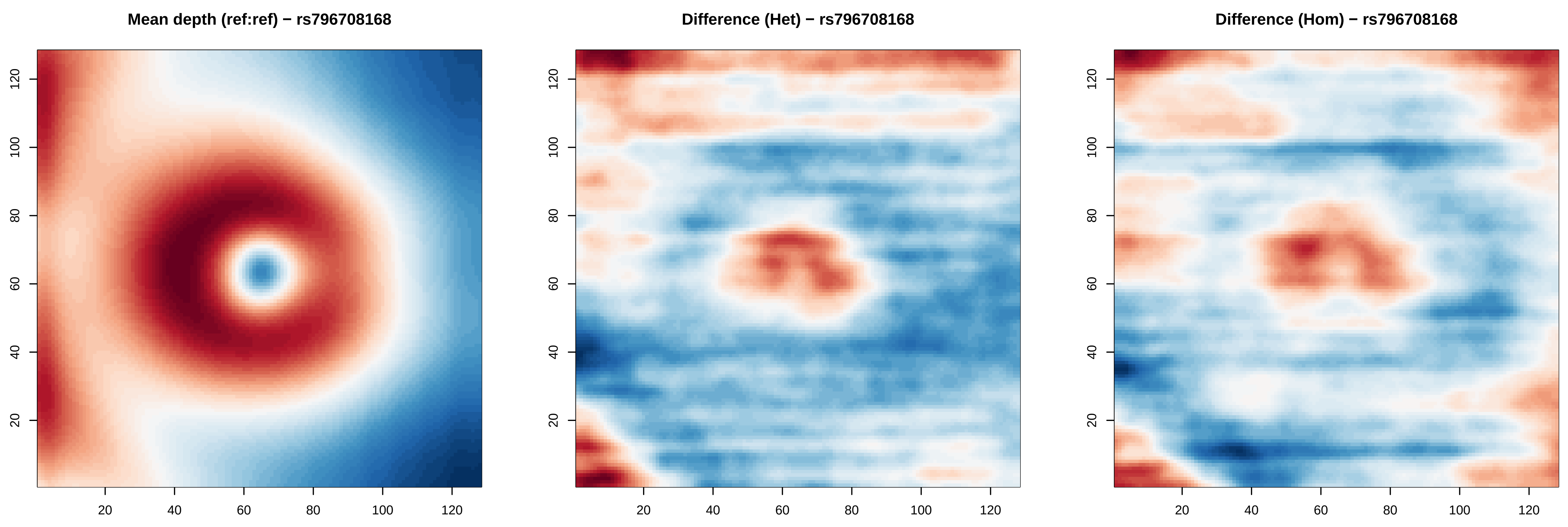
Mean depth (ref:ref) − rs796708168
Difference (Het) − rs796708168
Difference (Hom) − rs796708168
20
40
60
80
100
120

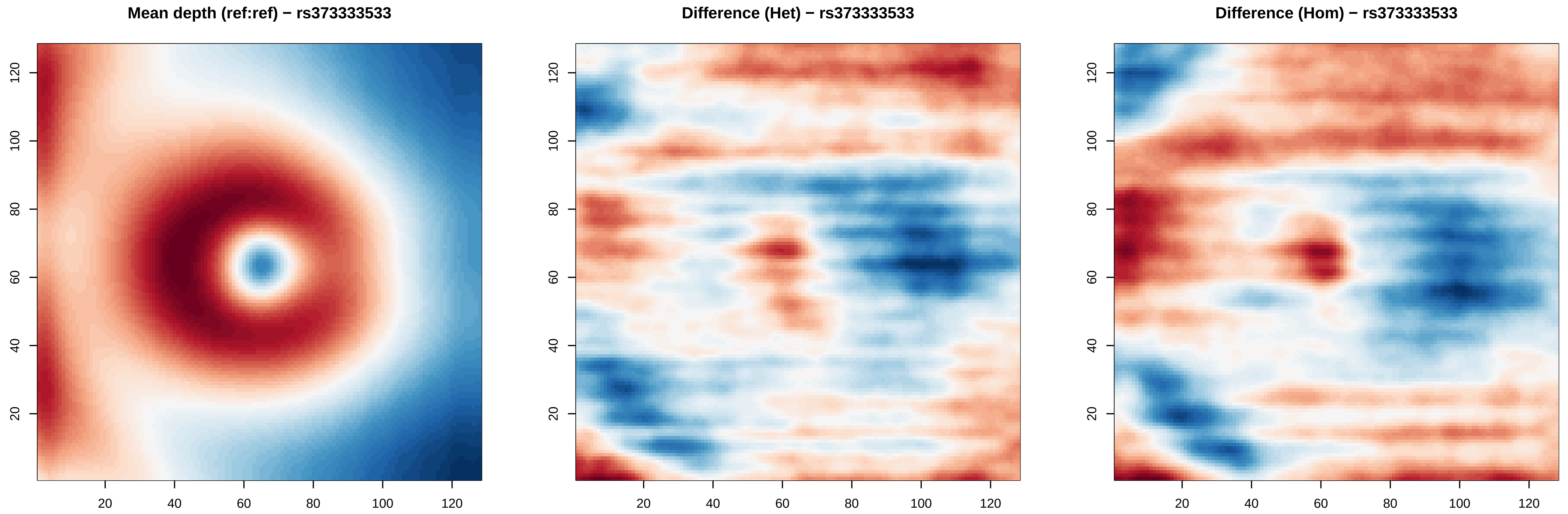
Mean depth (ref:ref) − rs373333533
Difference (Het) − rs373333533
Difference (Hom) − rs373333533
20
40
60
80
100
120

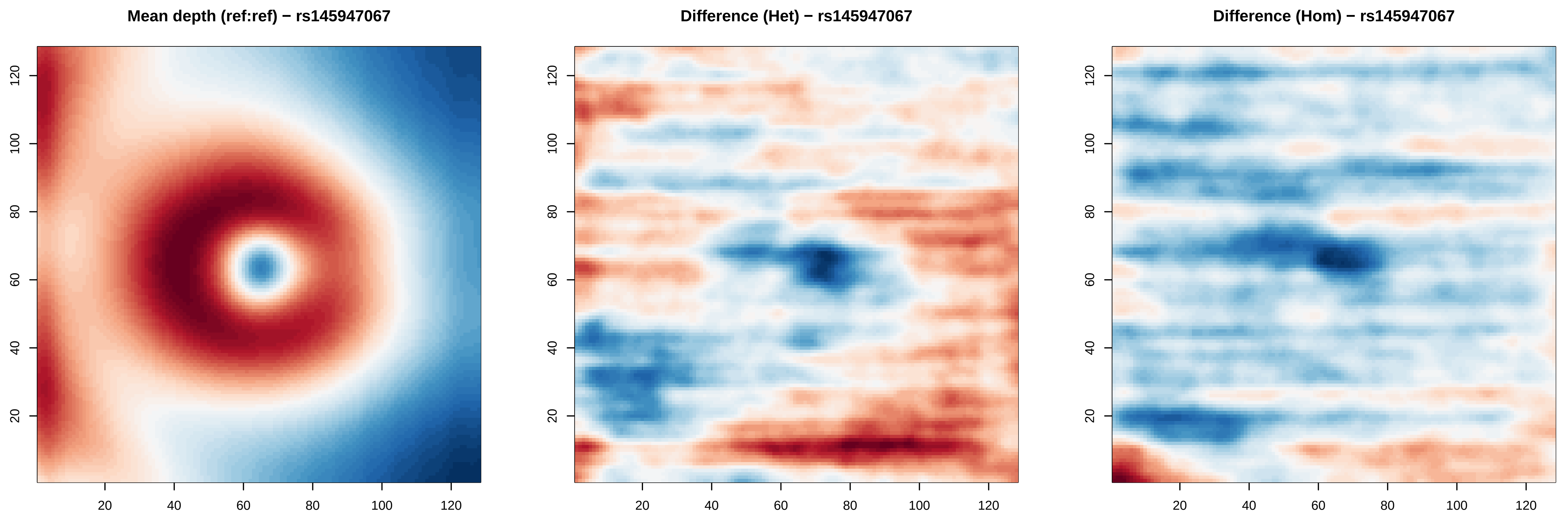

Mean depth (ref:ref) − rs145947067
Difference (Het) − rs145947067
Difference (Hom) − rs145947067
20
40
60
80
100
120

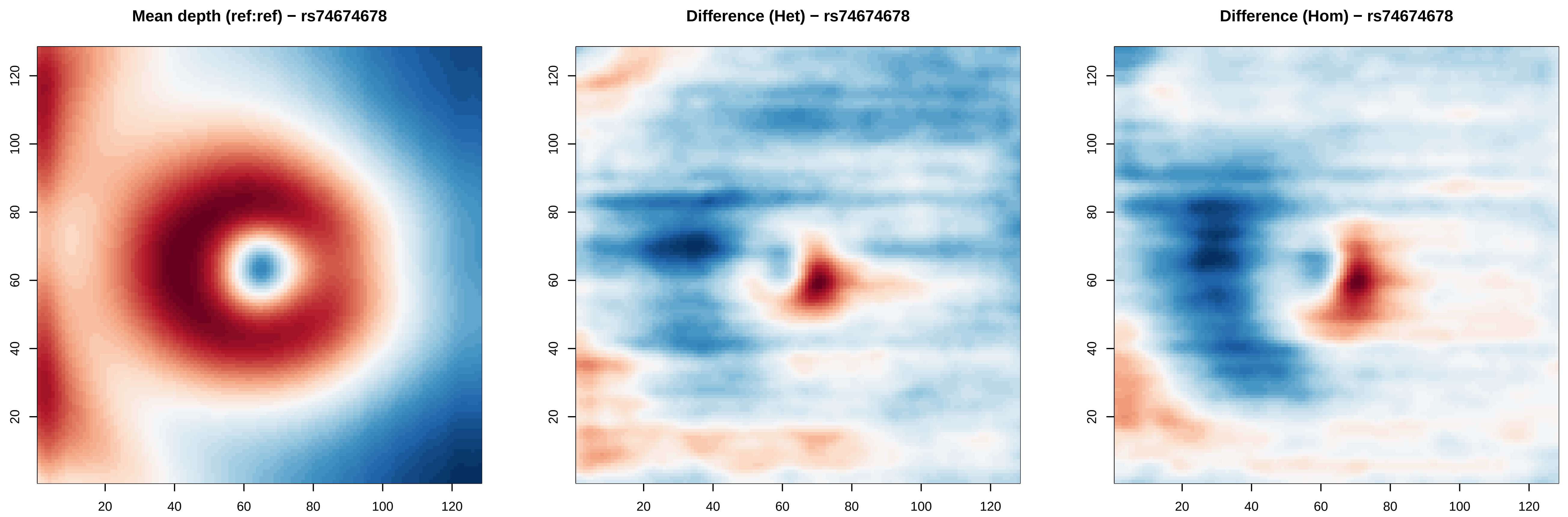

Mean depth (ref:ref) − rs74674678
Difference (Het) − rs74674678
Difference (Hom) − rs74674678
20
40
60
80
100
120

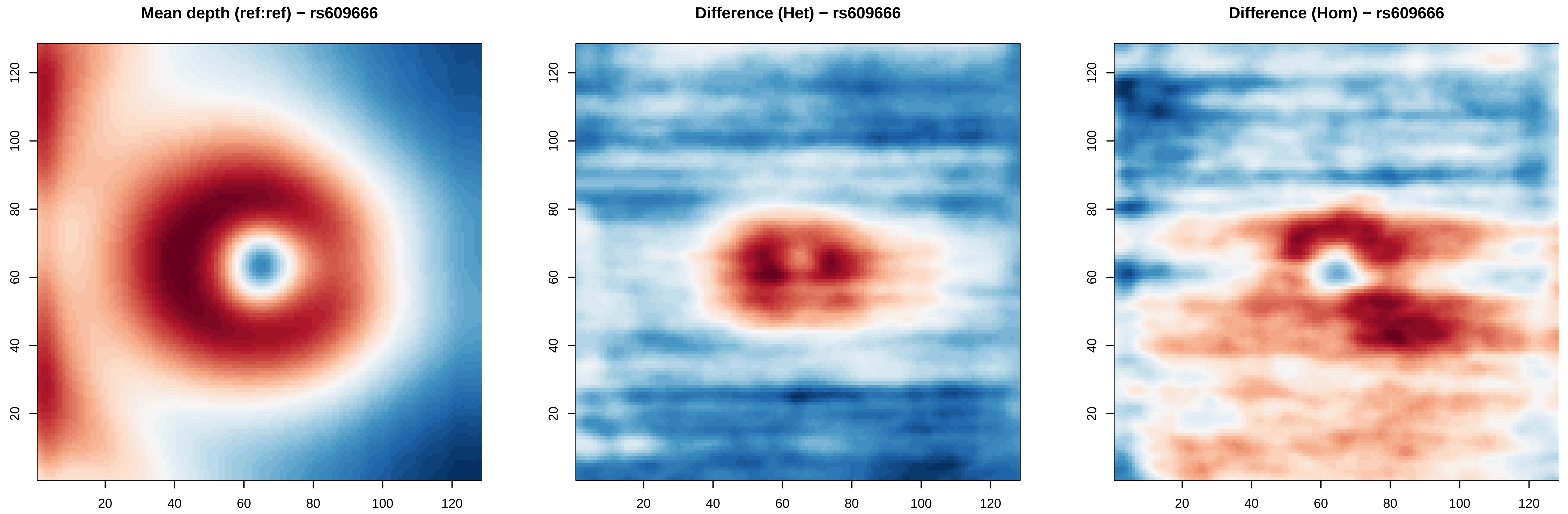

Mean depth (ref:ref) − rs609666
Difference (Het) − rs609666
Difference (Hom) − rs609666
20
40
60
80
100
120

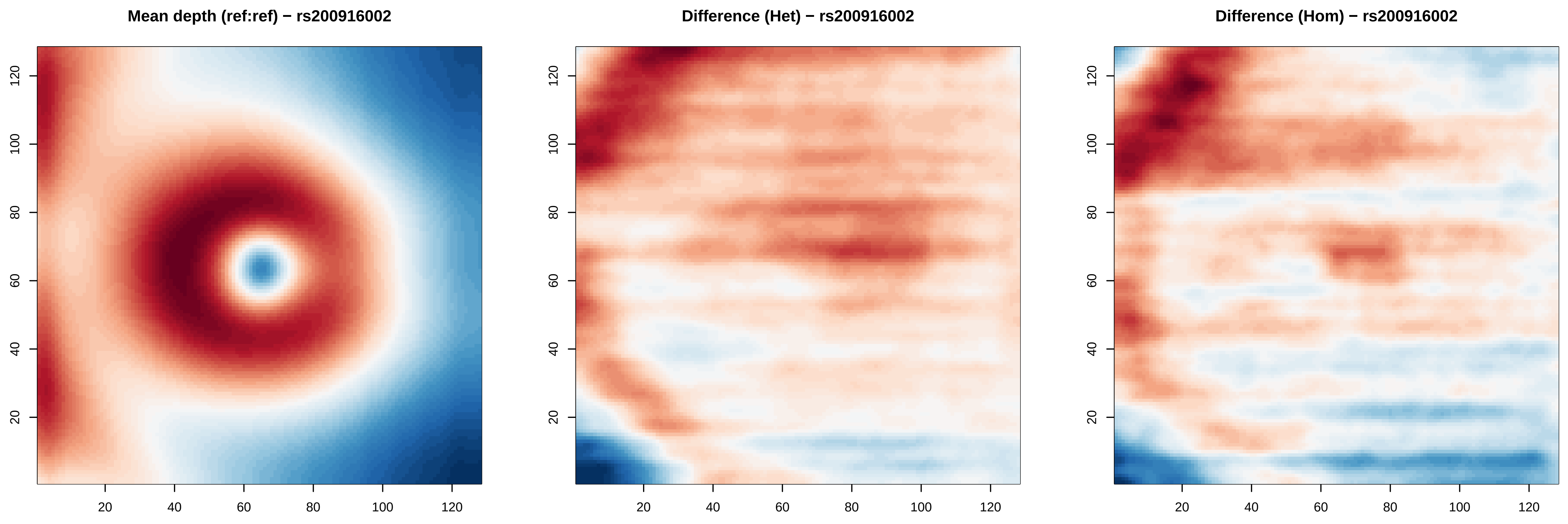

Mean depth (ref:ref) − rs200916002
Difference (Het) − rs200916002
Difference (Hom) − rs200916002
20
40
60
80
100
120

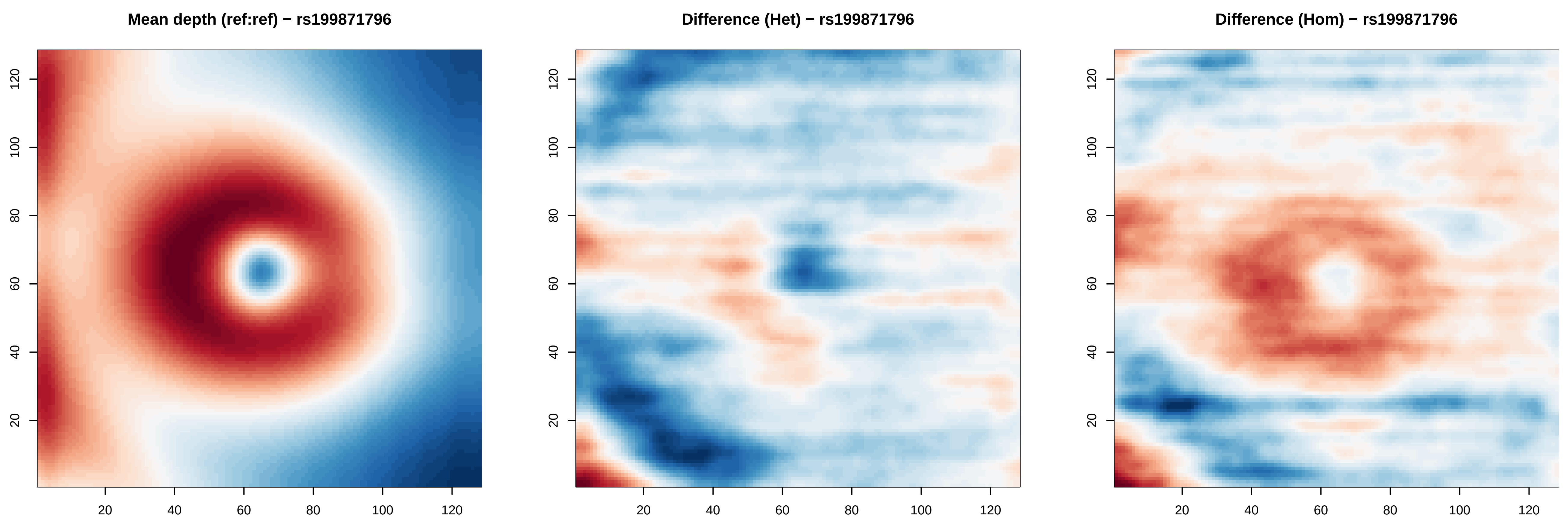

Mean depth (ref:ref) − rs199871796
Difference (Het) − rs199871796
Difference (Hom) − rs199871796
20
40
60
80
100
120

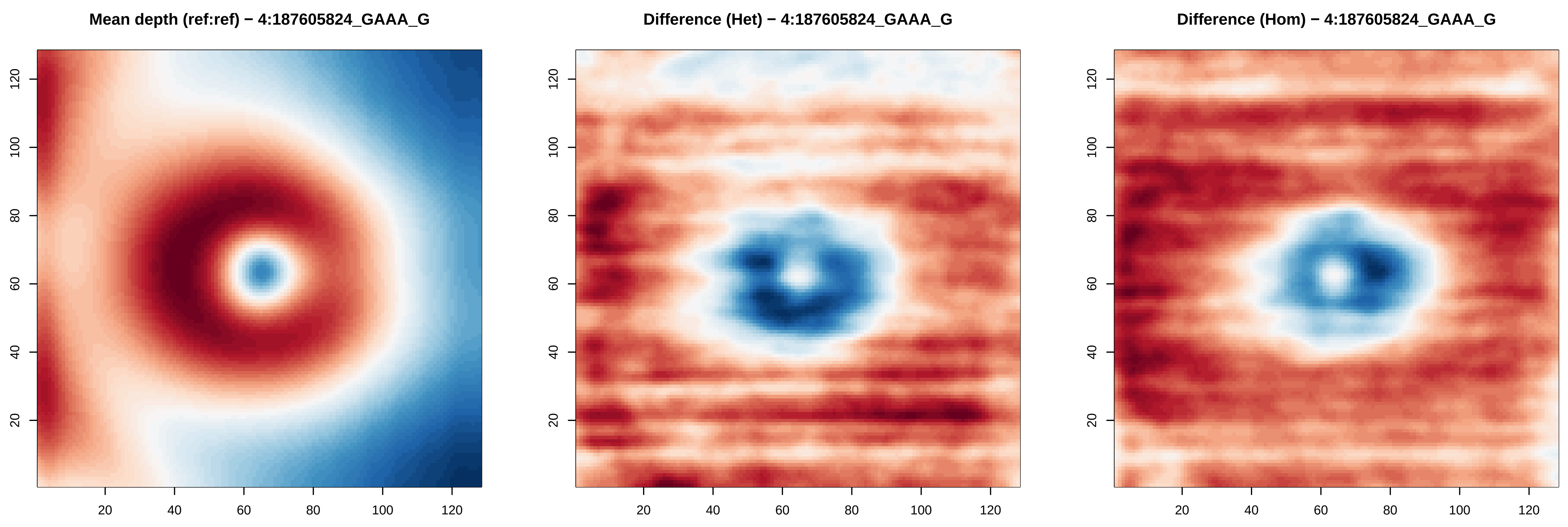

Mean depth (ref:ref) − 4:187605824_GAAA_G
Difference (Het) − 4:187605824_GAAA_G
Difference (Hom) − 4:187605824_GAAA_G
20
40
60
80
100
120

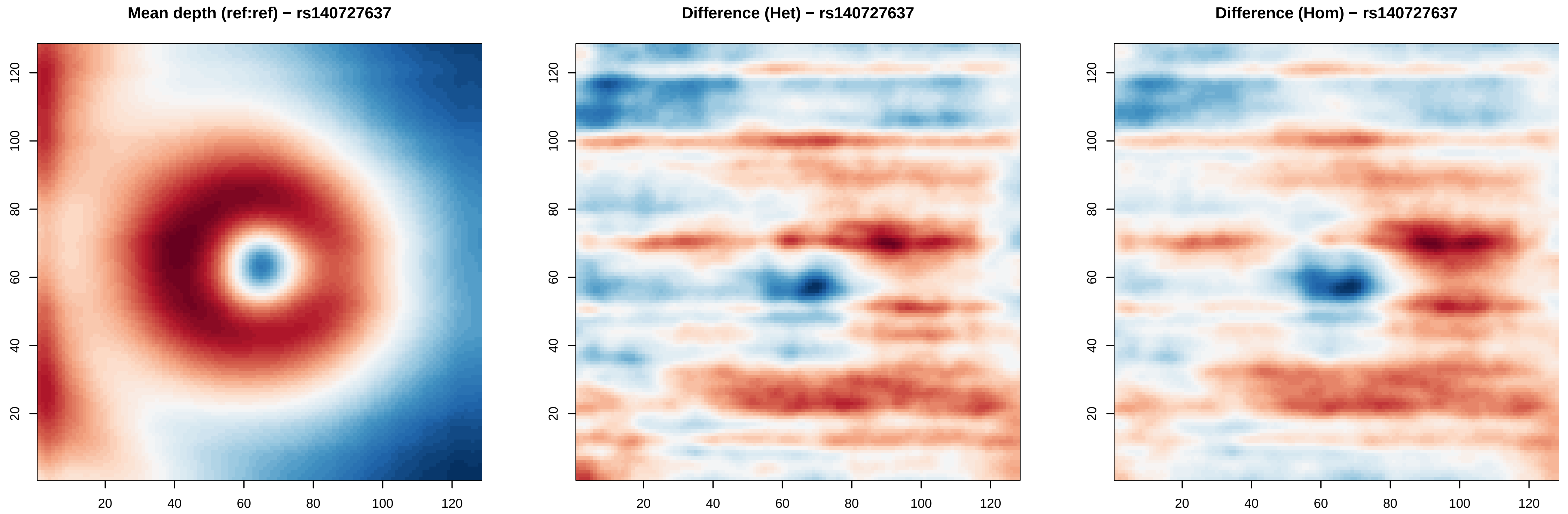
Mean depth (ref:ref) − rs140727637
Difference (Het) − rs140727637
Difference (Hom) − rs140727637
20
40
60
80
100
120
20
40
60
80
100
120

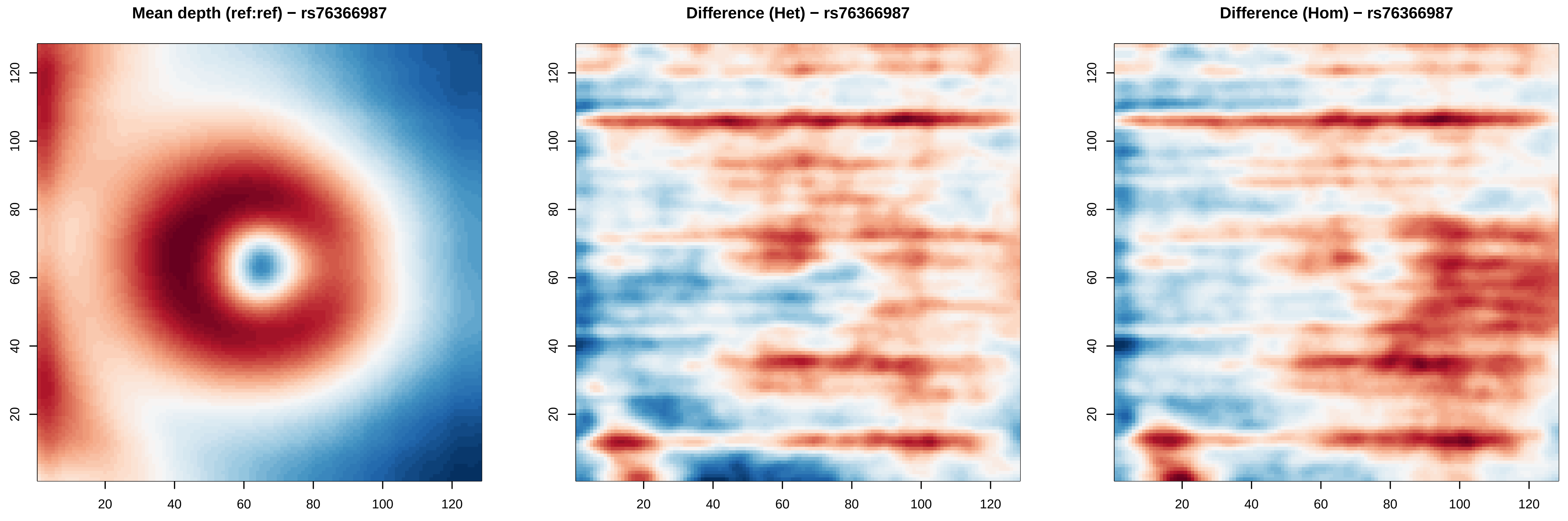

Mean depth (ref:ref) − rs76366987
Difference (Het) − rs76366987
Difference (Hom) − rs76366987
20
40
60
80
100
120

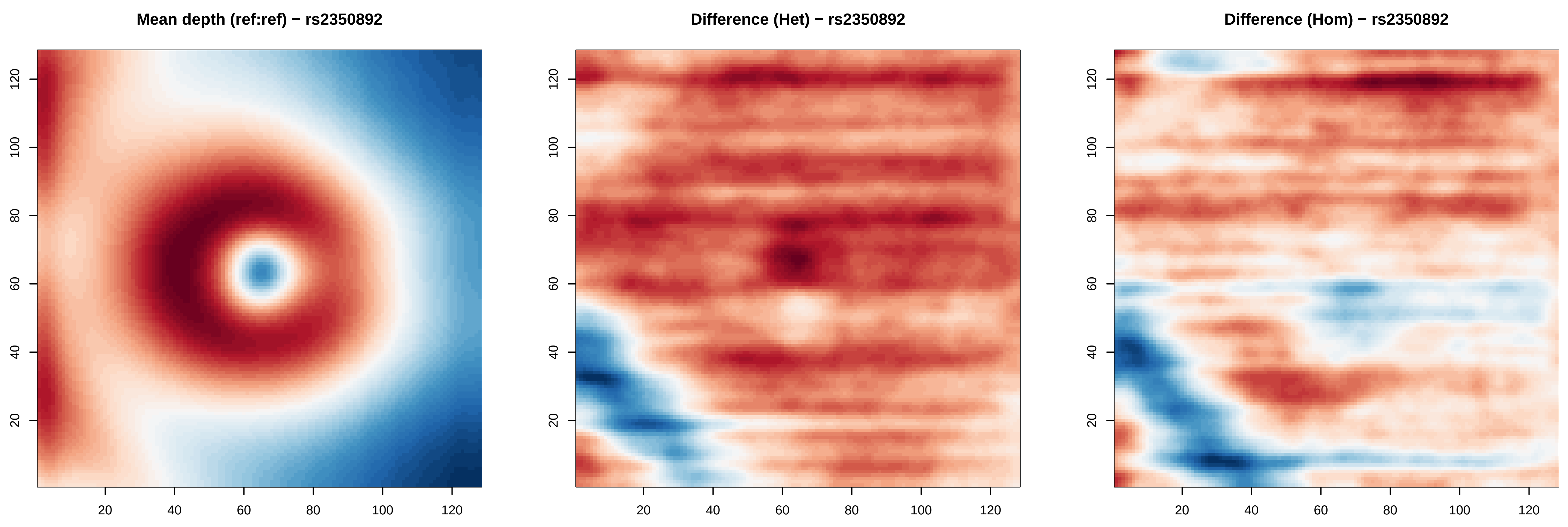
Mean depth (ref:ref) − rs2350892
Difference (Het) − rs2350892
Difference (Hom) − rs2350892
20
40
60
80
100
120

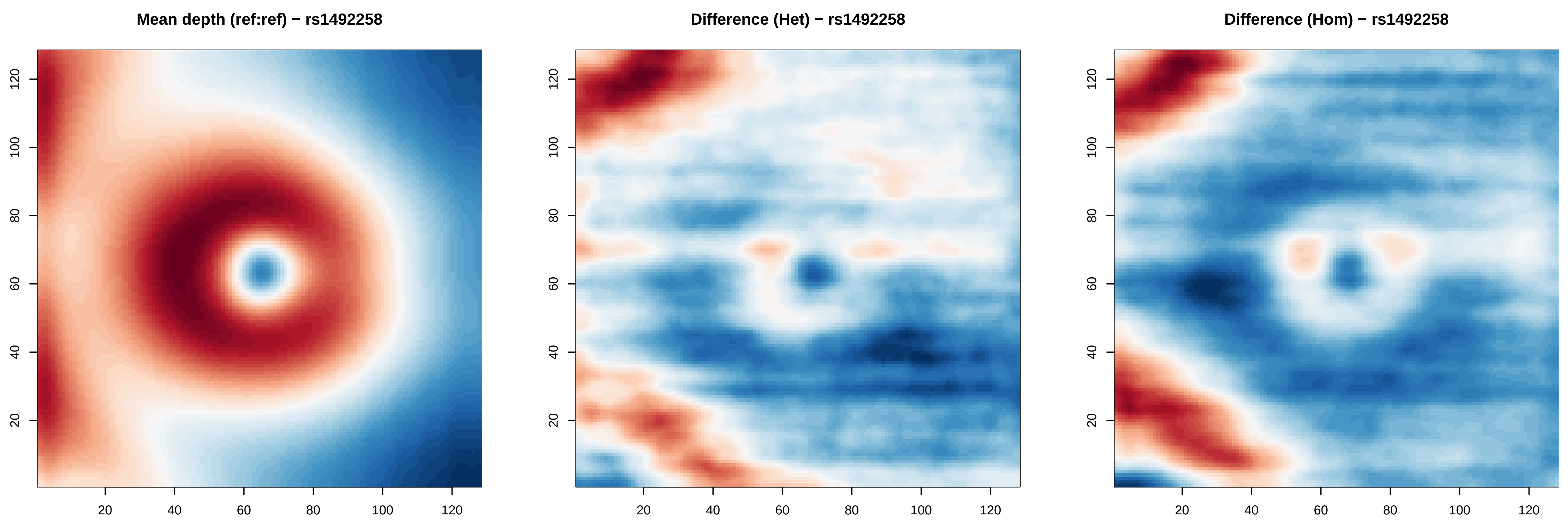

Mean depth (ref:ref) − rs1492258
Difference (Het) − rs1492258
Difference (Hom) − rs1492258
20
40
60
80
100
120

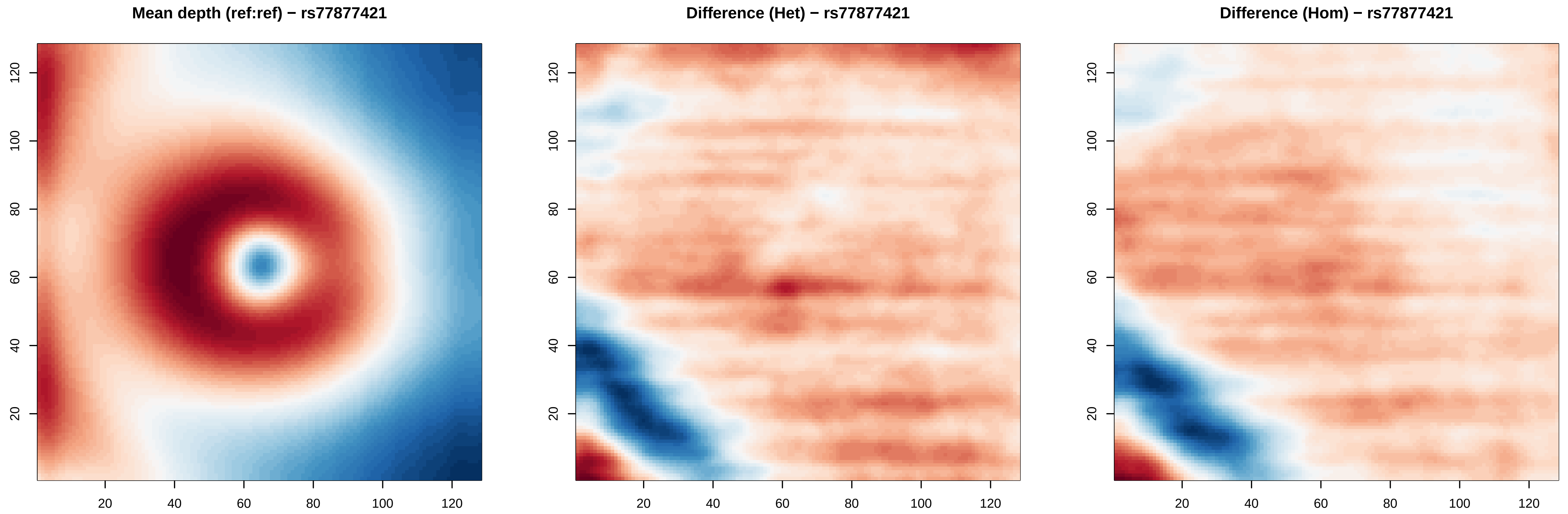
Mean depth (ref:ref) − rs77877421
Difference (Het) − rs77877421
Difference (Hom) − rs77877421
20
40
60
80
100
120

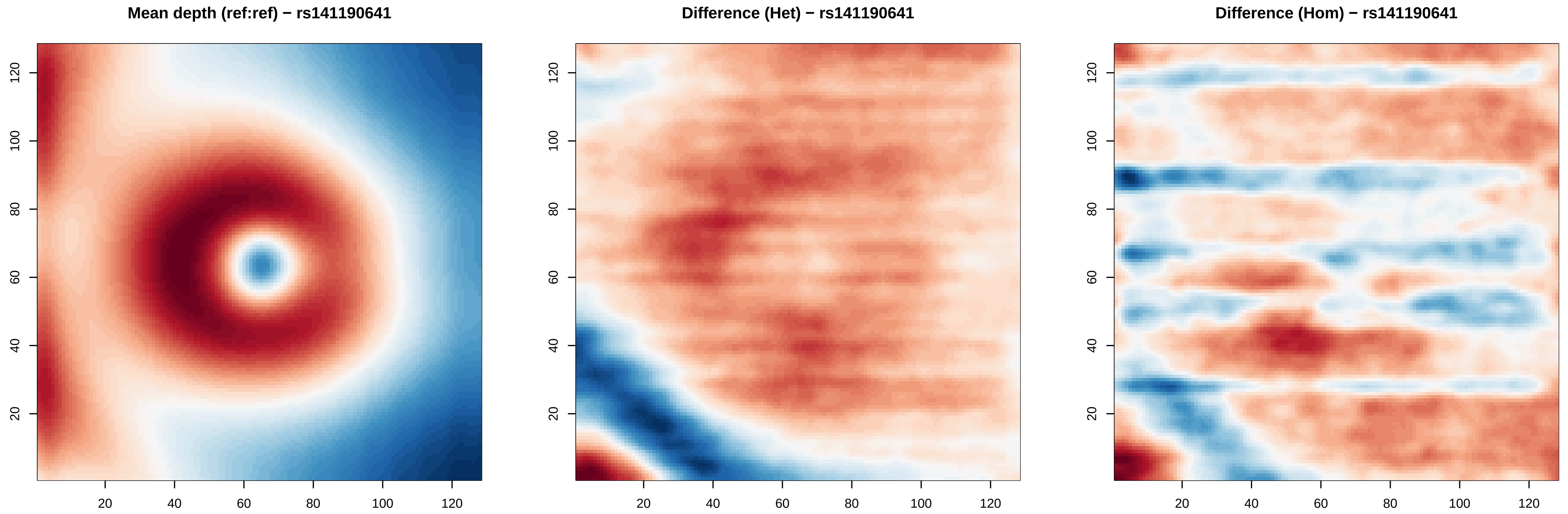
Mean depth (ref:ref) − rs141190641
Difference (Het) − rs141190641
Difference (Hom) − rs141190641
20
40
60
80
100
120

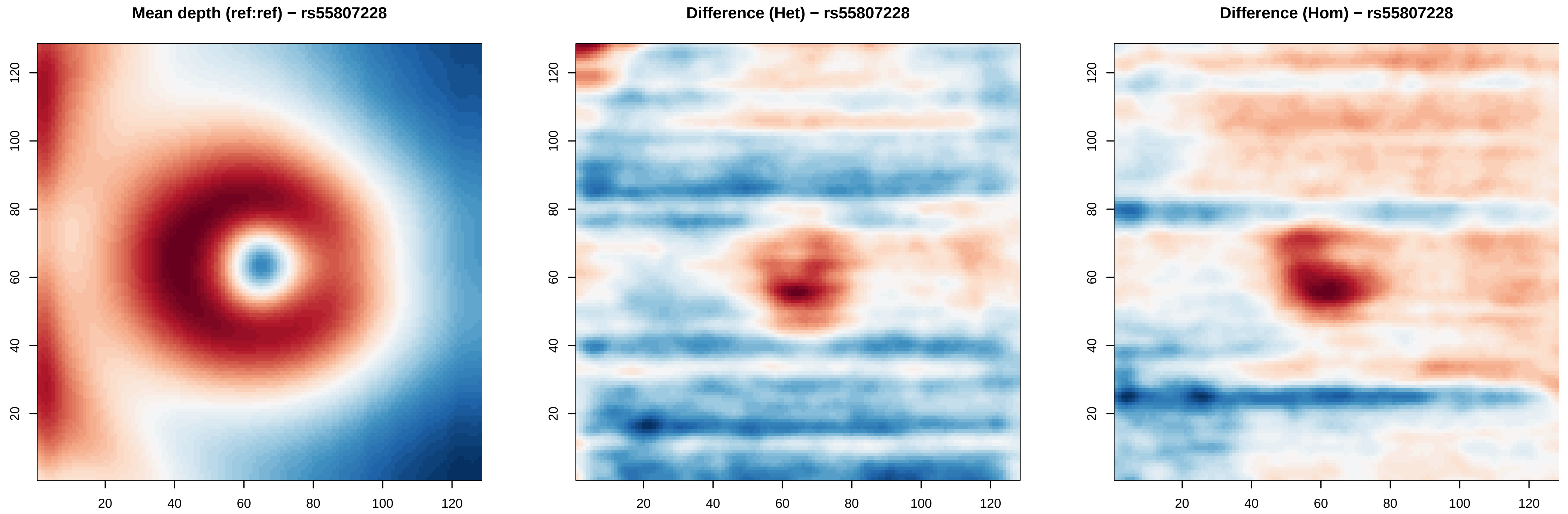
Mean depth (ref:ref) − rs55807228
Difference (Het) − rs55807228
Difference (Hom) − rs55807228
20
40
60
80
100
120

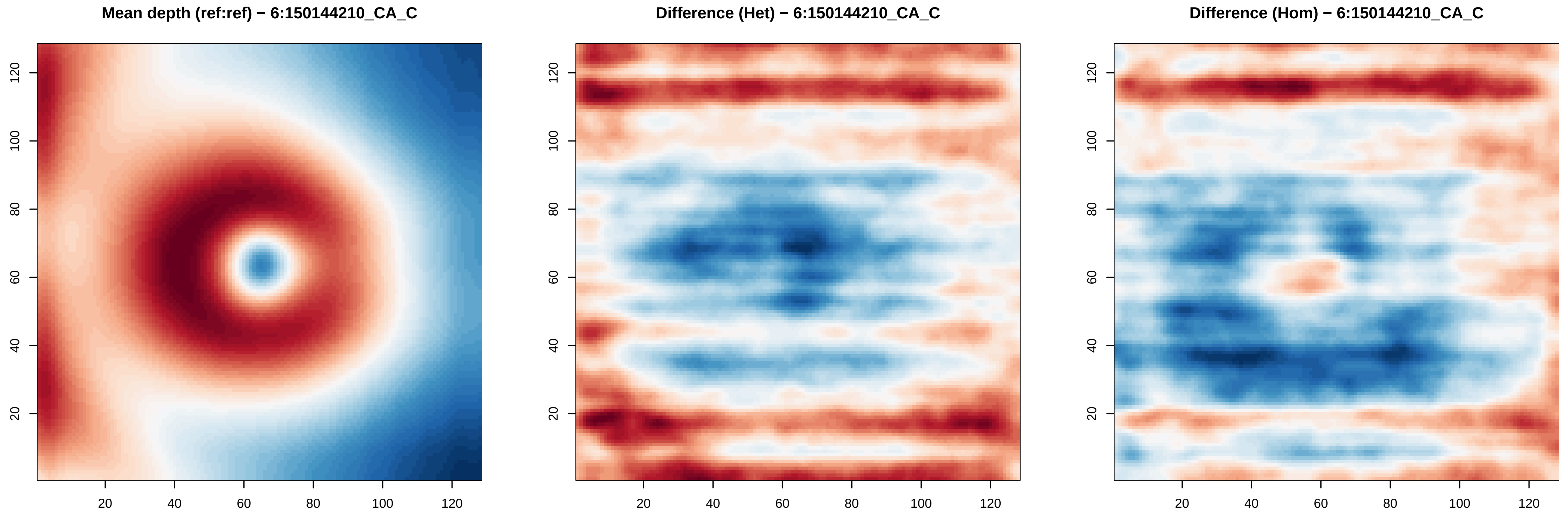

Mean depth (ref:ref) − 6:150144210_CA_C
Difference (Het) − 6:150144210_CA_C
Difference (Hom) − 6:150144210_CA_C
20
40
60
80
100
120

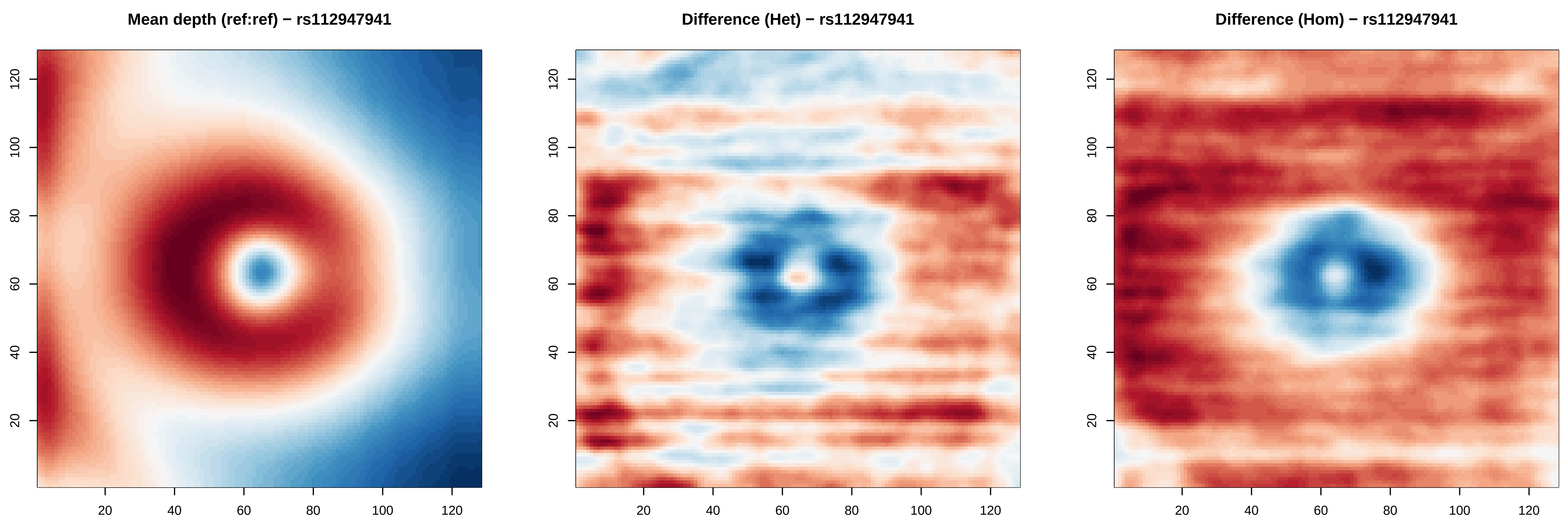
Mean depth (ref:ref) − rs112947941
Difference (Het) − rs112947941
Difference (Hom) − rs112947941
20
40
60
80
100
120

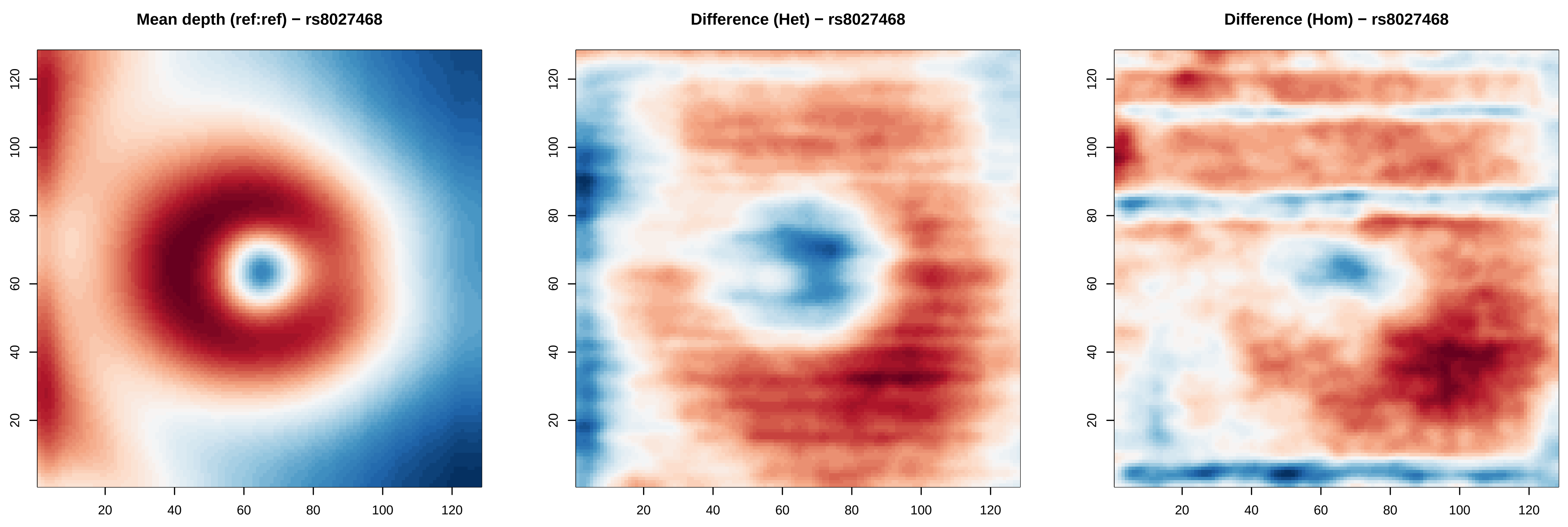

Mean depth (ref:ref) − rs8027468
Difference (Het) − rs8027468
Difference (Hom) − rs8027468
20
40
60
80
100
120

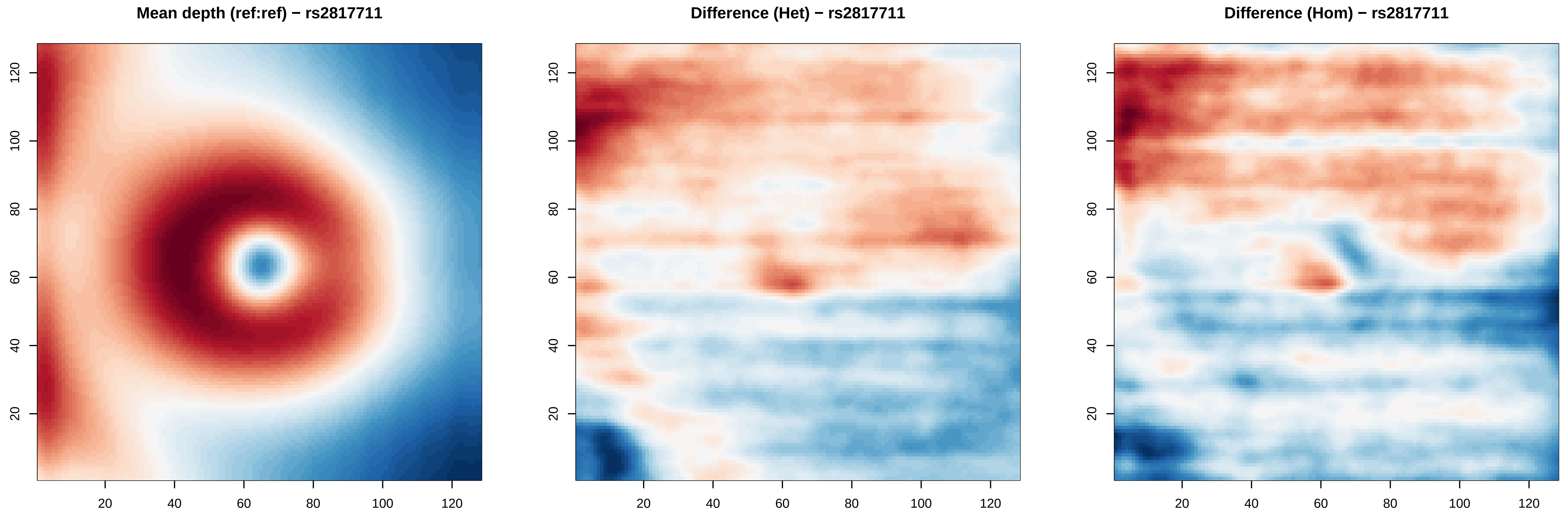

Mean depth (ref:ref) − rs2817711
Difference (Het) − rs2817711
Difference (Hom) − rs2817711
20
40
60
80
100
120

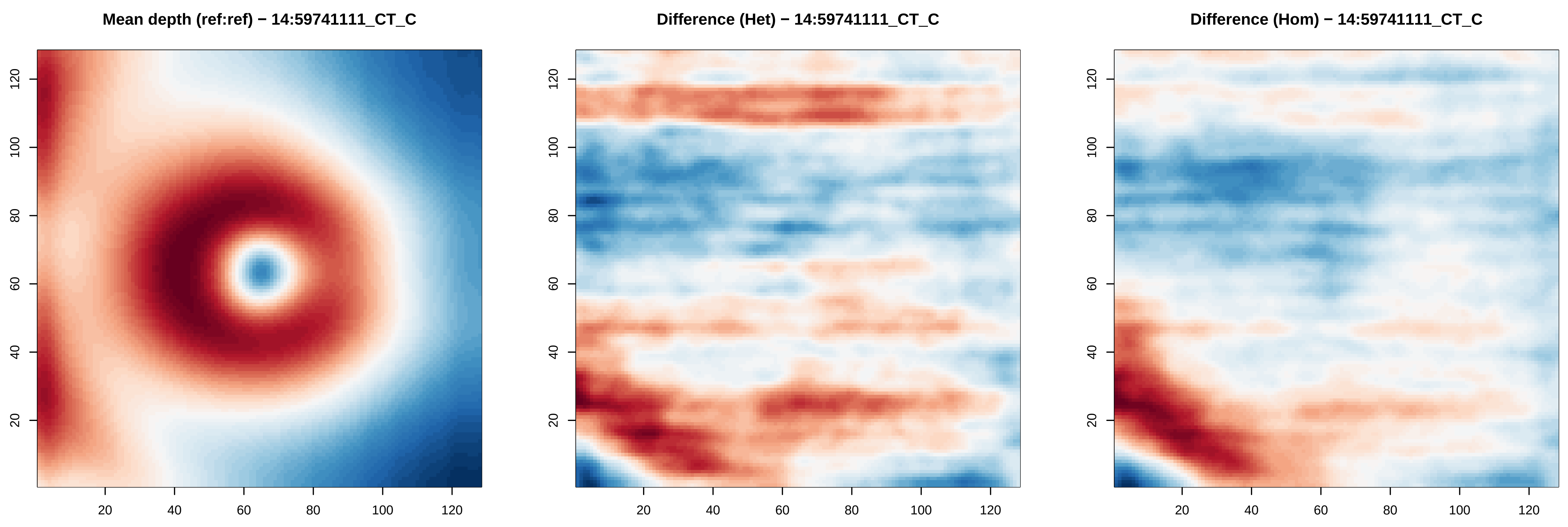

Mean depth (ref:ref) − 14:59741111_CT_C
Difference (Het) − 14:59741111_CT_C
Difference (Hom) − 14:59741111_CT_C
20
40
60
80
100
120

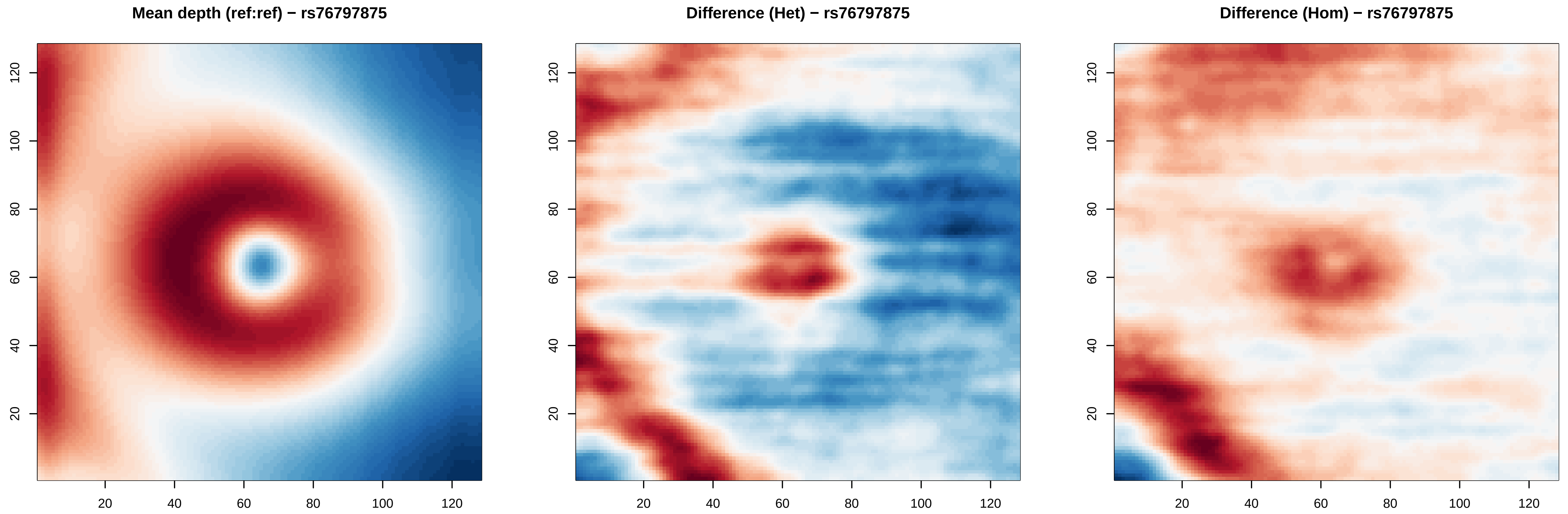

Mean depth (ref:ref) − rs76797875
Difference (Het) − rs76797875
Difference (Hom) − rs76797875
20
40
60
80
100
120

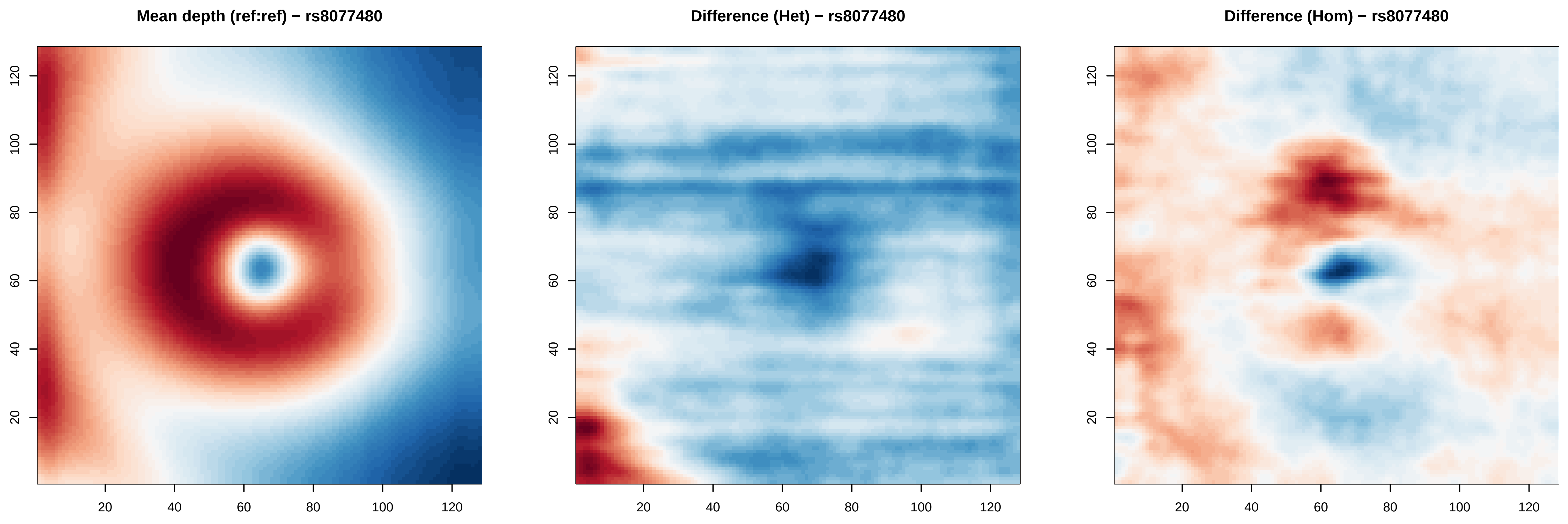
Mean depth (ref:ref) − rs8077480
Difference (Het) − rs8077480
Difference (Hom) − rs8077480
20
40
60
80
100
120

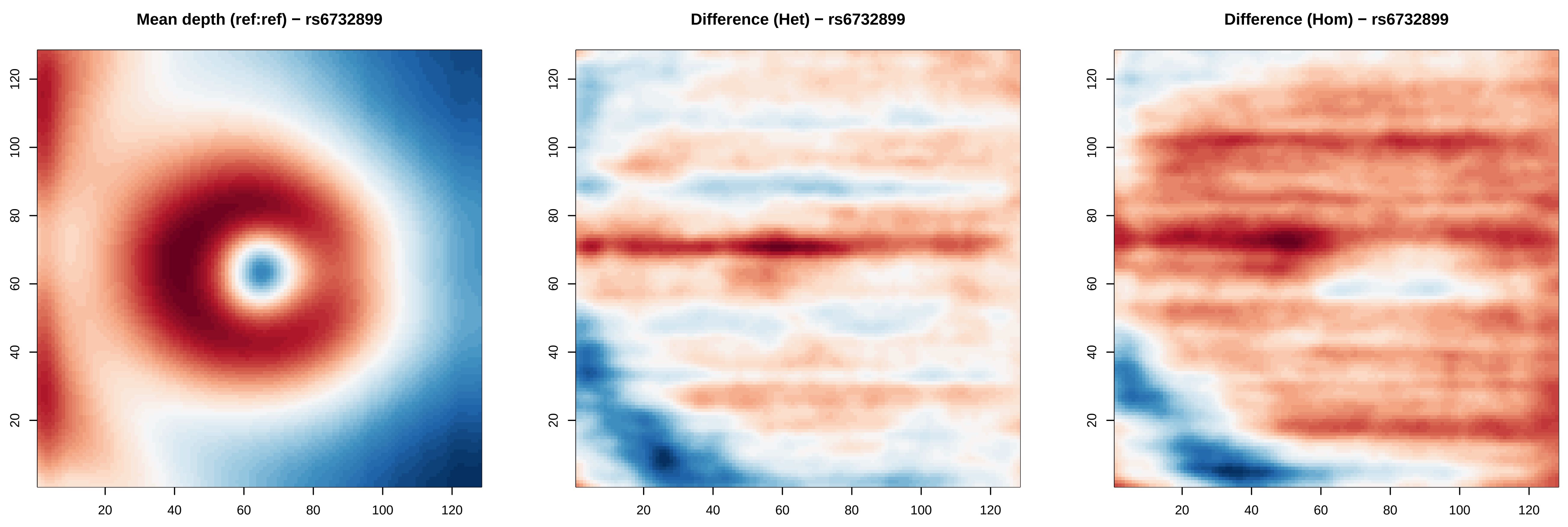
Mean depth (ref:ref) − rs6732899
Difference (Het) − rs6732899
Difference (Hom) − rs6732899
20
40
60
80
100
120

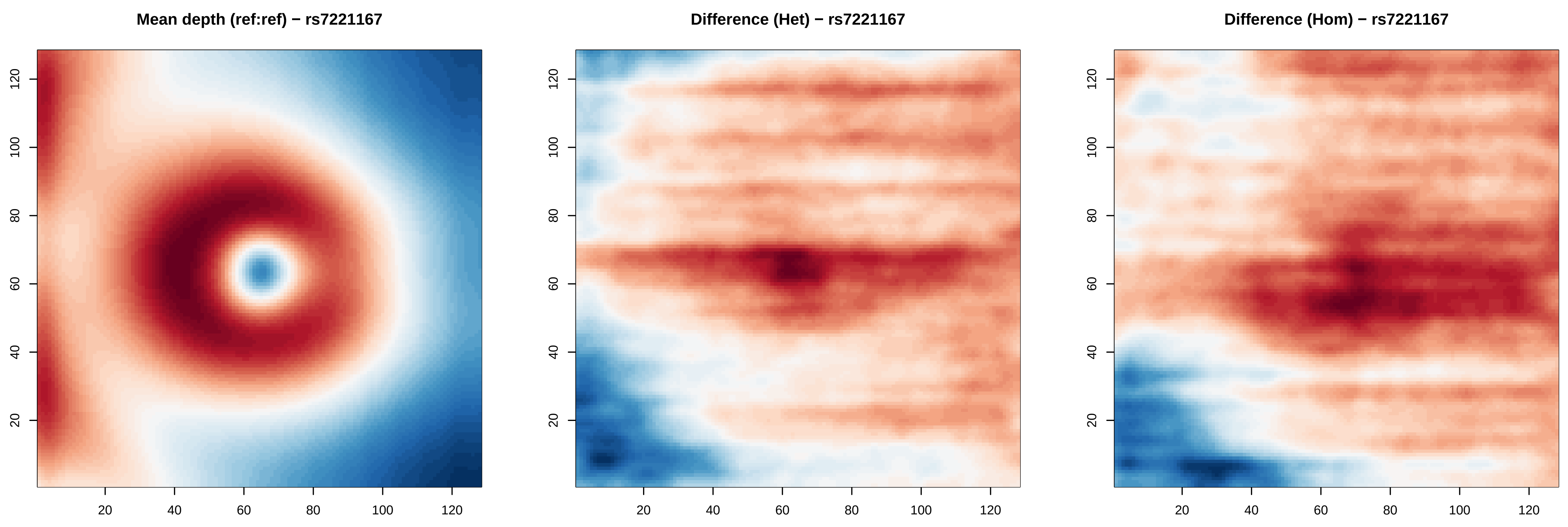
Mean depth (ref:ref) − rs7221167
Difference (Het) − rs7221167
Difference (Hom) − rs7221167
20
40
60
80
100
120

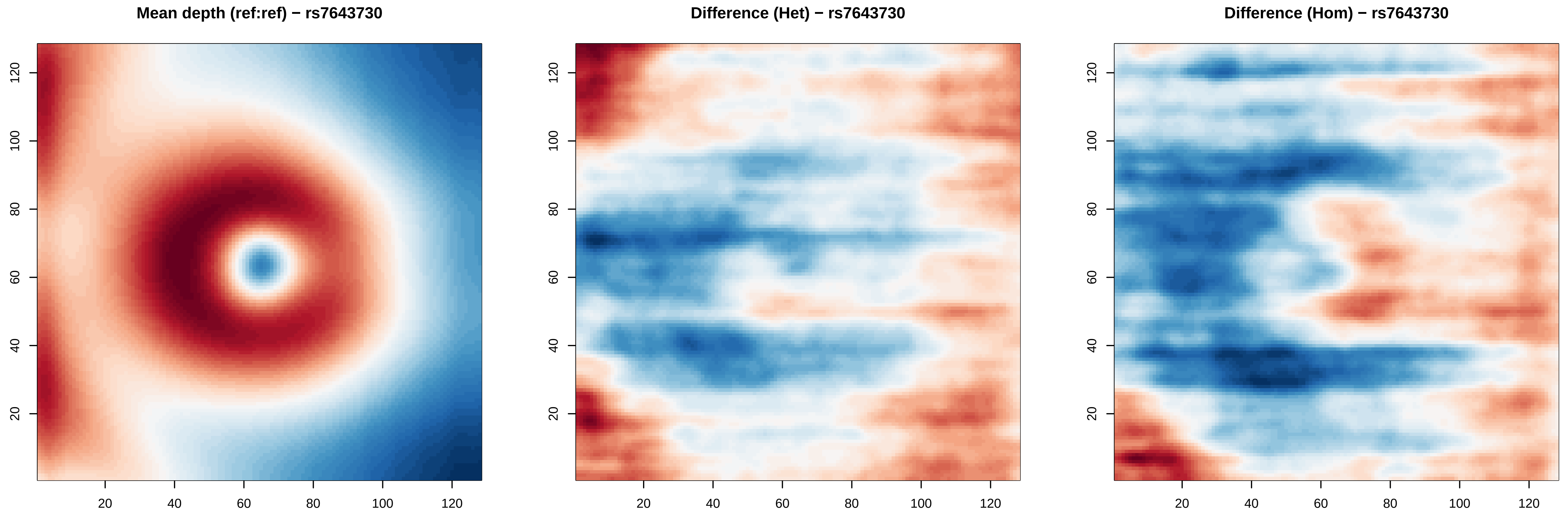
Mean depth (ref:ref) − rs7643730
Difference (Het) − rs7643730
Difference (Hom) − rs7643730
20
40
60
80
100
120

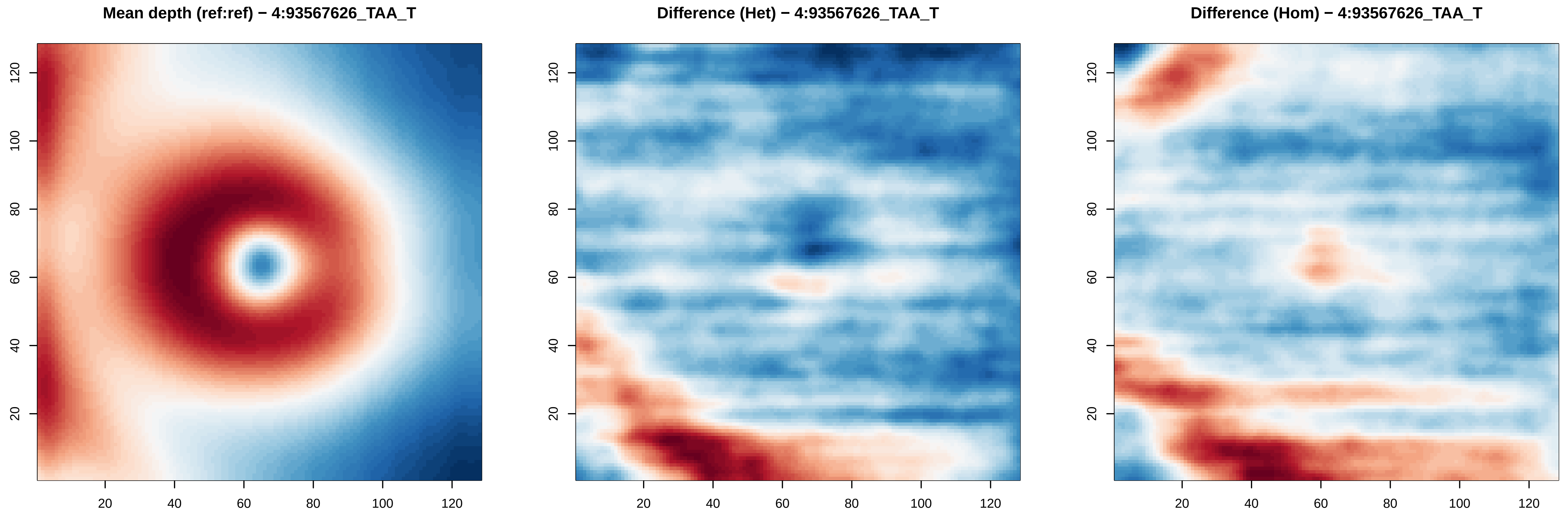
Mean depth (ref:ref) − 4:93567626_TAA_T
Difference (Het) − 4:93567626_TAA_T
Difference (Hom) − 4:93567626_TAA_T
20
40
60
80
100
120

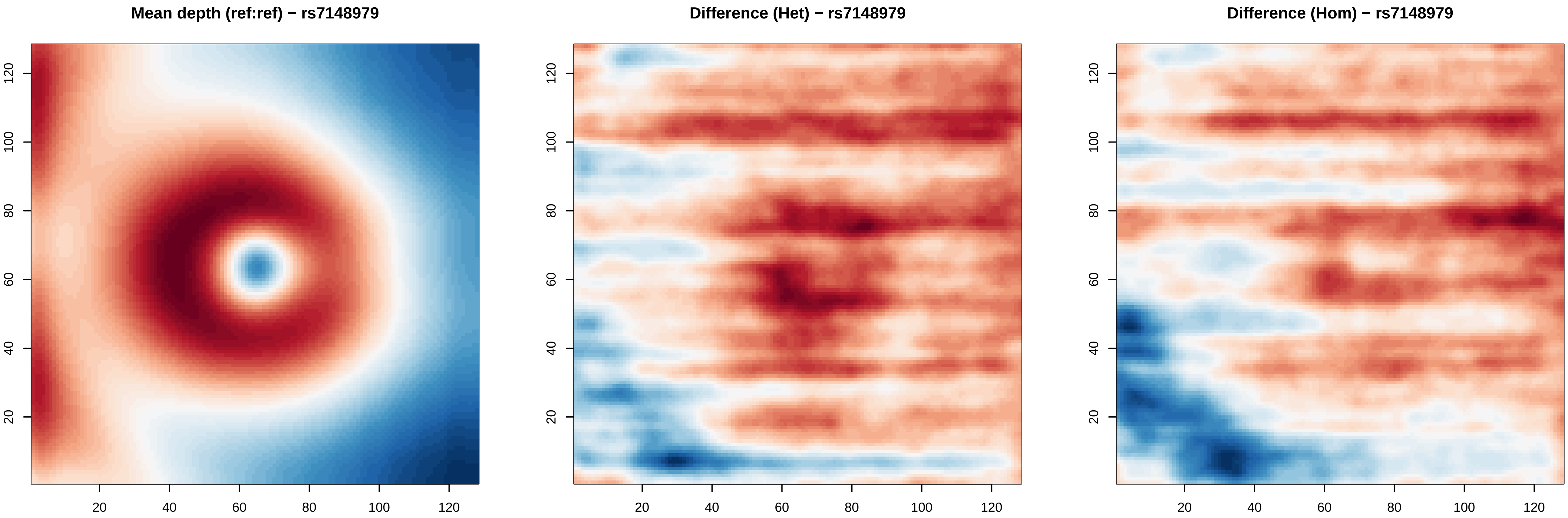

Mean depth (ref:ref) − rs7148979
Difference (Het) − rs7148979
Difference (Hom) − rs7148979
20
40
60
80
100
120

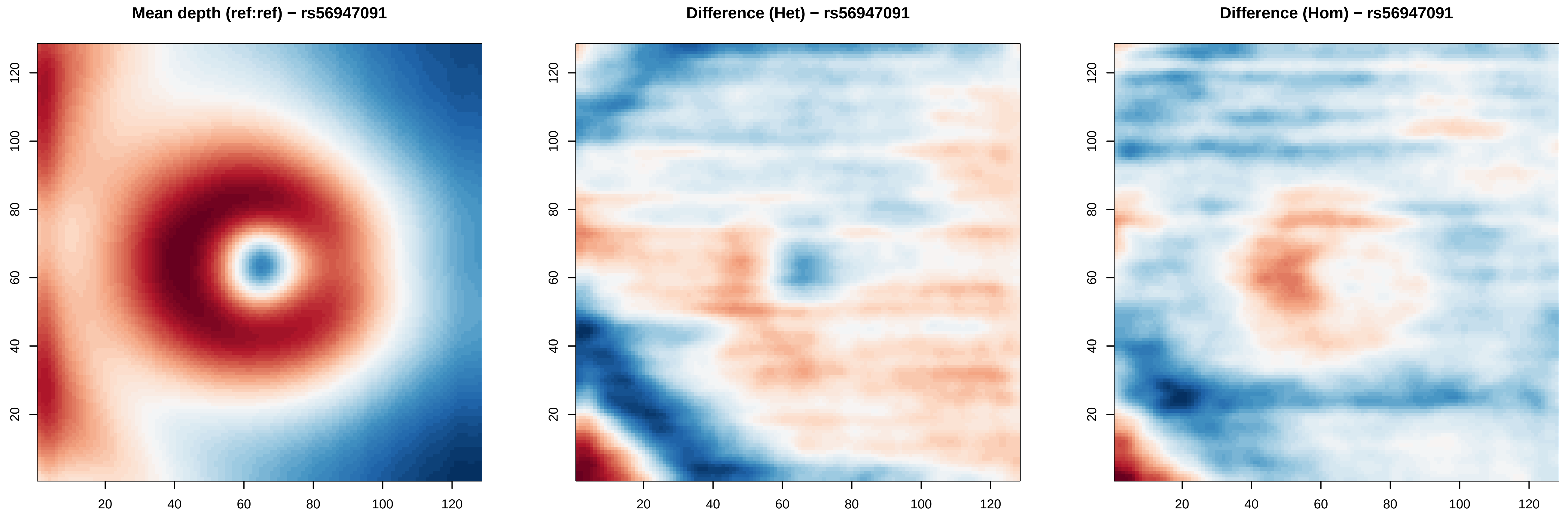

Mean depth (ref:ref) − rs56947091
Difference (Het) − rs56947091
Difference (Hom) − rs56947091
20
40
60
80
100
120

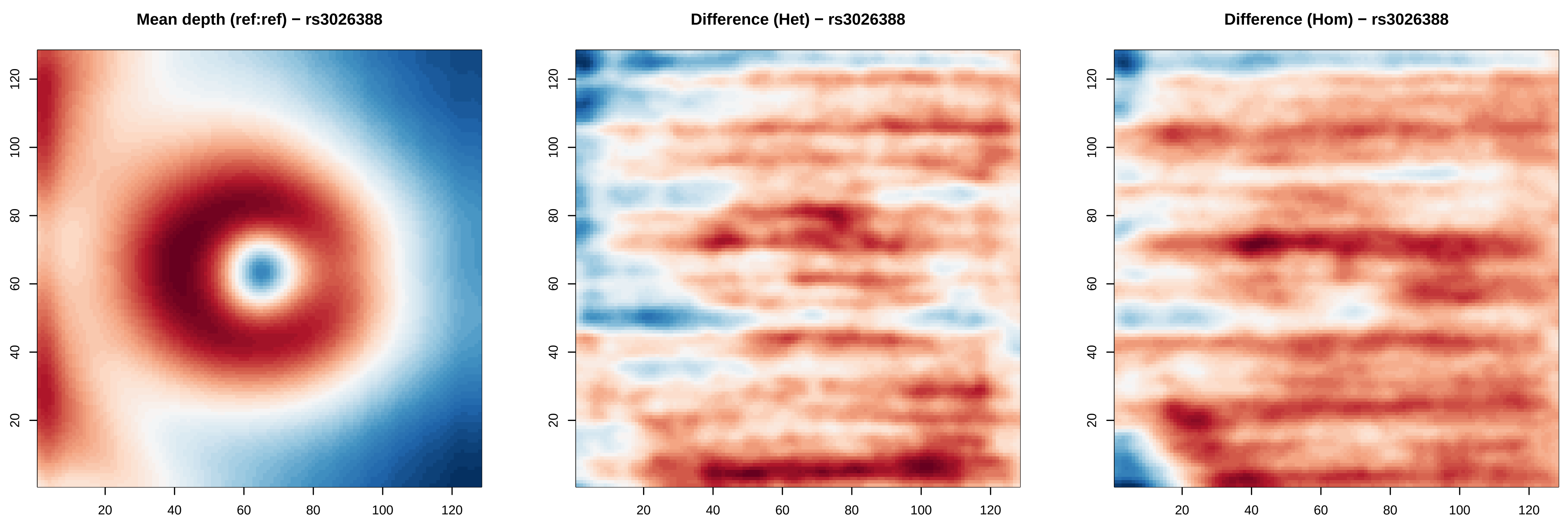
Mean depth (ref:ref) − rs3026388
Difference (Het) − rs3026388
Difference (Hom) − rs3026388
20
40
60
80
100
120

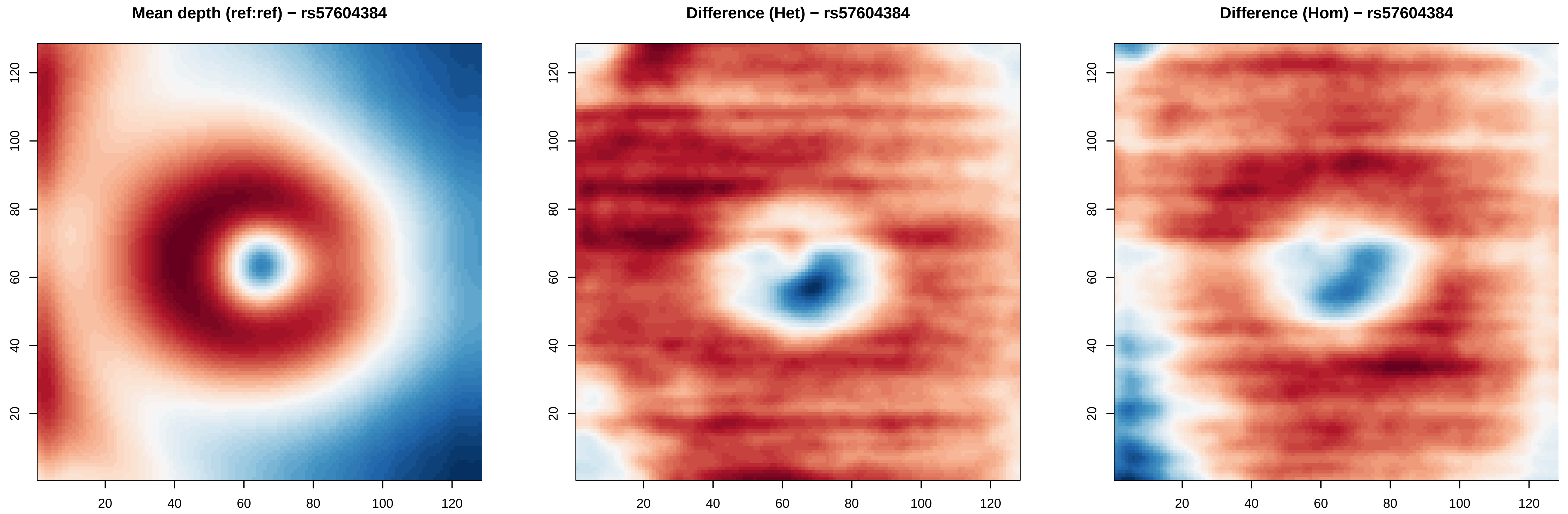
Mean depth (ref:ref) − rs57604384
Difference (Het) − rs57604384
Difference (Hom) − rs57604384
20
40
60
80
100
120

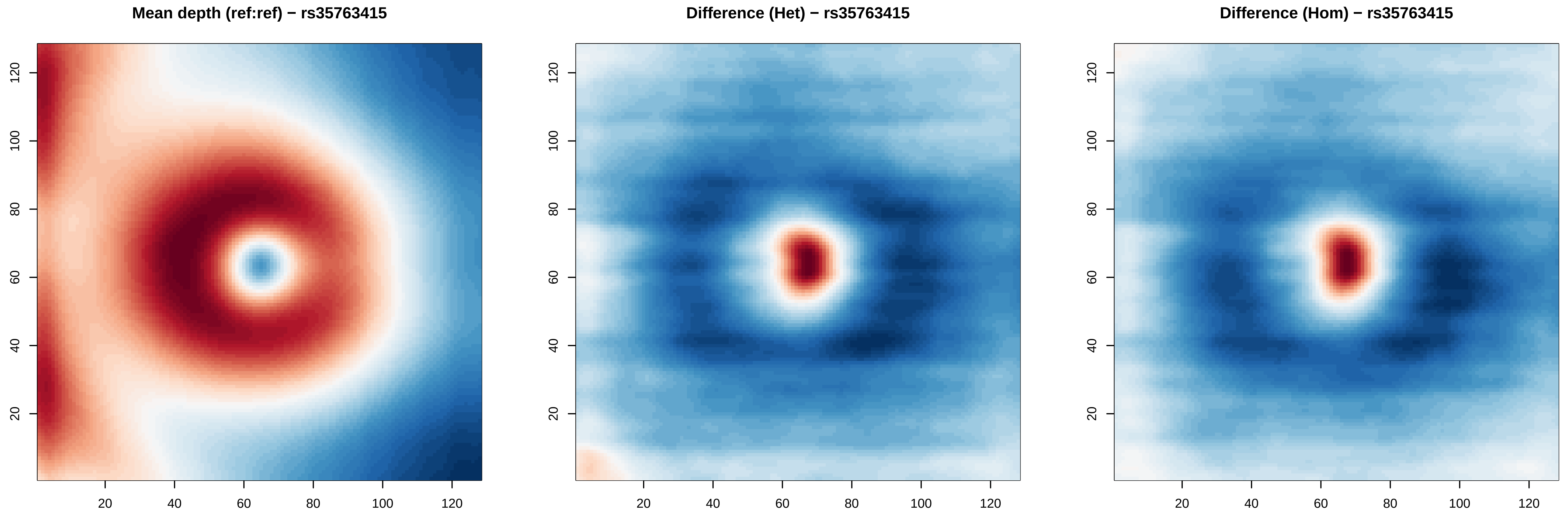

Mean depth (ref:ref) − rs35763415
Difference (Het) − rs35763415
Difference (Hom) − rs35763415
20
40
60
80
100
120

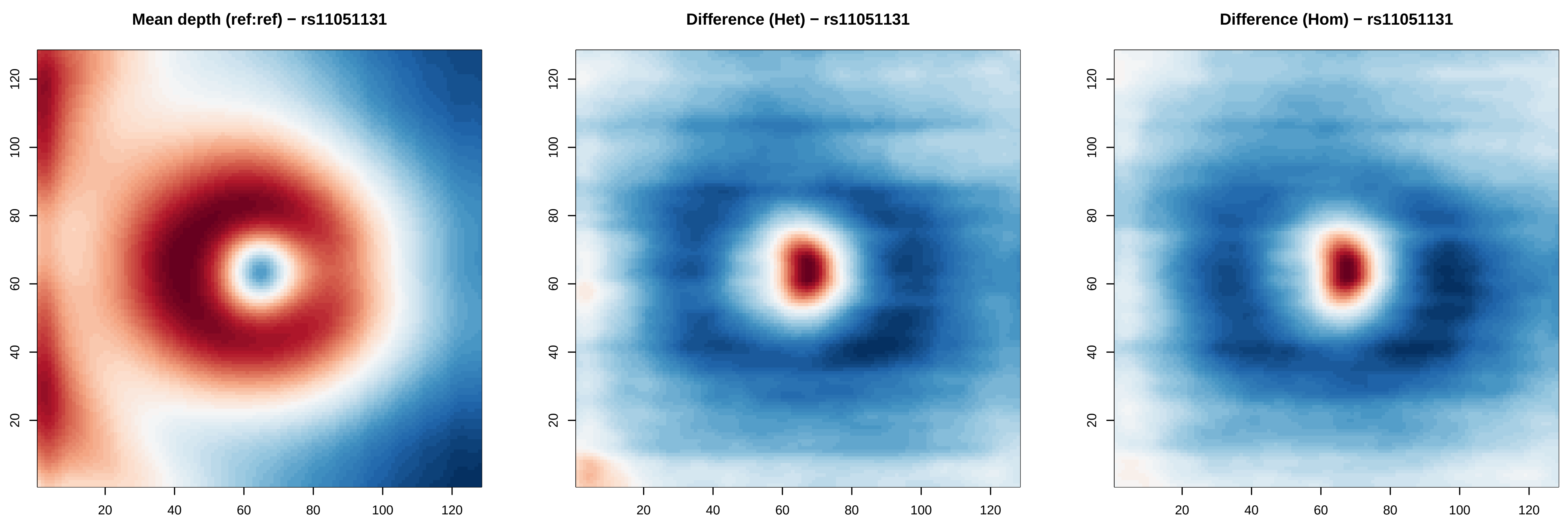

Mean depth (ref:ref) − rs11051131
Difference (Het) − rs11051131
Difference (Hom) − rs11051131
20
40
60
80
100
120

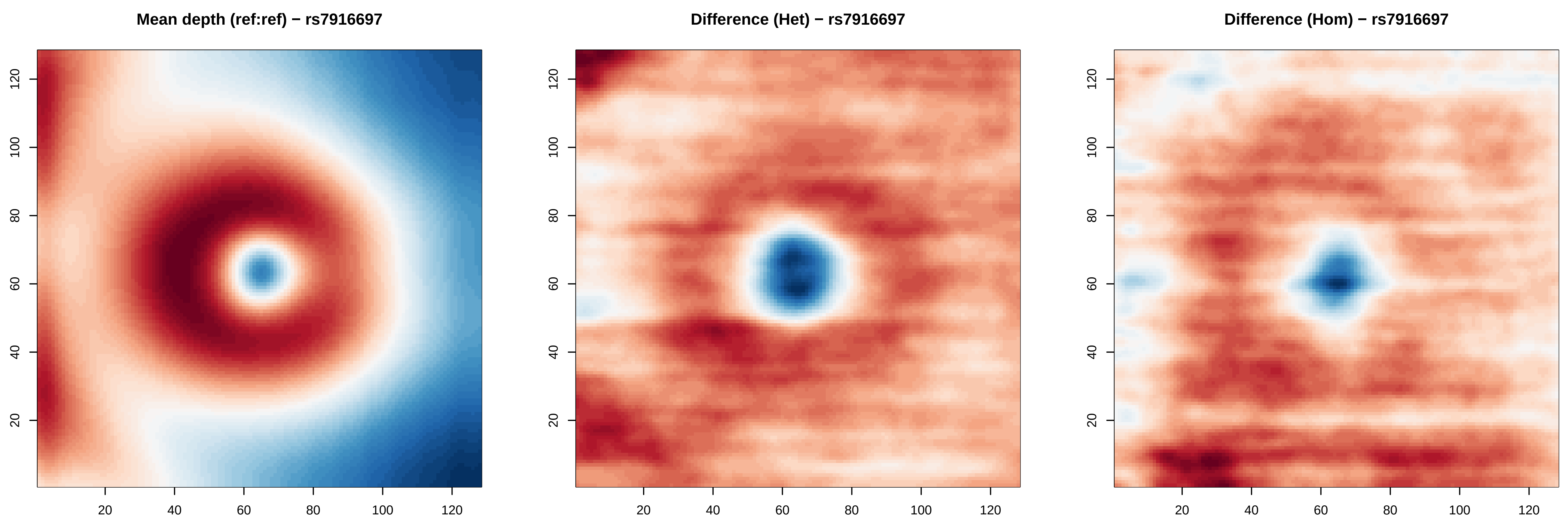

Mean depth (ref:ref) − rs7916697
Difference (Het) − rs7916697
Difference (Hom) − rs7916697
20
40
60
80
100
120

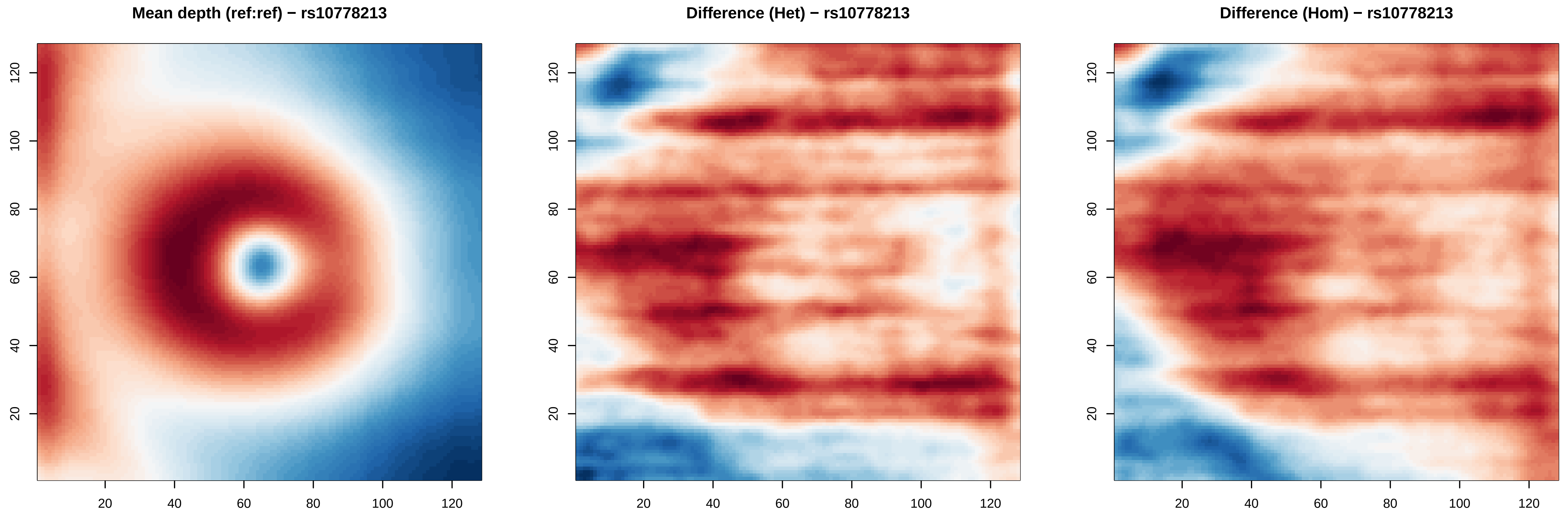
Mean depth (ref:ref) − rs10778213
Difference (Het) − rs10778213
Difference (Hom) − rs10778213
20
40
60
80
100
120

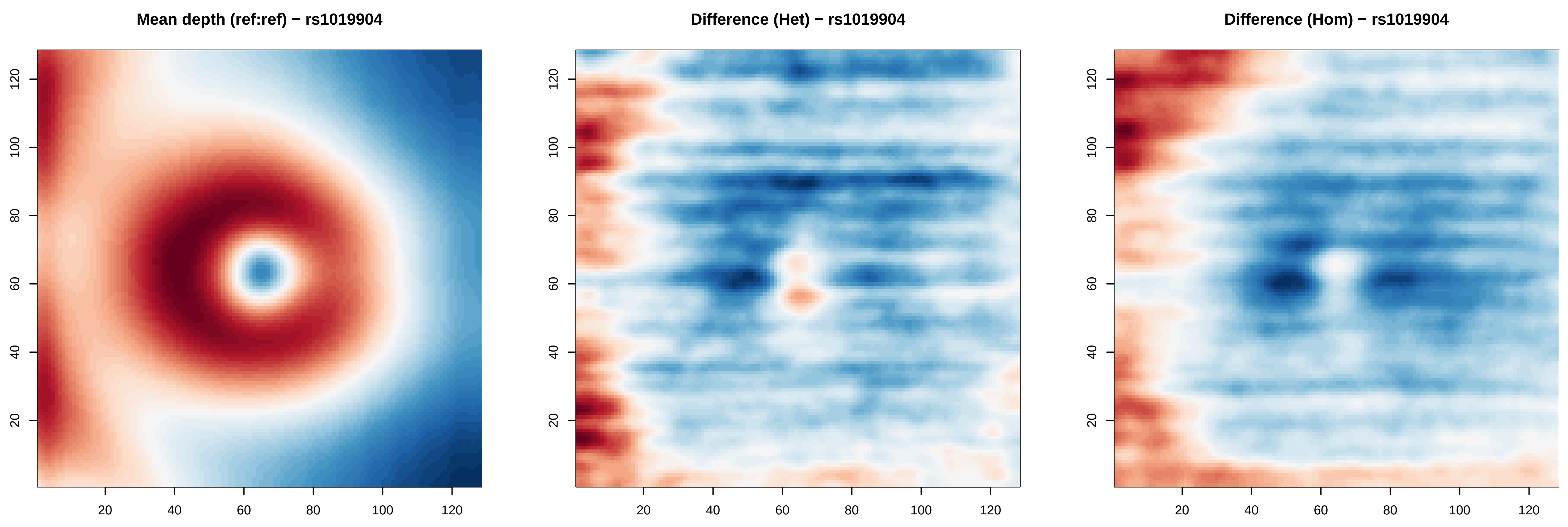

Mean depth (ref:ref) − rs1019904
Difference (Het) − rs1019904
Difference (Hom) − rs1019904
20
40
60
80
100
120

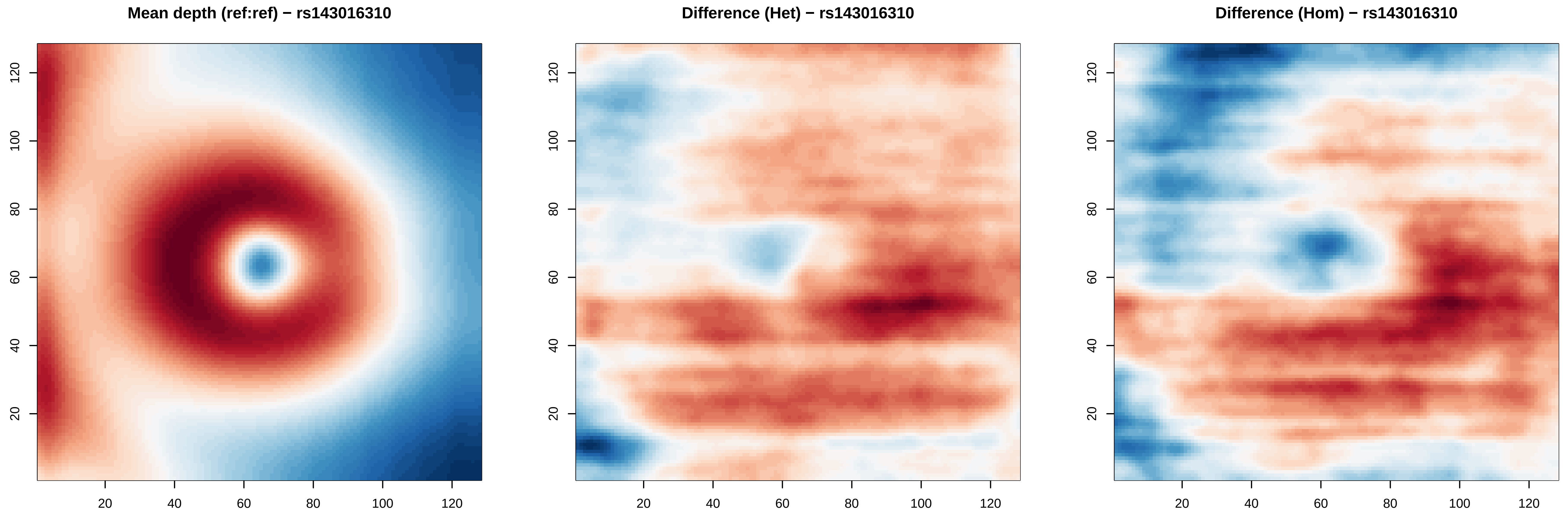
Mean depth (ref:ref) − rs143016310
Difference (Het) − rs143016310
Difference (Hom) − rs143016310
20
40
60
80
100
120

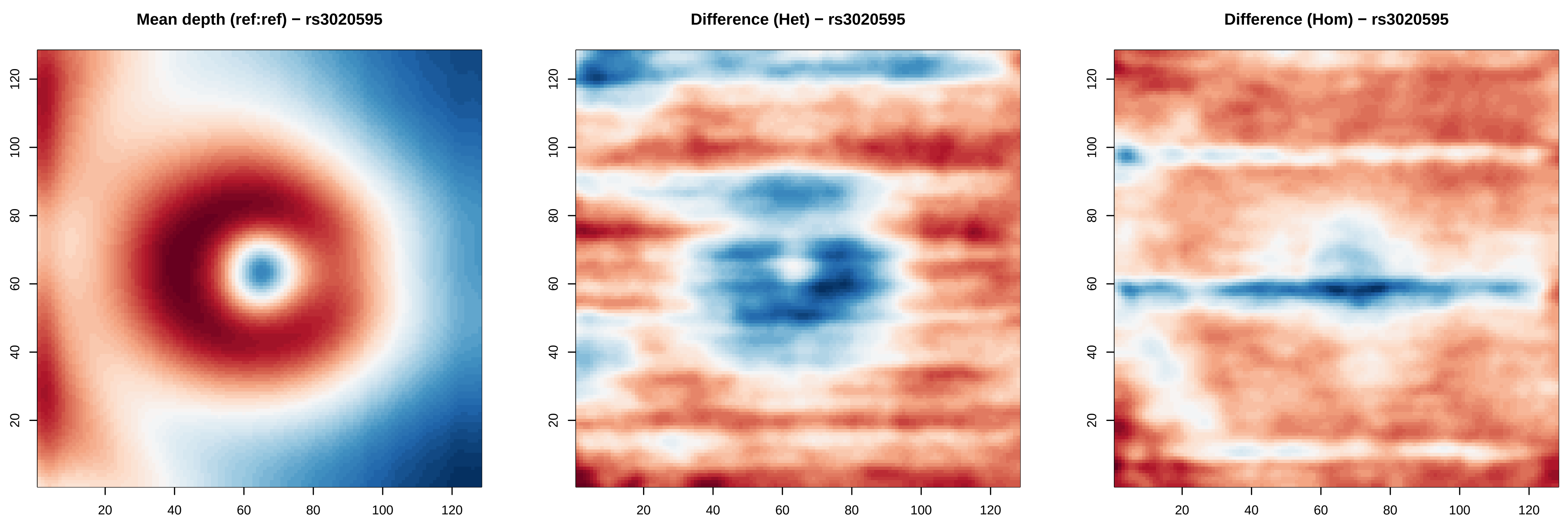
Mean depth (ref:ref) − rs3020595
Difference (Het) − rs3020595
Difference (Hom) − rs3020595
20
40
60
80
100
120

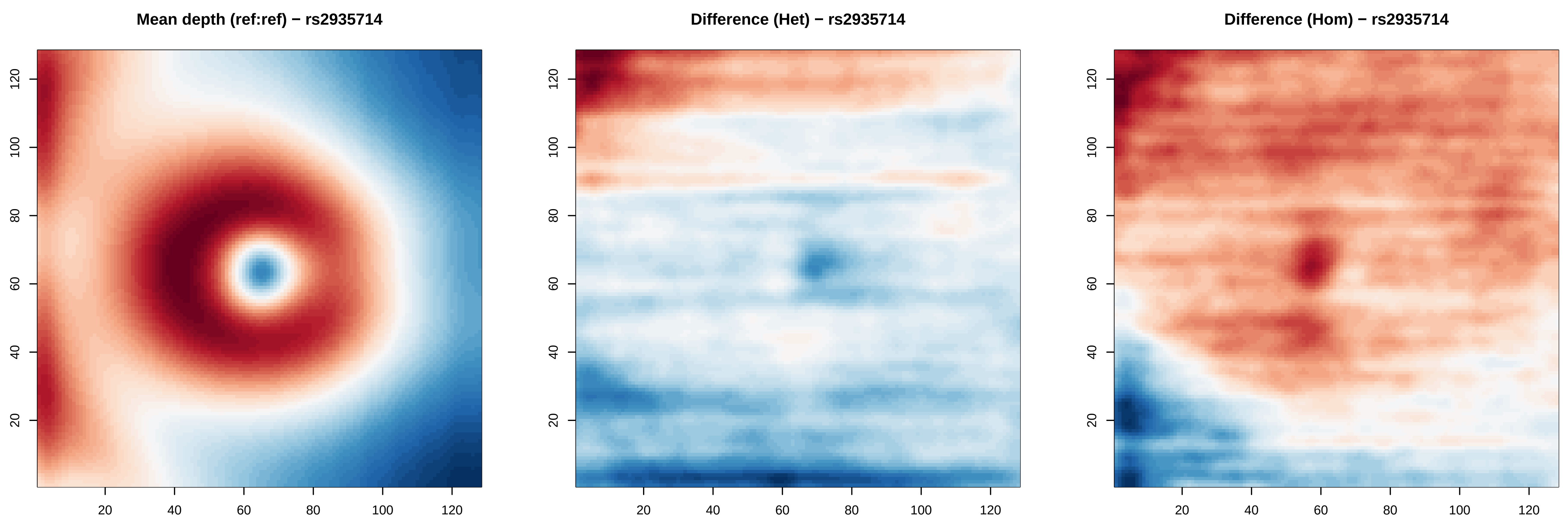
Mean depth (ref:ref) − rs2935714
Difference (Het) − rs2935714
Difference (Hom) − rs2935714
20
40
60
80
100
120

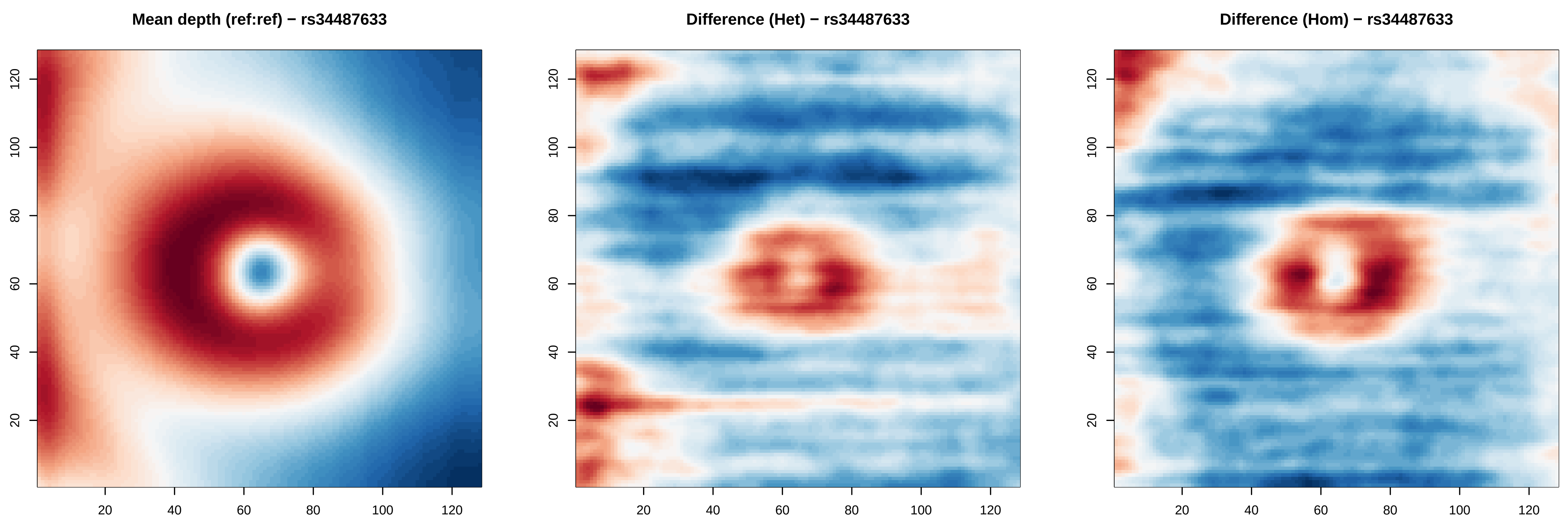
Mean depth (ref:ref) − rs34487633
Difference (Het) − rs34487633
Difference (Hom) − rs34487633
20
40
60
80
100
120

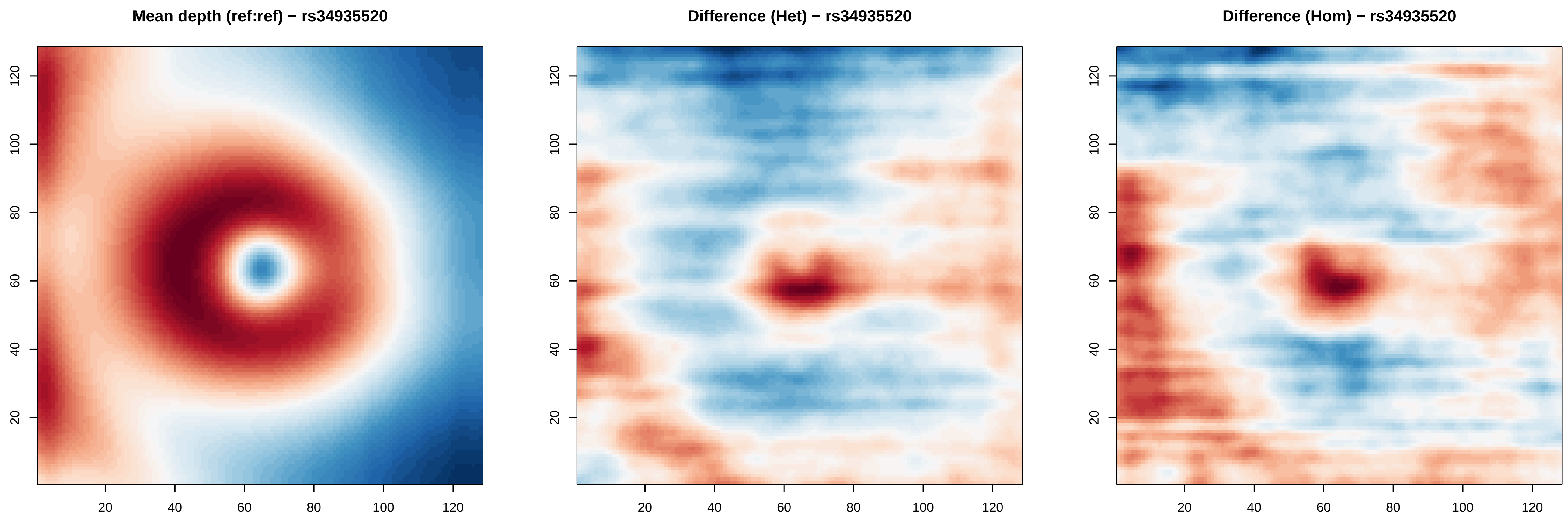
Mean depth (ref:ref) − rs34935520
Difference (Het) − rs34935520
Difference (Hom) − rs34935520
20
40
60
80
100
120

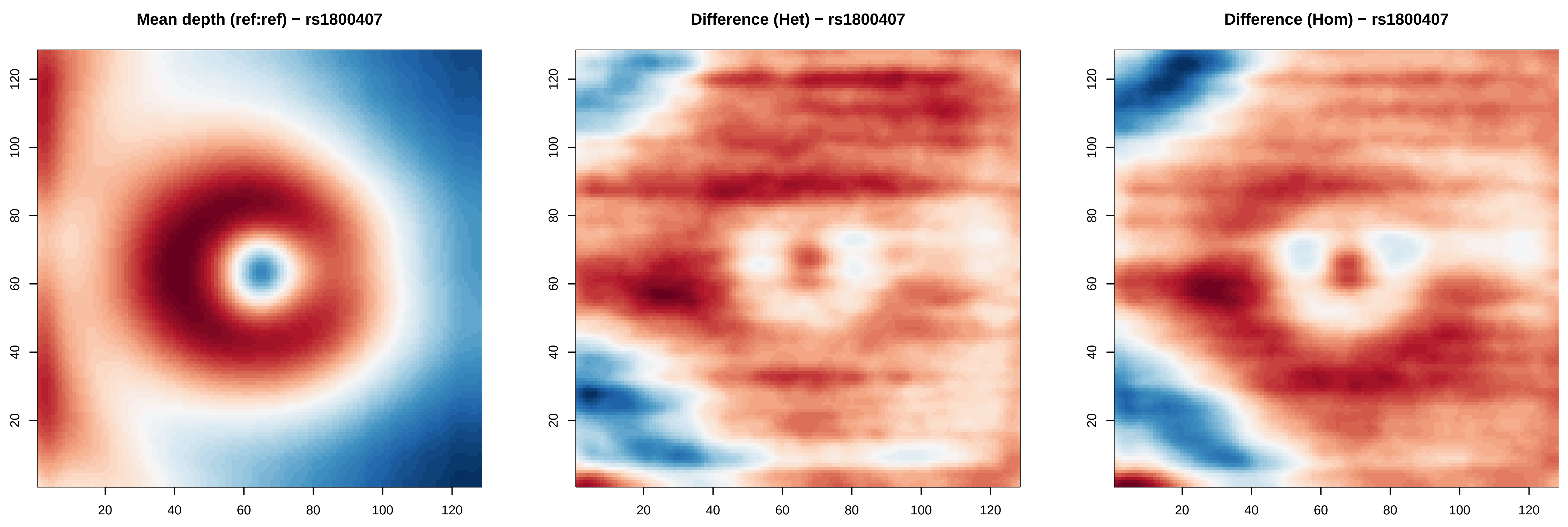
Mean depth (ref:ref) − rs1800407
Difference (Het) − rs1800407
Difference (Hom) − rs1800407
20
40
60
80
100
120

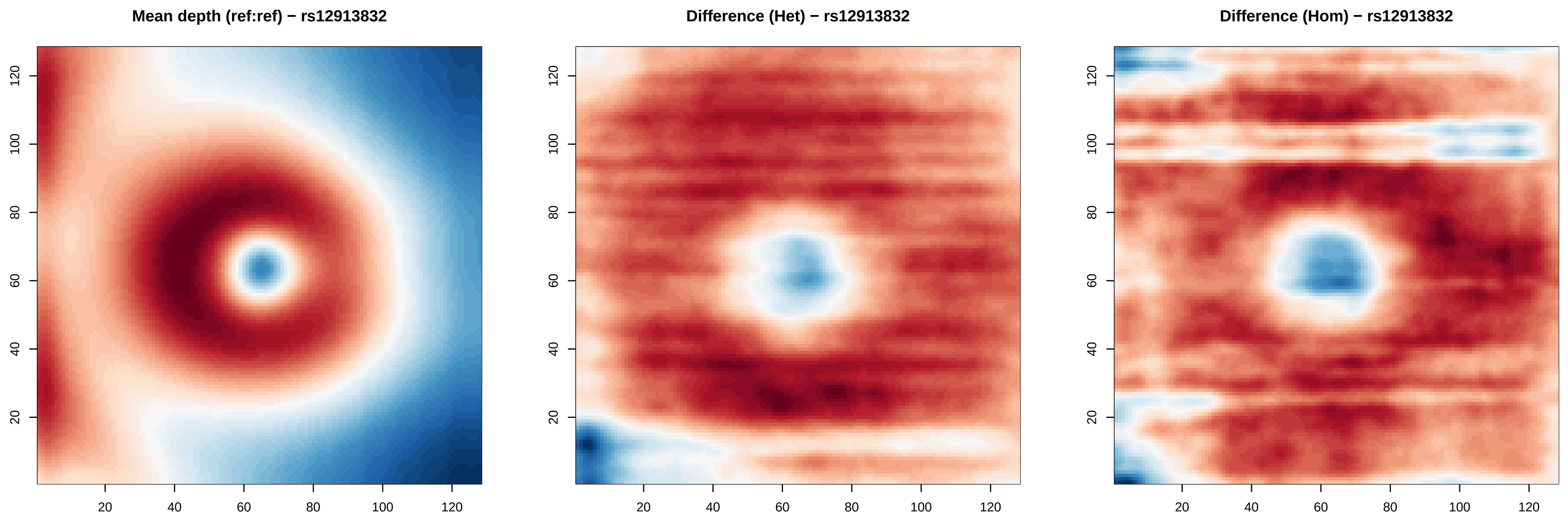

Mean depth (ref:ref) − rs12913832
Difference (Het) − rs12913832
Difference (Hom) − rs12913832
20
40
60
80
100
120

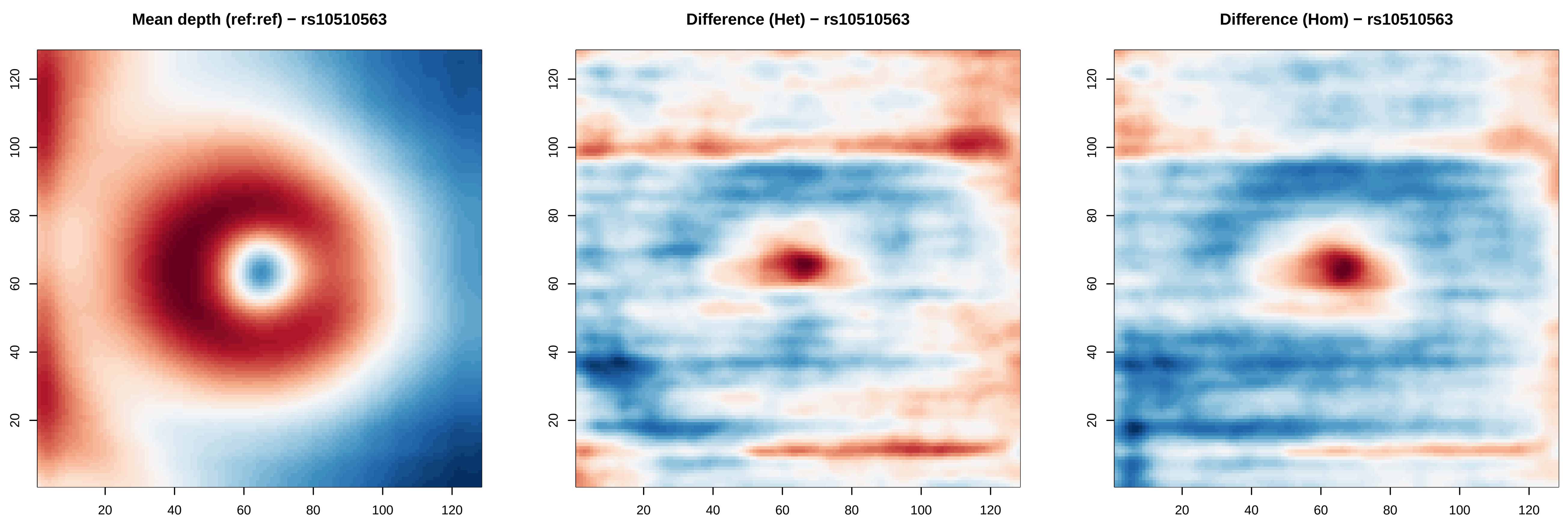
Mean depth (ref:ref) − rs10510563
Difference (Het) − rs10510563
Difference (Hom) − rs10510563
20
40
60
80
100
120

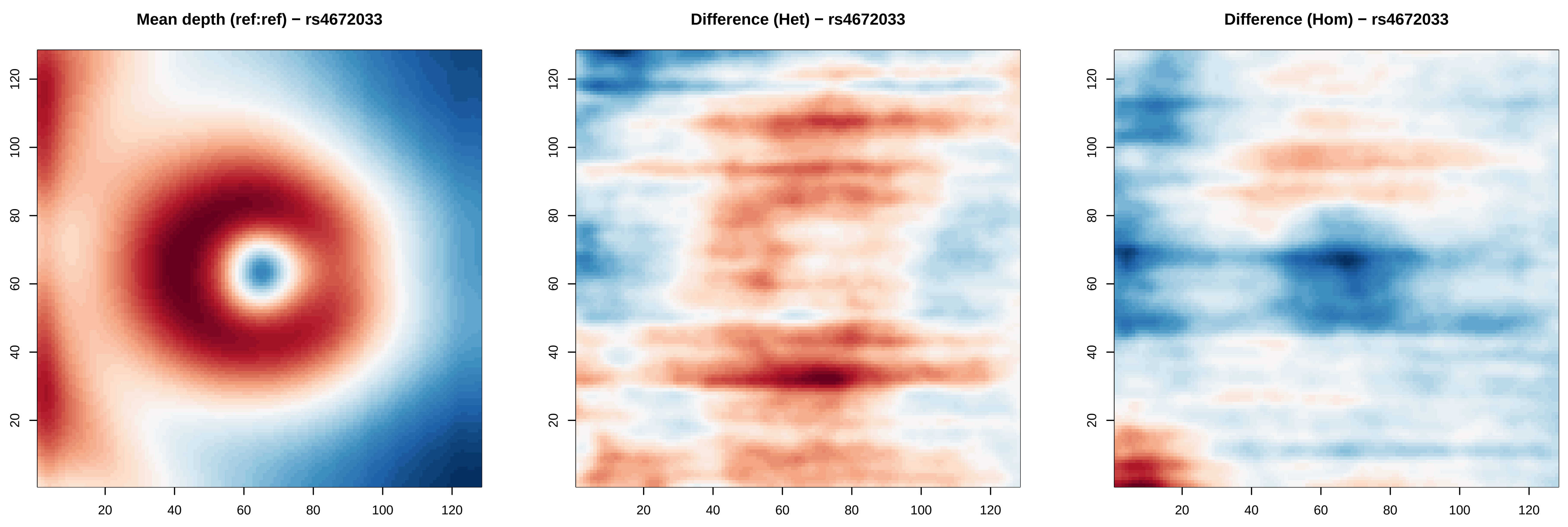
Mean depth (ref:ref) − rs4672033
Difference (Het) − rs4672033
Difference (Hom) − rs4672033
20
40
60
80
100
120

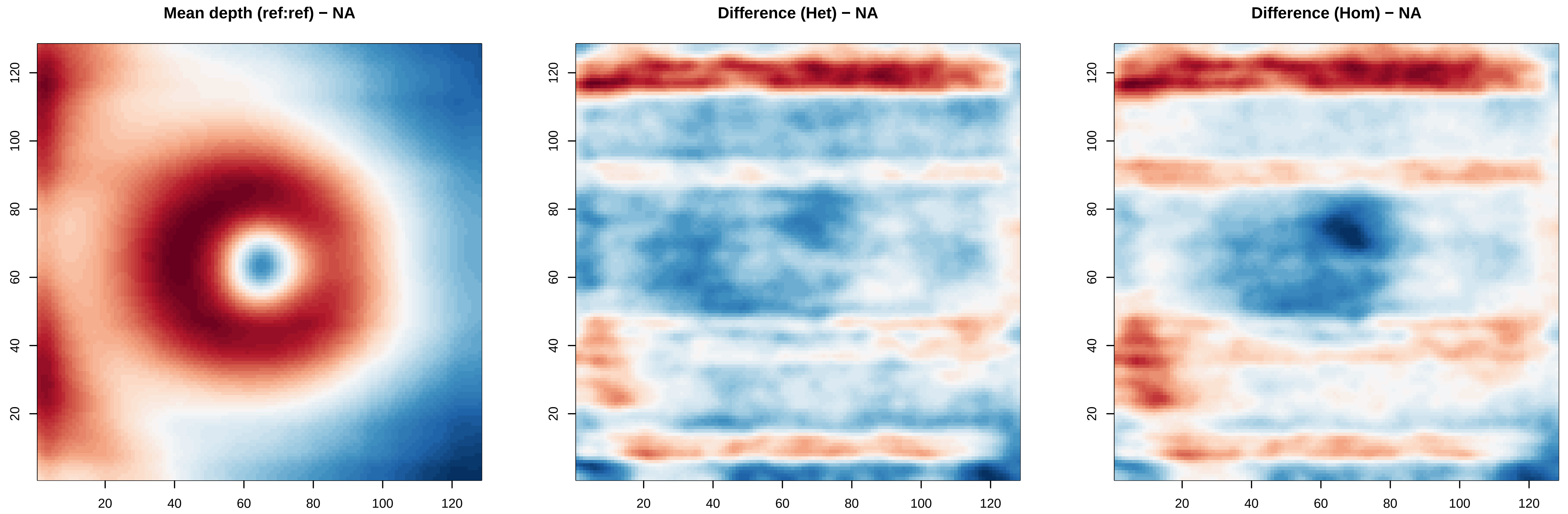

Mean depth (ref:ref) − NA
Difference (Het) − NA
Difference (Hom) − NA
20
40
60
80
100
120
